# Characterising COVID-19 as a Viral Clotting Fever: A Mixed Methods Scoping Review

**DOI:** 10.1101/2020.11.10.20228809

**Authors:** Justin Marley, Nisha Marley

**Affiliations:** Essex Partnership University NHS Foundation Trust; East Suffolk and North Essex NHS Foundation Trust

## Abstract

**Background:** The COVID-19 pandemic has claimed over 1 million lives globally and results from the SARS-COV2 virus. COVID-19 is associated with a coagulopathy. In this mixed-methods PRISMA-compliant scoping review, we set out to determine if ARDS, sepsis and DIC could account for the coagulopathy and if there were any other features of the coagulopathy we could determine so as to inform future research. Methods: We used a search strategy to identify papers with clinically relevant thromboembolic events in COVID-19. We then developed a technique referred to as an Abridged Thematic Analysis (ATA) to quickly identify themes in the papers so as to increase the yield of clinically relevant information. We further developed Validated Abridged Thematic Analysis (VATA) to validate the resulting taxonomy of themes. Finally we developed a number of methods that can be used by other researchers to take forwards this work. Results: We identified 56 studies with 10,523 patients, 456 patients with COVID-19 and thromboembolic events (TBE’s) and 586 thrombembolic events. There were an average of 1.3 TBE’s per patient. There were five main arterial territories with corresponding clinical sequelae: Acute limb ischaemia, myocardial infarcts, strokes, mesenteric ischaemia and pulmonary embolism. We also identified DVT’s. There were two further groups: medical-device-related coagulopathy and dermal lesions. In a subgroup of 119 patients we found mortality ranged from 26% in DVT to 79% in acute limb ischaemia although there was evidence of selection bias in the latter group. All patients were hospitalised and the average age of survivors was 63 versus 73 for those who died. 91/150 patients with TE’s had fever. From the ATA, we identified 16 characteristics of the clotting pathology in COVID-19. From the VATA, we identified 34 mechanisms leading to coagulopathy and grouped them according to Virchow’s triad of vascular damage, stasis and hypercoagulability. Coagulopathy occurred with and without each of ARDS, Sepsis and DIC. We conclude that COVID-19 leads to the syndrome of a viral clotting fever in a subgroup of patients and that the presentation of coagulopathy and fever should raise the possibility of COVID-19 as a differential. We make recommendations for future research studies.

## Introduction

On December 1^st^ 2019, the first patient with Pneumonia of unknown origin was reported in Wuhan, China followed by several cases associated with the Huanan seafood market reported by the Health Commission of Hubei province on December 31st 2019 (Gralinksi and Menachery, 2020)(Habibzadeh and Stoneman, 2020)(Peng et al, 2020). The illness was associated with a novel Coronavirus, SARS-COV2 which was isolated and characterised (Zhu et al, 2020). The resulting infection has been termed COVID-19.

Epidemiological characteristics of COVID-19 have been characterised (Fauver et al, 2020)(Read et al, 2020)(Ryu et al, 2020)(Adam et al, 2020) and facilitated using SARS-COV2 genome sequencing. (Verity et al, 2020) estimated the overall infection fatality rate in China at 0.66% (95% CI 0.39-1.66). The case fatality ratio varies according to age with 0·32% (0·27–0·38) in those aged less than 60 years and 13·4% (11·2–15·9) in those aged 80 years or older. A number of risk factors were identified for more severe illness including chronic cardiac disease, diabetes and obesity (Docherty et al, 2020)(Yang et al, 2020). Factors affecting the transmission dynamics of SARS-COV2 have been identified and informed public health measures (Cevik et al, 2020) (De Deyn et al, 2020)(Javid and Balaban, 2020).

Asymptomatic presentation has been well described (Oran and Topol, 2020) and is essential in understanding transmission dynamics but fever is recognised as a prominent clinical feature occurring in 98.6% of 138 symptomatic patients in an early study (Wang et al, 2020). Other studies have confirmed the significance of fever which has been reported in 83-98% of symptomatic cases (Wiersinga et al, 2020)(Chen et al, 2020)(Huang et al, 2020(a))(Guan et al, 2020)(Alizadehsani et al, 2020). Further studies have characterised the longer-term consequences of COVID-19 (Mahase, 2020) which has been termed ‘Long COVID’. Whilst the classical presentation of COVID-19 is described as bilateral pneumonia with acute respiratory distress syndrome in more severe cases, extrapulmonary manifestations have been described including neurological, renal, dermatological, cardiac and gastrointestinal symptoms (Gupta et al, 2020)(Elmunzer et al, 2020)(Adukia et al, 2020). Silent hypoxia has been described in COVID-19 (Wilkerson et al, 2020) and can present a clinical challenge in routine practice.

An abnormal coagulation state is an important finding in COVID-19 (Tang et al, 2020)(Wu et al, 2020) (Connors and Levy, 2020) with similarities to other coronavirus infections including MERS (Mackay and Arden, 2015) and SARS (Hui et al, 2020). D-Dimer levels were found to be elevated in a study of 1099 patients with COVID-19 (Guan et al, 2020) and other coagulation abnormalities have been identified (Boccia et al, 2020).

In this scoping review we focus on the COVID-19-related coagulopathy and it is necessary to set the scene in this introduction. We firstly provide an overview of SARS-COV2 and the pathological findings identified in COVID-19. We then outline the typical host response to a viral infection before moving on to discuss the clotting cascade and then Virchow’s triad which we have used as a framework for our analysis. We then consider three important pathologies in COVID-19 which have featured in the early discussions around the COVID-19-related coagulopathy – ARDS, DIC and septicaemia and which form a basis for our initial analysis of the literature.

### SARS-COV2

Coronaviruses are single-stranded RNA viruses (V’kovski et al, 2020) and are positive sense which means they can be read as mRNA by the ribosomes in the host cell and then translated into proteins. Taxonomically SARS-COV2 is a member of the suborder Coronoavirineae and the genus Betacoronaviridae which specifically infect mammals (V’kovski et al, 2020). Although viruses are not classified as living organisms, the sequence of events leading to replication is described as a life cycle (Ryu, 2017). (Ryu, 2017) outlines six steps in the virus life cycle: Attachment, penetration, uncoating, gene expression and genome replication, assembly and release.

During attachment, the virus attaches to the surface of the host cell followed by entry into the cytoplasm (penetration) and shedding of the viral capsid (uncoating). The viral RNA is then read (gene expression and genome replication) and then the gene products are assembled into a virion (assembly) and released from the cell (release).

Turning to coronaviruses there are four main structural proteins - spike, envelope, membrane and nucleocapsid (V’kovski et al, 2020)(see also Figure 1). The SARS-COV2 spike protein is divided into two functional components: S1 binds the receptor on the host cell and S2 facilitates the fusion of the virus with the host cell membrane. Like SARS, SARS-COV2 binds the ACE-2 receptor on the host cell. The ACE-2 receptor is found in tissues throughout the body (Hikmet et al, 2020). SARS-COV2 also requires the priming action of trans-membrane serine protease 2 (TMPRSS2) which activates the SARS-COV2 spike protein so that the virus can fuse with the cell membrane (Thunders and Delahunt, 2020)(Hoffman et al, 2020).

**Figure 1.**
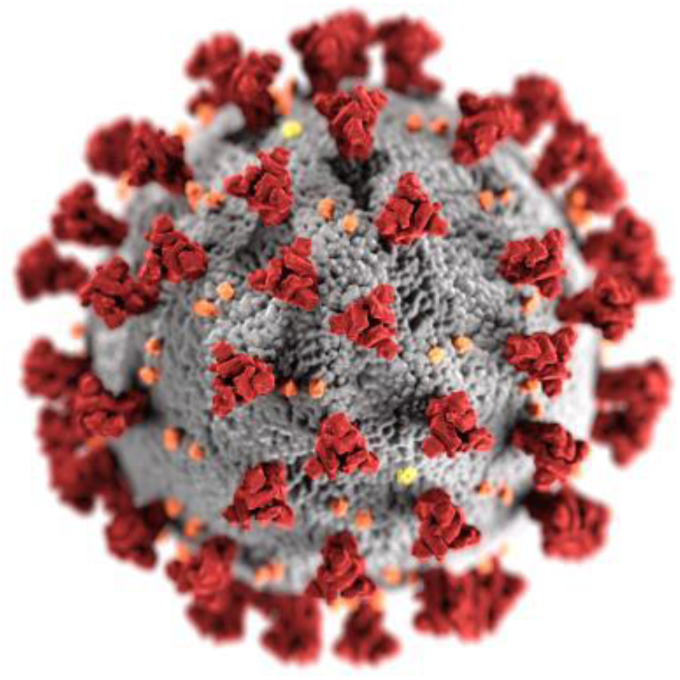
Illustration of SARS-COV2 Virus: Authors CDC/ Alissa Eckert, MSMI; Dan Higgins, https://phil.cdc.gov/Details.aspx?pid=23312, Image in Public Domain.

The degree of glycosylation of the S-protein is thought to interfere with the adaptive immune response by reducing the likelihood of antibody formation (V’kovski et al, 2020). The open reading frames coding for the structural proteins are contained within one third of the genome and the genome also codes for accessory proteins (V’kovski et al, 2020). The virus then appropriates the cell machinery to generate replication organelles including double-membrane vesicles. Viral RNA is transferred into the organelles which offer protection against the host cell’s cytosol-based RNA-sensing immune mechanisms (V’kovski et al, 2020). SARS-COV2 has 79-86% sequence homology with SARS-COV or SARS-COV-like viruses (Lu et al, 2020)

A further consideration is the effect of the virus on the host and this is inferred from a range of studies including post-mortem studies. (Cardot-Leccia et al, 2020) found evidence of reduced pericytes in alveolar capillaries together with thinning of the walls of the alveolar capillaries and venules. (Bouhaddou et al, 2020) present the results of a phosphoproteomics survey in SARS-COV2 infected cells and found evidence of induction of cell cycle arrest and p38 MAPK activation amongst other findings.

(Bösmüller et al, 2020) in a post-mortem series (n=4) found evidence of progression of pulmonary pathology with findings typical of ARDS in the more severe cases of pathology. In milder pathology they found evidence of increased mRNA expression of IL-1 beta and IL-6 as well as capillaritis with the presence of neutrophils and also microthromboses in the capillaries. (Tabary et al, 2020) review the pathological findings in COVID-19 and summarise the findings in the lungs, gastrointestinal tract, liver, kidney, skin, heart blood, spleen and lymph nodes, brain, blood vessels and placenta. Hepatocyte degeneration is noted in the liver as well as an altered vascular structure. Diffuse alveolar damage and lymphocyte infiltration is found in the lungs. Intramural non-occlusive thrombi and fibrin deposition are found in the placenta. Necrosis of the keratinocytes and Langerhans cell nests are found in the skin. Inflammatory cell infiltrates and endotheliitis are found in the blood vessels. Sinus fibrosis and white pulp atrophy are found in the spleen. Axonal injuries and leukocyte infiltration are found in the CNS. Proximal acute tubule injury, tubular necrosis and interstitial fibrosis are found in the kidneys. The post-mortem findings will represent the more severe end of COVID-19 in general depending on the cause of death and so there are limitations on the generalisation to the milder course of the illness.

### The Host Response to Viral Infections

The human immune system is broadly divided into the adaptive and innate immune response and what follows is a simplified account of their function so as to contextualise subsequent material in this paper. The innate immune system responds to novel microbes in the early phase of an infection in contrast with the adaptive immune system which predominantly recognises and responds to a previously encountered microbe. The innate immune system includes physical barriers in the body together with a range of cells including phagocytes (neutrophils and macrophages), mast cells, natural killer cells and the complement cascade (Abbas et al, 2018). The adaptive immune system consists mainly of B and T-lymphocytes. The T-lymphocytes recognise antigens on microbes, referred to as epitopes upon which the T-cell will clone itself to increase the number of circulating T-cells capable of recognising the antigen (Abbas et al, 2018). In this case the T-cell has not previously encountered the microbe and is thus referred to as a naieve T-cell although any subsequent encounters will result in a more effective initial adaptive immune response. T-cells are further divided into helper T-cells which activate B-cells to respond to antigens and cytotoxic T-cells which are capable of killing other cells. T-cells and B-cells have antigen-receptors on their cell surface but B-cells are also capable of secreting antibodies that mirror their cell-surface antigen-receptor (Abbas et al, 2018). Antibody secretion results in binding of the antibody to the antigen in a process referred to as opsonisation. Opsonisation causes the affected antigen-containing body (e.g. cell) to be targeted by phagocytes which proceed to kill target cells.

### The Clotting Cascade

Coagulation is the process by which a liquid changes to a solid or semisolid state. Coagulation of blood is essential in haemostasis, the process which prevents blood leaking from damaged blood vessels. The haemostatic system is a balance between procoagulant and anticoagulant mechanisms (Konkle, 2017). On the one hand the platelets adhere to damaged surfaces and aggregate whilst fibrin forms a clot. Fibrin has complex physical properties and the conditions in which fibrin clots are formed determine the structure of the clot which in turn influences the fibrinolytic susceptibility (Wolberg et al, 2012).

There are a number of anticoagulant mechanisms that provide a counterbalance. There are four main antithrombotic systems – the fibrinolytic system and the systems involving protein C & S, tissue factor pathway inhibitor (TFPI) and antithrombin. The fibrinolytic system degrades fibrin clots in a complex interplay which features the activation of plasminogen to plasmin which then acts on fibrin. Tissue plasminogen activator activates plasminogen.

The clotting cascade is the mechanism by which clotting is initiated and amplified in response to triggers. The clotting cascade can in the simplest form be divided into the intrinsic pathway, the extrinsic pathway and the common pathway (see Figures 2-4). The clotting cascade consists of a number of clotting factors. Historically the clotting factors received many names with one factor receiving 14 different names (Giangrande, 2003). The nomenclature of the clotting factors was rationalised in a series of meetings of the International Committee for the Nomenclature of Blood Clotting Factors between 1955 and 1958 and resulted in the use of Roman numerals. A selection of clotting factors with Roman numeral equivalents is shown in Table 1 and (Palta et al, 2014) note that the first four factors are referred to by the clotting factor names rather than the Roman numerals.

**Table 1.** Clotting factors names with equivalent Roman numerals.

| Roman Numeral | Clotting Factor Name |
| --- | --- |
| I | Fibrinogen |
| II | Prothrombin |
| III | Tissue Factor |
| V | Proacclerin |
| VII | Proconvertin |
| VIII | Antihæmophilic Factor |
| IX | Christmas Factor |
| X | Stuart-Prower Factor |
| XI | Plasma thromboplastin antecedent |
| XII | Hageman Factor |

**Figure 2.**
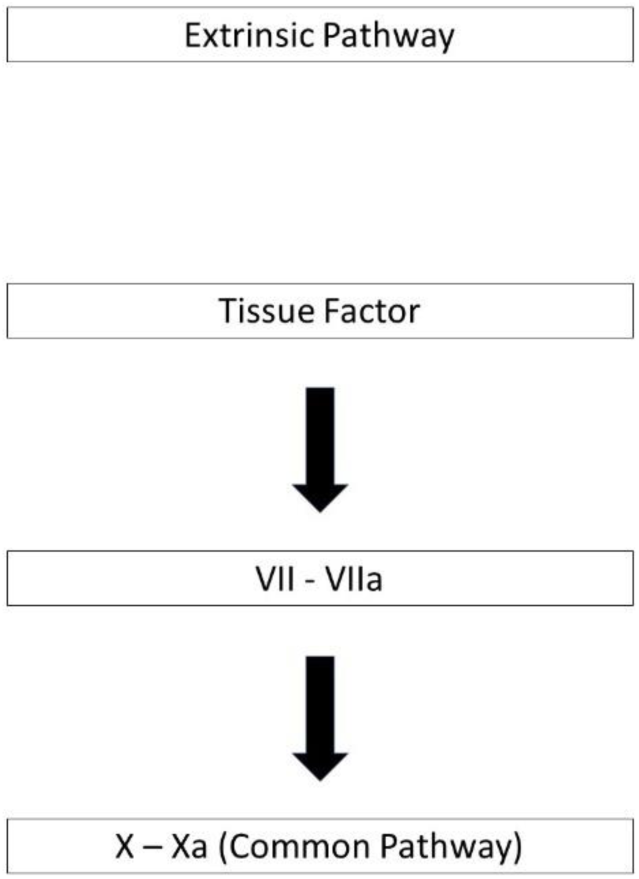
The Extrinsic Pathway.

**Figure 3.**
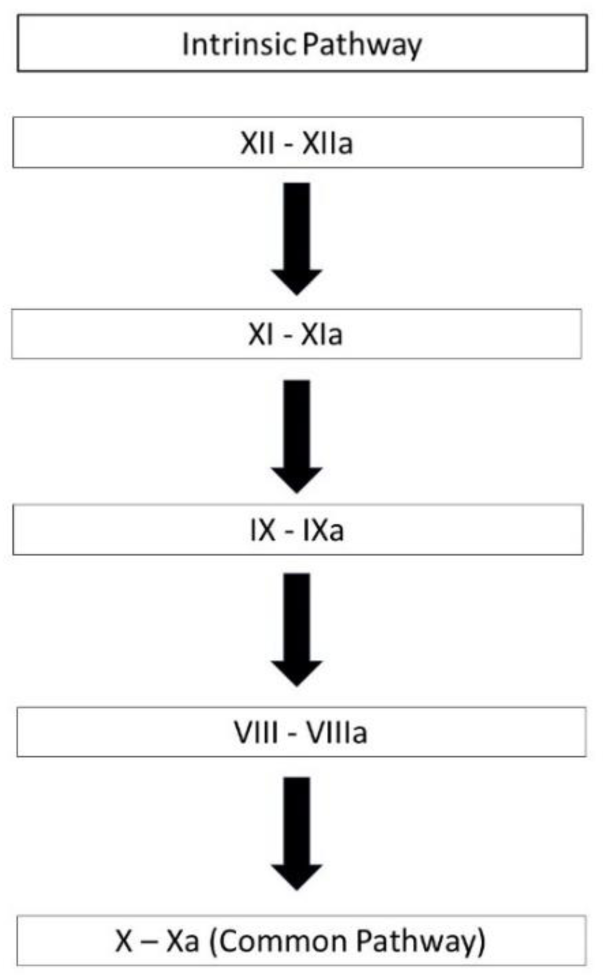
The Intrinsic Pathway.

**Figure 4.**
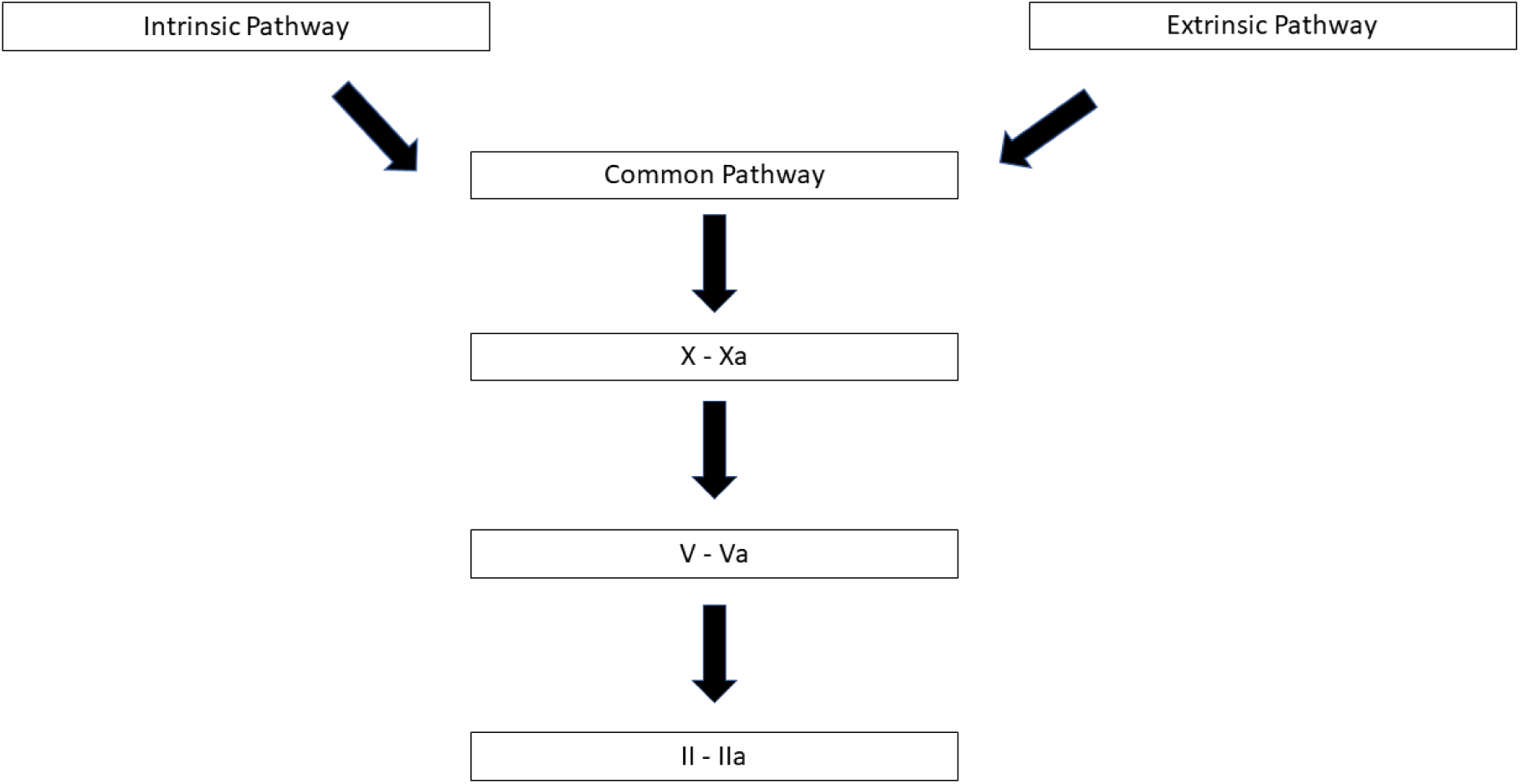
The Common Pathway.

The extrinsic pathway (see Figure 2) is initiated with the exposure of Factor VII to Tissue Factor. Tissue Factor is expressed on circulating particles released by monocytes and platelets and components of the vascular subendothelium including smooth muscle cells and fibroblasts (Konkle, 2017). Expression of Tissue Factor results from damage to the blood vessel walls. The exposure of Factor VII to Tissue Factor leads to activation of Factor VII (Factor VIIa). Factor VIIa then activates Factor X in the common pathway..

The intrinsic pathway (see Figure 3) is also known as the contact pathway (Smith et al, 2015). The intrinsic pathway is initiated *in vivo* by the inflammatory response. The intrinsic pathway *in vitro* can be initiated when the blood comes into contact with glass and other artificial surfaces. In the intrinsic pathway, Factor XII is activated to Factor XIIa. Factor XIIa then activates Factor XI to Factor XIa which in turn activates Factor IX to Factor IXa. Factor IXa then activates Factor VIII to Factor VIIIa.

The intrinsic and extrinsic pathways both act on the common pathway (see Figure 4) via activation of Factor X to Factor Xa. Factor Xa then activates Factor V to Factor Va. Finally Factor Va activates Prothrombin (formerly known as Factor II).

There is evidence to suggest that the intrinsic pathway’s role is to amplify the extrinsic pathway (Palta et al, 2014).

## Virchow’s Triad

Virchow’s triad is a well-established model of coagulation and provides a central framework for the interpretation of the data in this paper. Virchow’s triad is named after the 19^th^ century physician Dr Rudolph Virchow. Prior to Virchow there were different explanations for coagulation. Hippocrates advocated the model of the four humors (Yapijakis, 2009) which was further refined by Galen (Neder, 2020) and remained dominant for 1300 years. In this model, an imbalance in the four humors in the body accounted for disease. Organs had special functions. The spleen for instance was thought to function to remove black bile (Wilkins, 2002). Deep vein thrombosis was explained as a retention of humor (Galanaud et al, 2013). The model of the four humors eventually fell out of favour leaving the way open for new models.

Dr Richard Wiseman, royal physician to King Charles II, identified stasis and the characteristics of the blood as causes of thrombosis in a paper published in 1676 (Bagot and Arya, 2008) thus identifying two of the components of Virchow’s triad. In the late nineteenth century, Jean Cruvielhier, a French pathologist promoted an inflammatory theory of thrombosis. Virchow reacted against this theory and following experimental work, published his seminal work on thrombosis and emboli in 1856.

Virchow focused on pulmonary emboli and viewed them as originating in the deep veins. Rather than accounting for the process of thrombogenesis, Virchow identified a triad of effects resulting from the thrombus (Bagot and Arya, 2008) as well as explaining the propagation of the thrombus. Over the next one hundred and sixty-five years, the meaning of Virchow’s triad has been reworked and this interpretation is supported by accumulating evidence.

Virchow’s triad as it is now understood refers to three factors that predispose towards coagulation: Stasis of the blood, vessel wall damage and hypercoagulability. Whilst individual components of the triad are more easily investigated, the simultaneous *in vivo* characterisation of all three components is technically more challenging (Wolberg et al, 2012).

(Wolberg et al, 2012) differentiate arterial thrombi arising from ruptured atherosclerotic plaques and venous thrombi resulting from a combination of immobility, inflammation and hypercoagulability. There are numerous factors that lead to hypercoagulability and (Wolberg et al, 2012) describe a central role for Tissue Factor.

Blood flow is another key feature of Virchow’s triad. Laminar flow describes a streamlined flow which contrasts with turbulent flow in which some parts of a liquid will travel at a different velocity resulting in different characteristics of the overall flow. Atherosclerotic plaques create stenosis and can produce turbulent flow with high shear rates contrasting with low shear rates downstream of a plaque (Wolberg et al, 2012). The downstream area with low shear rates can predispose to thrombogenesis.

In areas of low shear (post-stenotic), viscosity increases and many factors predispose to thrombosis (Lowe, 2003). The white platelet-rich head of the arterial thrombi in the high-shear region is contrasted with the red tail of the thrombus in the low-shear region which is rich in red-cells.

In summary, Virchow’s triad involves a complex interplay between three factors that can each predispose to coagulation but together form a potent combination.

## Investigating COVID-19-Related Coagulopathies

Three possible aetiologies for a hypercoagulable state in Covid-19 are listed in Table 2 and were used in the initial examination of the literature in this paper.

**Table 2.** Pathologies Relevant to COVID-19 Related Coagulopathy.

|  |
| --- |
| Sepsis |
| ARDS |
| Disseminated Intravascular Coagulation |

## Sepsis

Sepsis is an important differential for COVID-19-related coagulopathy. The sepsis definition was revised in 2016 by a taskforce of the European Society of Intensive Care Medicine and the Society of Critical Care Medicine and the resulting definition was termed sepsis-3 (Singer et al, 2016). Sepsis-3 defines sepsis as a combination of infection with a dysregulated immune response and organ dysfunction. The construct of severe sepsis was removed in this definition and more severe pathology is described with the construct of septic shock which includes circulatory involvement (vasopressors are required to achieve a MAP >= 65mmHg) and cellular and metabolic abnormalities (with serum lactate levels > 2mmol/l). Previously the construct of the systemic inflammatory response syndrome (SIRS) was used in the definition of sepsis. In SIRS there is a normal immune response occurring with or without an infective aetiology (Abbas, 2018). The taskforce determined that SIRS did not have construct validity for sepsis.

Organ dysfunction in sepsis has been assessed using the SOFA score and in the sepsis-3 definition an increase of at least 2 points is used as a threshold for organ failure. The SOFA score has been specifically developed for the evaluation of organ failure in severe sepsis (Vincent et al, 1996). A number of practical drawbacks with the use of the SOFA score led to the development of the qSOFA score which is simpler to use in clinical practice (Singer et al, 2020) and also referenced in the sepsis-3 definition. (Giesen and Singer, 2018) suggest that organ dysfunction is functional rather than structural and may reflect an adaptive hibernation-like mechanism.

Sepsis commonly leads to coagulation with elevated D-Dimers, fibrinolysis and turnover of thrombin markers (Hunt, 2009). Clinically significant haemostatic changes have been identified in up to 70% of patients with sepsis (Levi, 2018). (Levi, 2018) describes a central role for cytokines in sepsis-related coagulopathy with IL-1, IL-6 and Tumour Necrosis Factor having a prominent role. (Levi, 2018) also notes that fibrinolysis may be downregulated and there is a reduction in the levels of Protein C/S, TFPI and antithrombin all of which may contribute to a coagulopathy. (Levi, 2018) also notes the important balance between inflammation and sepsis-related coagulopathy. (Iba et al, 2017) published their findings from the development of the sepsis-induced coagulopathy scoring system which has been widely used.

The importance of sepsis in COVID-19 has been recognised with the development of consensus guidelines for the management of sepsis in a critical care setting (Alhazzani et al, 2020). (Dumitrascu et al, 2020) provide an example of COVID-19-related coagulopathy with sepsis in a case where ophthalmic artery occlusion developed despite thromboprophylaxis.

In summary the definition of sepsis has been through several iterations, sepsis is generally associated with coagulopathy and the importance of managing sepsis in COVID-19 has been recognised.

## ARDS

Acute Respiratory Distress Syndrome (ARDS) is a syndrome of respiratory failure that is predominantly diagnosed and managed in an ICU setting. ARDS was first described in 1967 (Ashbaugh, 1967) and the definition has changed in response to practical and prognostic challenges (Moss and Thompson, 2009). The Berlin definition were published in 2012 (ARDS Definition Task Force, 2012) and include a threshold value for PaO2/FiO2, bilateral chest infiltrates evident on chest imaging, an absence of atrial hypertension or a threshold value for the pulmonary arterial pressure and an acute onset. ARDS was graded into mild, moderate and severe and requirements were made for PEEP or CPAP thresholds.

A number of treatment approaches have been developed for ARDS including prone positioning (Anzueto and Gattinoni, 2009), mechanical ventilation (Brower and Brochard, 2009), fluid therapy (Calfee et al, 2009), cell-based therapy (Gupta et al, 2009) and surfactant therapy (Spragg and Lewis, 2009) although some are experimental. The consensus guidelines for treatment of sepsis in a critical care setting includes the management of ARDS (Alhazzani et al, 2020).

(Mitchell, 2020) reviews the relationship of thromboinflammation to acute lung injury in COVID-19. There is noted to be damage to the endothelium and thrombosis in the perialveolar capillaries. (Mitchell, 2020) notes the antithrombotic and anti-inflammatory mechanisms of the vascular endothelium and that in COVID-19 there is loss of the contact with the basement membrane, exposing procoagulant factors. (Mitchell, 2020) also notes the influx of neutrophils and macrophages seen in COVID-19 in response to acute lung injury and their role in promoting thrombosis and inhibiting fibrinolysis. The Berlin definition does not include the construct of acute lung injury (ALI) due to inconsistencies in the use of the terminology (Fanelli et al, 2013) although this is referenced in the older literature and refers to a milder form of lung injury.

(Wheeler and Rice, 2009) outline a number of relationships that are relevant to the question of COVID-19-related coagulopathy. They note that up to 50% of cases of acute lung injury result from sepsis but also patients with acute lung injury may be more likely to develop sepsis. (Wheeler and Rice, 2009) also note that there is increased tissue factor expression, fibrin generation and impaired fibrinolysis sharing similarities to the pathology seen in sepsis. (Wheeler and Rice, 2009) note that in more severe cases of sepsis, the lung is usually involved and they suggest that this may be related to the extensive capillary network with exposure to the endothelial cells. The relationship between sepsis and ARDS is important not just in terms of coagulation but also for the risk of sepsis with prolonged ventilation (Valenza et al, 2009)

ARDS has been identified in COVID-19 but the utility of the syndrome has been questioned (Tobin, 2020). COVID-19-related ARDS may arise in multiple non-specialist settings which together with the silent hypoxia associated with COVID-19 has potential implications for the early recognition of ARDS in COVID-19. The association of ARDS with coagulopathy means that this is an important consideration in the aetiology of COVID-19-related coagulopathy.

## Disseminated Intravascular Coagulation

Disseminated intravascular coagulation (DIC) is an excessive activation of coagulation that can lead to significant mortality and morbidity. The 2001 definition by the International Society on Thrombosis and Haemostasis (ISTH) describes DIC as an acquired and generalised intravascular activation of coagulation (Taylor et al, 2001). DIC is associated with many diseases and conditions (see Table 3).

**Table 3.** Aetiology of Disseminated Intravascular Coagulation adapted from (McKay, 1968), (de Gopegui et al, 1995), (Venugopal, 2014)

|  |
| --- |
| A. Intravascular haemolysis |
| Haemolytic transfusion reactions |
| Haemolytic anaemia |
| B. Tissue injury |
| Massive tissue injury |
| Massive transfusion |
| Massive inflammation |
| Surgical procedures |
| Heat stroke |
| Severe trauma |
| Crush injuries |
| Massive burns |
| Severe hypo/hyperthermia |
| Severe pancreatitis |
| C. Infections |
| Bacterial infections |
| Gram negative (endotoxin) |
| Gram positive (bacterial coat mucopolysaccharide) |
| Viraemia (e.g. HIV, dengue) |
| Parasitic infections (e.g. malaria) |
| Fungal infection (e.g. invasive pulmonary Aspergillosis) |
| Protozoal infection |
| D. Obstetric complications |
| Fatty liver disease of pregnancy |
| Amniotic fluid embolism |
| Abruptio placentae |
| Pre-eclampsia |
| Retained products of placenta |
| E. Venoms/Toxins |
| Snake bite |
| Bee/insect sting |
| F. Malignancy |
| G. Miscellaneous |
| Diabetes mellitus |
| Vascular disorders - aortic aneurysms, giant haemangiomas (Kasabach-Merritt syndrome) |
| Anoxia |
| Graft versus host disease |
| Endothelial damage |
| Release of tissue thromboplastin |
| Proteolytic enzymes |
| Particulate or colloidal matter |
| Ingestion of certain lipid substances |

Activation of coagulation is recognised as a part of the host response to infection in sepsis (Okamoto, 2016). In DIC, the coagulation response is pathological and can result from the host response to infection. (Semeraro et al, 2014) describe the mechanisms of sepsis-related coagulopathy including disseminated intravascular coagulation. One of the mediators of the relationship between the host response to infection and coagulopathy is complement and (Kurosawa et al, 2014) review the complex relationship between complement and coagulation.

(Iba et al, 2019) distinguish between the microthrombosis that occurs predominantly in capillary venules in DIC and in arterioles in thrombotic microangiopathy. The distinction between thrombotic microangiopathy and DIC is also covered by (Wada et al, 2018). (Toh et al, 2017) note the importance of the multidisciplinary team in decision making with DIC as well as the nuances of interpretation of the haematological parameters. (Wada et al, 2014) distinguish four types of DIC according to their characteristics and recommend stratifying DIC according to these types when undertaking diagnosis and treatment. (Papageorgiou et al, 2018) review the treatments approaches for DIC.

There is an emerging evidence base for DIC in COVID-19. In a position statement by the Italian Haematology Society (Marietta et al, 2020), DIC was suggested as a cause of the hypercoagulable state in COVID-19. (Levi, 2020) argues that COVID-19-related coagulopathy is distinct from DIC and notes that the thrombocytopenia is not as profound as expected in DIC, that most patients with COVID-19 would not reach the criteria for overt DIC and that there isn’t evidence for excessive thrombin production. (Lillicrap, 2020) on the other hand cites the evidence that the criteria for overt DIC are more likely in non-survivors of COVID-19.

(Joob and Wiwanitkit, 2020) describe a case of COVID-19 with petechial rashes and low platelet count, initially diagnosed as Dengue fever. Dengue fever is also associated with DIC which in turn can result in petechial rashes. This case highlights the diagnostic challenges in COVID-19.

## Aim

The primary aim of this study was to determine if clotting pathology in COVID-19 occurs in the presence or absence of sepsis, disseminated intravascular coagulopathy and ARDS.

The secondary aims of this study were to use an iterative approach within the scoping review to:

1. Characterise the clotting pathology in COVID-19 with reference to the literature
2. To utilise the identified characteristics to develop a testable model.
3. To identify knowledge gaps
4. To make recommendations on research methodology based on the findings
5. To generate testable hypotheses in addition to those in the model.

## Methodology

The PRISMA extension for scoping reviews checklist was used for this paper (Tricco et al, 2018) and was a key framework for this paper. Additionally we developed a number of the methods used in this scoping review which we outline below.

### Overall Structure of Study

To test the hypothesis that there was a clotting mechanism independent of ARDS, sepsis and DIC we used a simple search strategy to identify the main papers. We described the quantitative findings and also applied a thematic analysis to identify themes both for the clinical findings in relation to the coagulopathy as well as the suggested explanatory mechanisms. We then utilised an iterative (Brunton et al, 2017) semi-structured search strategy to identify further papers relevant to the results of the thematic analysis for explanatory mechanisms. These papers were utilised in the development of a theoretical framework for the characterisation of the COVID-19-related coagulopathy.

### Protocol and Registration

There was no protocol due to the exploratory and iterative nature of the scoping review and no registration given the need to avoid a delay due to the COVID-19 pandemic.

### Rationale for Using a Mixed Methods Scoping Review

Whilst there is evidence to suggest that COVID-19 is associated with a coagulopathy, there are diverse views on this coagulopathy and a number of potential mediators as above. COVID-19 has only recently been described and the evidence base is developing. Given the above, a scoping review was selected in favour of a meta-analysis or systematic review where the research questions are clearer and the research evidence base may be more well-developed. A mixed methods approach was used to maximise the yield from the identified studies.

### Literature Search for Papers with Evidence of Clotting

As this is a scoping review, broad search terms were used to identify evidence of clinically significant clotting disorders. The search terms used in the Pubmed database (Pubmed, 2020) are shown in Table 4.

**Table 4.**
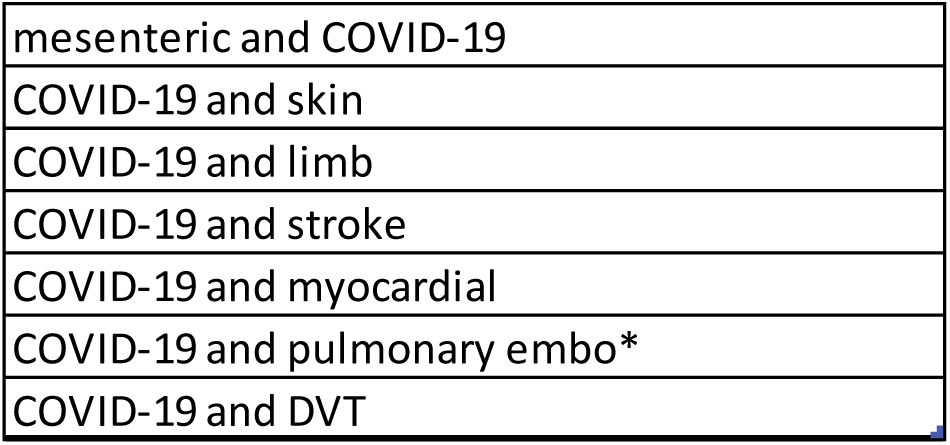
Pubmed Search Terms.

|  |
| --- |
| mesenteric and COVID-19 |
| COVID-19 and skin |
| COVID-19 and limb |
| COVID-19 and stroke |
| COVID-19 and myocardial |
| COVID-19 and pulmonary embo* |
| COVID-19 and DVT |

The inclusion and exclusion criteria are shown in Table 5. The COVID-19 initiative has made COVID-19 related papers available during the pandemic (GPMB, 2020). We therefore included only papers that were freely available including through the COVID-19 initiative as this review did not receive any external funding.

**Table 5.**
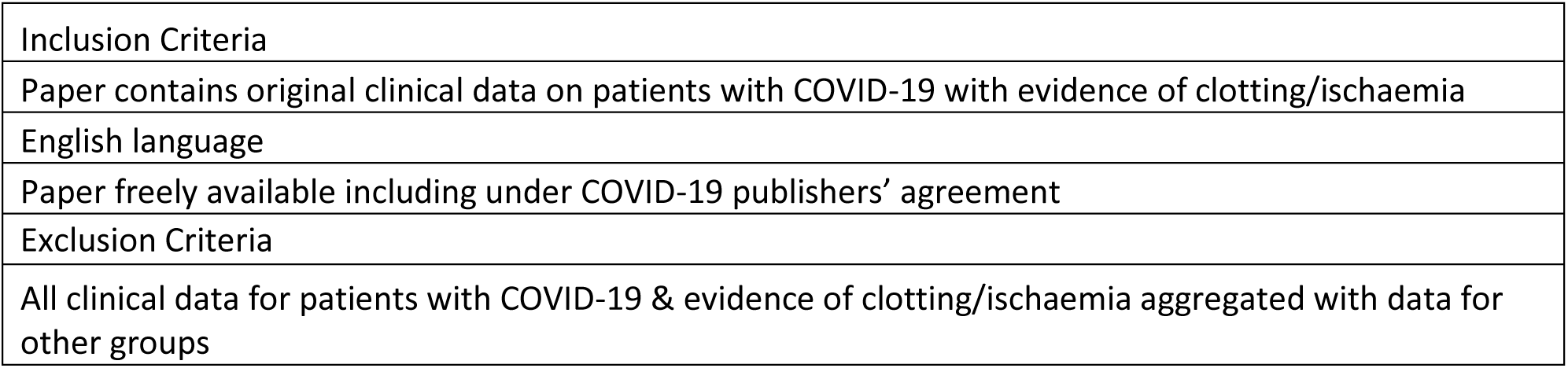
Inclusion and Exclusion Criteria.

|  |
| --- |
| Inclusion Criteria |
| Paper contains original clinical data on patients with COVID-19 with evidence of clotting/ischaemia |
| English language |
| Paper freely available including under COVID-19 publishers' agreement |
| Exclusion Criteria |
| All clinical data for patients with COVID-19 & evidence of clotting/ischaemia aggregated with data for other groups |

The abstracts were evaluated and after exclusions, the remaining papers were examined in detail and further exclusions took place. We excluded papers that did not present any of the data for patients with COVID-19 separately from patients without COVID-19 as this was needed for the analysis.

### Analysis of Main Papers

#### Software

We used Microsoft Excel® for Windows 365 to store and analyse the data.

We used Microsoft PowerPoint for the Diagrammatic Mapping.

#### Data Analysis

A template was created using an Excel spreadsheet and the data was filled on analysing the papers. Several columns were used to describe the pathology where more than one pathology existed in the same patient. We utilised reports of pathology, descriptions of imaging findings, operative findings, autopsy and biopsy evidence. In cases where there was clear evidence of end-organ ischaemia, we counted this as an thromboembolic event and have grouped this together with clotting episodes where thromboembolism was identified or inferred from the evidence (e.g. loss of patency of a blood vessel on imaging).

For each of the studies we collected information on the number of cases, individual thromboembolic complications, sex, mortality, number of non-COVID-19 or non-thromboembolic cases and whether information was recorded on D-Dimers, fibrinogen, platelet count, prothrombin time, PaO2/FiO2, bilateral chest infiltrates evident on chest imaging, an absence of atrial hypertension or a threshold value for the pulmonary arterial pressure, acuteness of onset, confirmation of ARDS, sepsis, DIC, antithrombin, Protein C, whether respiratory rate or blood pressure were recorded, whether qSofa or SOFA score were recorded, mental status, main country of authors. We looked for evidence of randomisation or blinding as well as a power analysis.

Whilst the analysis was underway we identified a further potential mediator of the COVID-19-related coagulopathy - activation of the alternative complement pathway (Magro et al, 2020) and added this to the analysis of the 56 papers. Where clarification was needed, we contacted the authors of the papers.

A subset of 34 papers was identified in which individual patient data was available including sex, age and pathology. This enabled us to look at sex as a biological variable. The data was aggregated and analysed, further separated according to outcome and sex and comparisons between groups based was undertaken. We utilised a two proportion Z-test to determine the statistical significance of the difference in proportions between various sample proportions.

We used the Benjamini and Hochberg procedure to correct for multiple comparisons using a false discovery rate of 0.25 (McDonald, 2014)(Benjamini and Hochberg, 1995).

#### Abridged Thematic Analysis and Validated Abridged Thematic Analysis

A qualitative analysis was undertaken, using what we refer to as an abridged thematic analysis which we extended by adding(see Figure 5). The 56 main papers are classed as a secondary source for the purposes of thematic analysis (Braun and Clarke, 2006).

**Figure 5.**
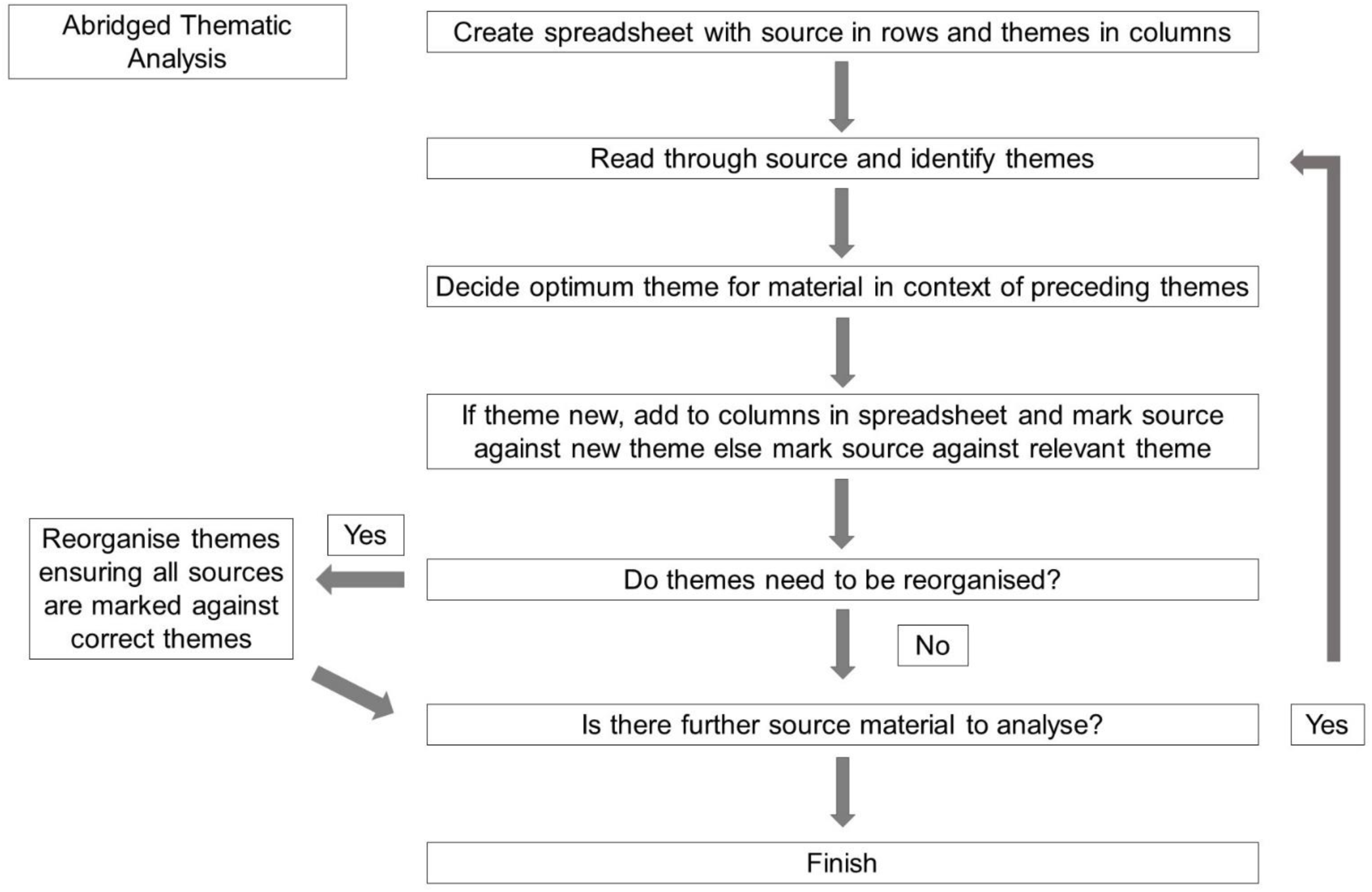
Outline of Abridged Thematic Analysis Procedure.

We abridged the thematic analysis by removing the coding process and working out the themes as we moved through the papers. We have outlined the process in figure 6 to enable this to be reproduced.

**Figure 6.**
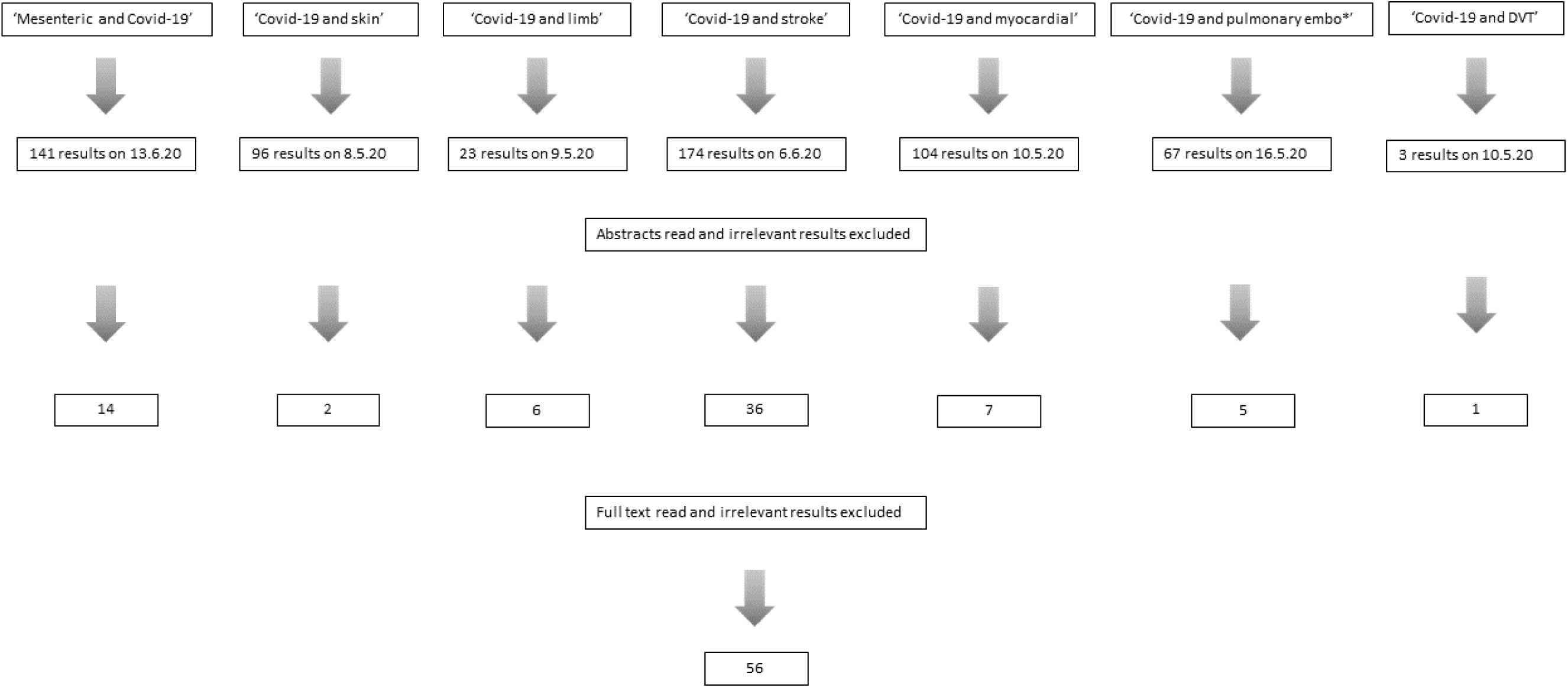
illustrating the process of selection of papers for final analysis.

The application of the abridged thematic analysis was two-fold.

1. Firstly we looked for any characterisation of the coagulopathy. We identified any distinct characteristics reported by the authors based on their clinical findings and determined if they qualified as a theme and if so which theme. When we had identified all of the themes, we then quantified the frequency of the theme in the papers.
2. Secondly we looked for explanatory mechanisms for the COVID-19-related coagulopathy. The authors were clinicians providing a specialised expert perspective on their clinical experience with patients with COVID-19 and bringing their expert knowledge to bear on the question of aetiology. Again we identified the themes and organised them into an initial structure. We then undertook a validation process utilising an exploratory literature search (detailed below). We then analysed the identified papers and determined if there was sufficient evidence to confirm or refute the identified aetiological mechanisms. After this subsequent analysis, we were able to remove certain aetiologies and restructure the taxonomy of the aetiologies. We referred to this extended process as validated abridged thematic analysis (VATA).

#### Exploratory Literature Search

The exploratory literature search involved using specified search terms in Pubmed and selecting English language articles that were freely available including under the COVID-19 agreement and which were predominantly meta-analyses or systematic reviews in order to gain a rapid overview of the subject. We used these together with case studies or case series. We also used ‘forward searching’ from within the citations index of identified papers (Petticrew and Roberts, 2006) as well as personal knowledge of papers we were already familiar with (Grewal et al, 2020). For more selective questions we used additional resources including MedRxiv (RRID:SCR_018222), bioRxiv (RRID:SCR_003933), DOAJ - Directory of Open Access Journals (RRID:SCR_004521) and AMEDEO: The Medical Literature Guide (RRID:SCR_002284). We thus used a combination of primary, secondary and tertiary literature (Grewal et al, 2020). We have included search terms used in the exploratory literature search as part of the VATA as open data in the supplemental data.

#### Indexing the Results from the Validated Abridged Thematic Analysis

After validating the identified themes against the clinical literature, we then reorganised the themes and documented a justification for the final taxonomy of themes. We used alphanumeric identifiers to label the themes. We also utilised the resulting taxonomy to generate a narrative summary which is presented at the end of the paper.

#### Diagram Mapping

We mapped the final taxonomy of themes onto corresponding diagrams using the alphanumeric identifiers and a set of rules which we outline below.

1. Each diagram is labelled with an alphanumeric identifier which maps onto the taxonomy
2. Concepts/constructs/phenomenon are represented by boxes with text descriptions
3. The relationships between concepts/constructs/phenomenon are identified by arrows
4. The arrows are colour coded according to the strength of evidence for the relationship and whether the evidence exists in the COVID-19 literature or in the general (i.e. non-COVID-19) literature.
5. The arrows encompass a broad range of logical relationships resulting from deductive, inductive and abductive reasoning based upon the evidence.
6. The relationships encompass multiple ontological levels.

## Results

### Results of Main Literature Search

The initial results of the literature search are shown in Figure 6 and includes the dates of the searches. The searches identified a total of 608 papers which were reduced to 71 papers after a review of the abstracts and finally 56 papers after inspection of the full text and application of the inclusion/exclusion criteria. A number of the papers reported on mixed clotting/ischaemic pathologies (e.g. stroke and lower limb deep vein thrombosis) and so the final 56 papers are pooled. The final papers are listed in Table 6.

**Table 6.**
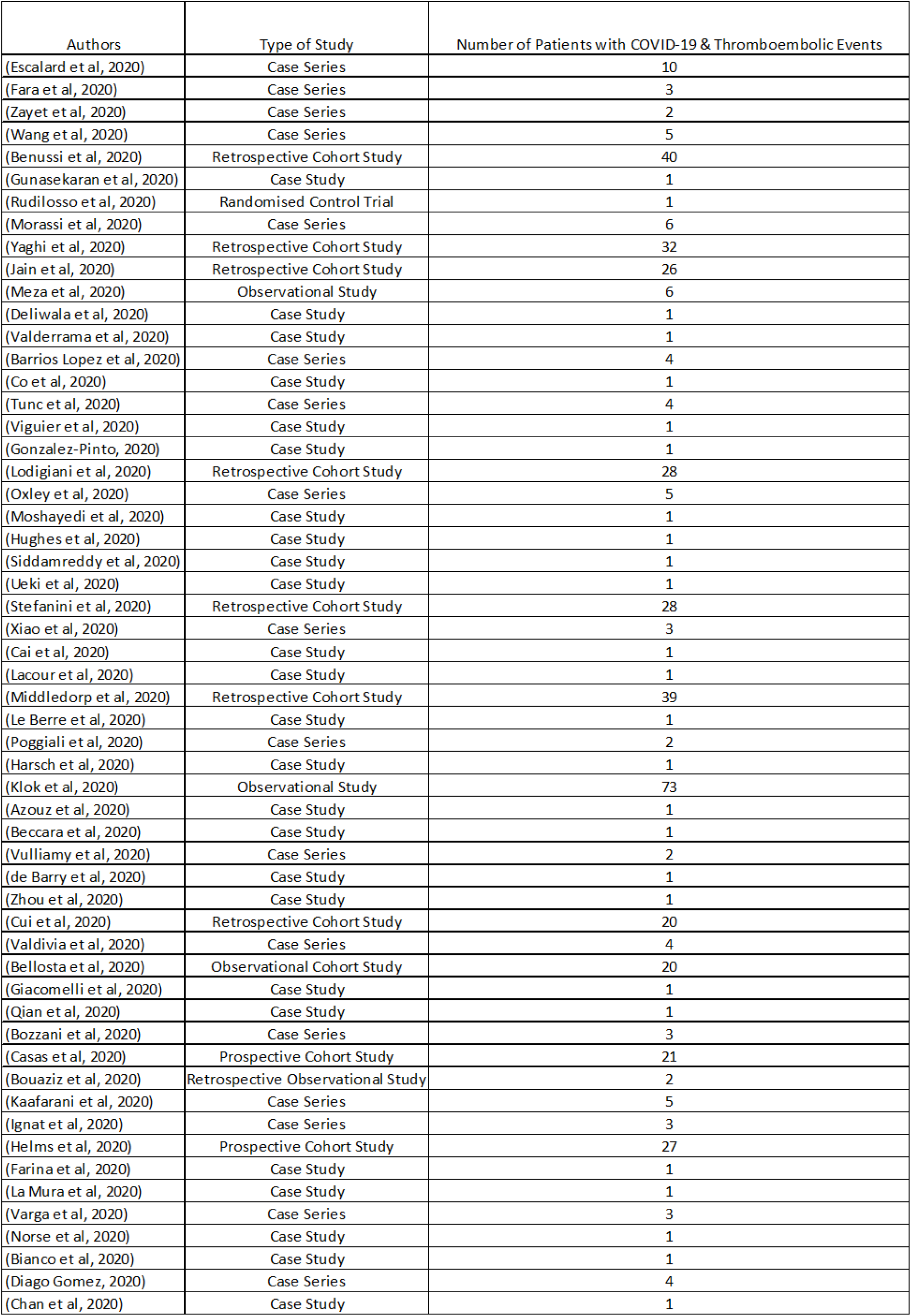
Summary of Identified Studies.

### Country of Publication of Included Papers

The countries of publication are shown in Figure 7. There were no authors based in Africa, Oceania or South America. The authors of 84% of the papers included in the analysis were based in five countries (Italy, USA, France, Spain and China).

**Figure 7.**
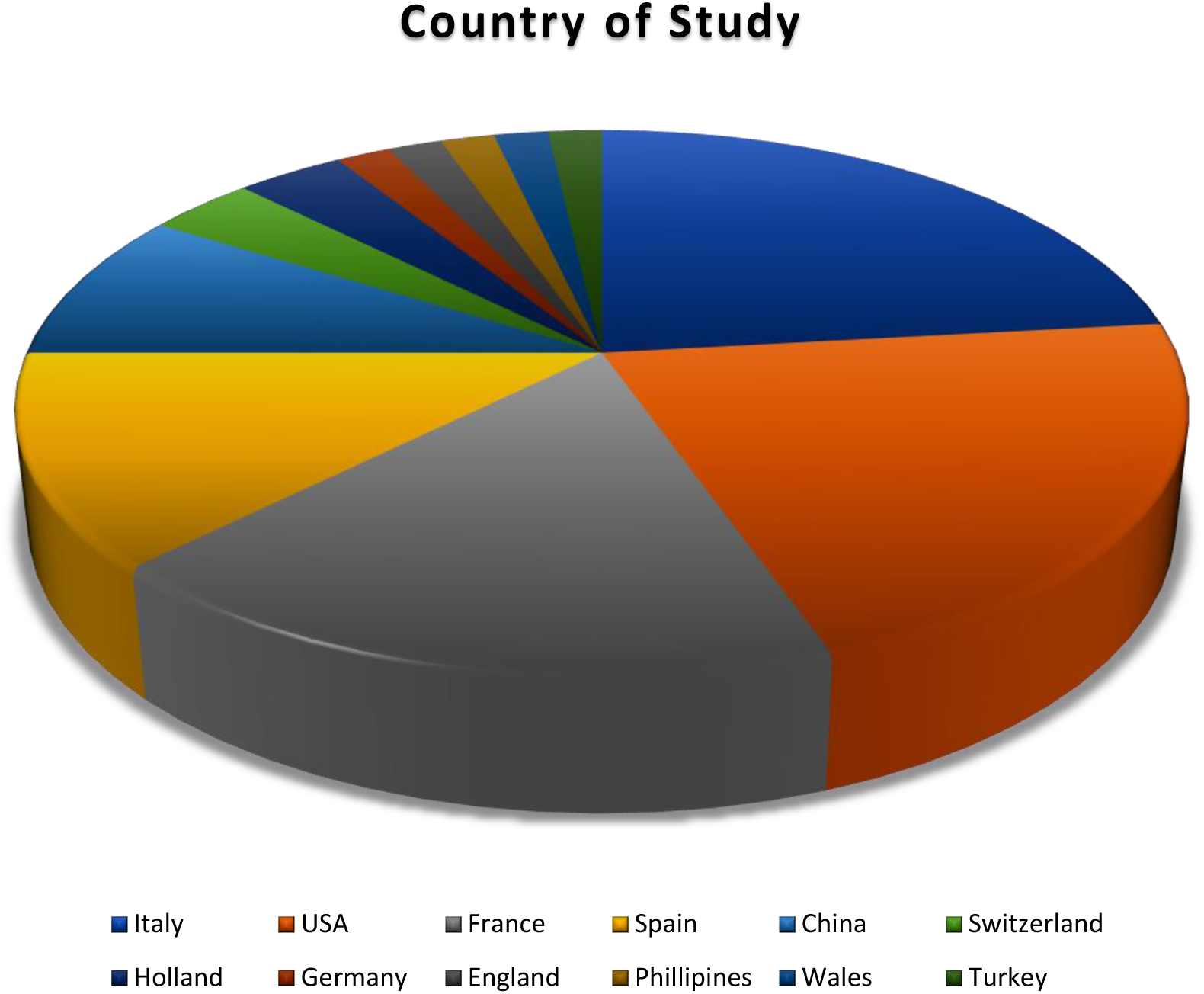
Country of Publication.

### Thromboembolic/Ischaemic Events in 56 Papers

The descriptive statistics for thromboembolic/ischaemic events in the 56 papers are shown in Table 7.

**Table 7.** Descriptive Statistics for Thromboembolic/Ischaemic Events in the 56 Papers.

|  |  |
| --- | --- |
| Total Number of Patients in All Studies | 10523 |
| Total Number of Patients with COVID-19 and Thromboembolic/Ischaemic Events | 456 |
| Total Number of Thromboembolic/Ischaemic Events | 586 |
| Average Number of Thromboembolic/Ischaemic Events Per Patient | 1.3 (1 d.p.) |

The analysis shows that just over 4% of the total number of patients in all the studies had a combination of COVID-19 and thromboembolic/ischaemic events. Furthermore the average number of thromboembolic/ischaemic events was above 1.

The distribution of the average number of ischaemic/clotting events per patient is illustrated in Figure 8. The spread of the data was not normally distributed but instead was skewed towards the mode which was one thromboembolic/ischaemic event per patient.

**Figure 8.**
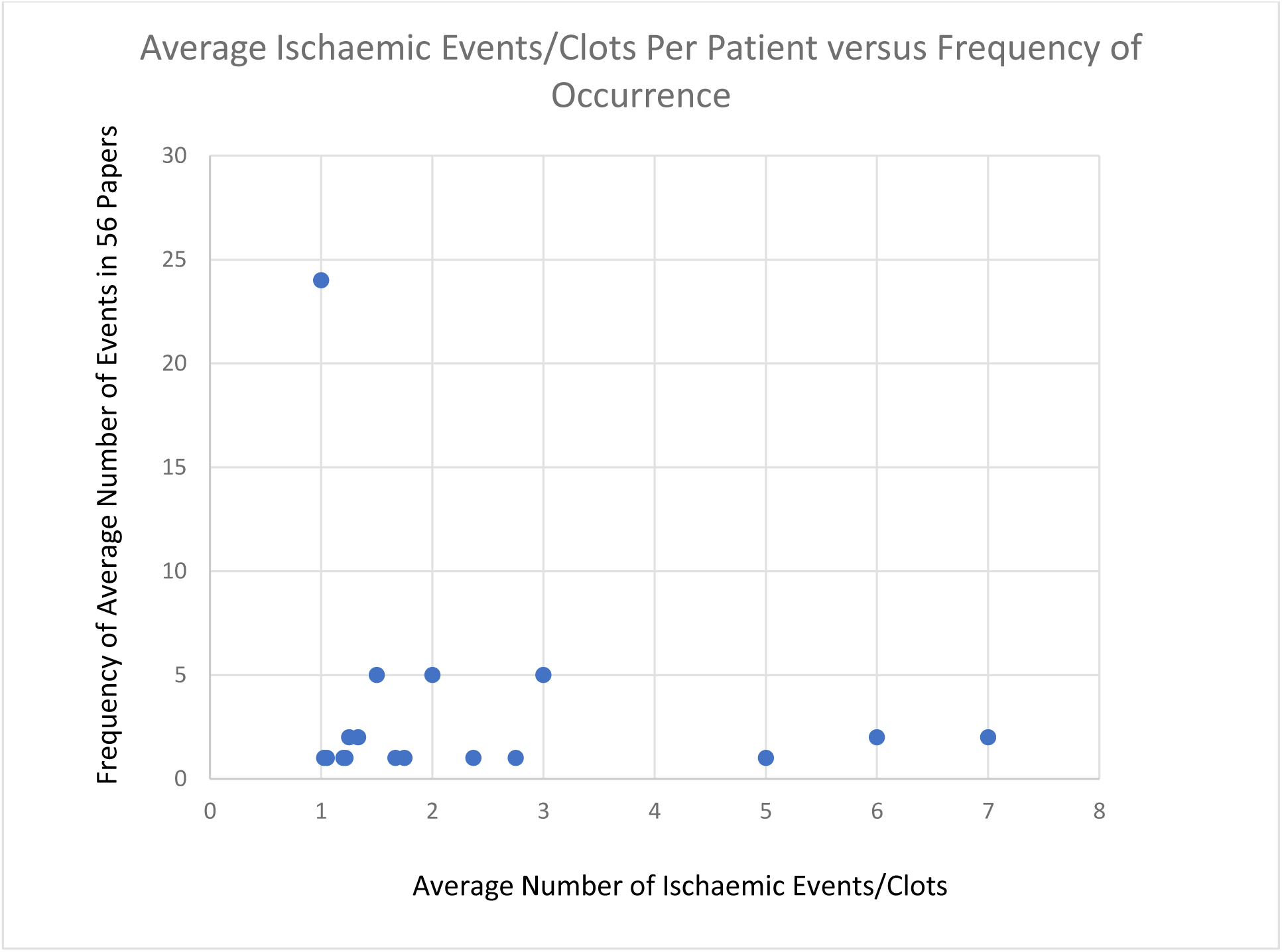
Average Number of Clots/Ischaemic Events Per Patient v Frequency of Occurrence.

The number of thromboembolic/ischaemic events is categorised in Table 8. Strokes are differentiated according to the description. Where the occluded artery/arteries are not identified, the pathology has been identified through the infarcted regions. Several infarcted regions may result from the occlusion of a single artery but the location is not predictable due to anatomical variants and collateral circulation and therefore in these cases the distinct regions are counted. Skin livedo and necrosis are counted as thromboembolic/ischaemic events although data was not available in these cases on the potentially relevant arterial patency (e.g. acute limb ischaemia). Upper limb DVT was differentiated according to the absence or presence of a catheter with the latter group being included with medical device-associated clotting.

**Table 8.**
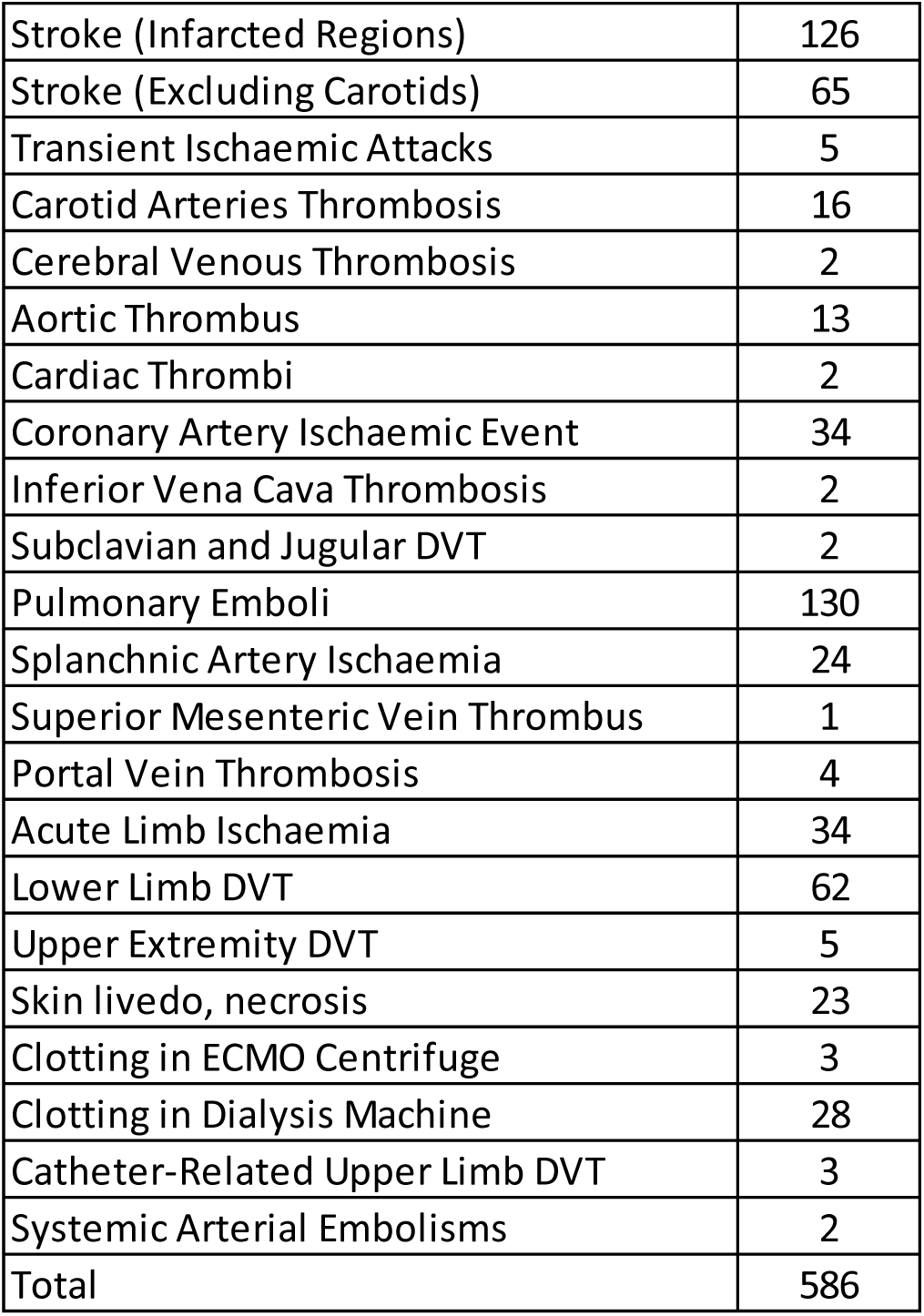
Thromboembolic/Ischaemic/Clotting Events in 56 main papers.

Table 9 shows the number and percentage of papers reporting a range of syndromes, diagnoses and blood test results relevant to COVID-19-related coagulopathy. The D-Dimers and platelet count were the two most frequently reported parameters in the 56 papers.

**Table 9.** Number (%) of Papers Reporting Clinical Variables/Diagnoses/Syndromes for 56 Main Papers.

|  | Number of Papers | % of Total Papers (2 s.f.) |
| --- | --- | --- |
| Platelet | 22 | 39 |
| PT Prolongation | 18 | 32 |
| Fibrinogen | 15 | 27 |
| D-Dimer | 35 | 63 |
| Antithrombin | 4 | 7 |
| Protein C | 3 | 5 |
| DIC confirmation | 4 | 7 |
| ARDS confirmation | 7 | 13 |
| Sepsis confirmation | 3 | 5.4 |
| AH50 | 0 | 0 |

### ARDS

Out of the fifty-six main papers, seven studies provided evidence of ARDS with a total of 159 patients with COVID-19 and ARDS. With the exception of (Helms et al, 2020) these were all case studies or case series and are shown in Table 10. (Helms et al, 2020) reported 150 patients with 64 thrombotic complications and without evidence of disseminated intravascular coagulation. The authors also compared the patients with COVID-19 and ARDS to a group with ARDS but without COVID-19. They reported a significantly increased likelihood of thrombotic events in the patients with COVID-19 (Odds Ratio 2.6 [1.1–6.1], p=0.035). The SOFA scores did not differ between the two groups.

**Table 10.** Papers with ARDS Confirmation.

|  |  |
| --- | --- |
| (Deliwala et al, 2020) | Single case with stroke |
| (Siddamreddy et al, 2020) | Single case with inferior wall STEMI |
| (Ueki et al, 2020) | Single case with inferoposterior STEMI |
| (Bellosta et al, 2020) | 4 fatal cases with limb ischaemia |
| (Qian et al, 2020) | Single case with upper and lower limb ischaemia |
| (Ignat et al, 2020) | Single case of bowel ischaemia with ARDS |
| (Helms et al, 2020) | 150 patients with ARDS with 64 thrombotic events |

### Sepsis

In the 56 main papers, two of the papers provide evidence of sepsis and are shown in Table 11. (Norse et al, 2020) report the case of a 62-year-old man who developed mesenteric ischaemia requiring bowel surgery and later died of septic shock. In another paper the sepsis results from an E.Coli infection comorbid with COVID-19 and there is mesenteric ischaemia (La Mura et al, 2020).

**Table 11.** Papers Confirming Sepsis.

|  |  |
| --- | --- |
| (La Mura et al, 2020) | E.Coli sepsis with mesenteric ischaemia |
| (Norse et al, 2020) | Case study with mesenteric ischaemia |

There are other papers where sepsis is discussed but the confirmation is less clear. (Barrios Lopez et al, 2020) report on 4 cases of ischaemic stroke with COVID-19 and suggest septic shock as a cause.

Other papers mention the use of the SOFA or qSOFA score which are intended for use in sepsis although they have been used for critically ill patients more generally and are shown in Table 12.

**Table 12.** Papers with SOFA/qSOFA/SIC score.

|  |  |
| --- | --- |
| (Benussi et al, 2020) | 35 patients with ischaemic stroke, higher qSOFA on admission with COVID-19 |
| (Kaafarani et al, 2020) | 141 patients with COVID-19, SOFA score aggregated, subset of patients with mesenteric ischaemia |
| (Helms et al, 2020) | 150 patients with COVID-19 and 64 thrombotic complications, SOFA scores aggregated |
| (La Mura et al, 2020) | E.Coli sepsis with mesenteric ischaemia |
| (Poggiali et al, 2020) | Case study with elevated SIC score and DVT |

(La Mura et al, 2020) mentions both the diagnosis and the SOFA score. (Poggiali et al, 2020) present two cases of COVID-19 who present with venous thromboembolism. They present their findings with the Sepsis-Induced Coagulopathy (SIC) score as evidence for this pathology. The main focus of (Helms et al, 2020) is on ARDS although they exclude DIC (depending on the scoring system) and have included aggregated SOFA scores.

### Disseminated Intravascular Coagulation

DIC was specifically confirmed or excluded in four papers which are shown in Table 13.

**Table 13.** Papers Confirming or Excluding DIC.

|  |  |
| --- | --- |
| (Klok et al, 2020b) | DIC excluded in all cases |
| (Lodigiani et al, 2020) | 8 patients with overt DIC, 2 (25%) with thromboembolic events, 7 (88%) died |
| (Validivia et al, 2020) | 1 case of acute limb ischaemia with DIC and death |
| (Helms et al, 2020) | 0-6 patients with DIC depending on scoring system |

(Helms et al, 2020) have used different scoring systems to assess DIC in patients and identified between 0 and 6 patients with DIC depending on the scoring system.

(Klok et al, 2020b) excluded DIC in all cases while (Lodigiani et al, 2020) identified eight patients with overt DIC. (Klok et al, 2020b) excluded DIC in all cases. (Lodigiani et al, 2020) identified 8 patients with DIC, with 25% of patients manifesting thromboembolic events and 88% of patients dying underlining the important consequences of DIC. (Valdivia et al, 2020) report a single case of acute limb ischaemia with DIC and death. (Helms et al, 2020) investigated DIC as a secondary outcome and used various scoring methods to ascertain caseness. No patients were identified with DIC using the ISTH “overt” score, 6 cases of DIC were identified using the JAAM-DIC score and 22 patients were identified using the SIC score indicating those at risk of DIC. Thus there was evidence of thromboembolic complications in patients with COVID-19 and where DIC had both been confirmed or excluded.

### A Subset of 34 Studies with Demographic Data

We identified a subset of 34 studies which contained individual information on the age and gender of the patient as well as the corresponding pathology, enabling a more detailed characterisation of the pathology. The 34 studies are listed in Table 14.

**Table 14.** 34 Papers with Individual Clinical Information on Pathology, Gender and Mortality.

|  |  |  |
| --- | --- | --- |
| (Fara et al, 2020) | (Deliwala et al, 2020) | (Valderrama et al, 2020) |
| (Barrios Lopez et al, 2020) | (Co et al, 2020) | (Viguier et al, 2020) |
| (Gonzalez-Pinto, 2020) | (Lodigiani et al, 2020) | (Oxley et al, 2020) |
| (Hughes et al, 2020) | (Siddamreddy et al, 2020) | (Ueki et al, 2020) |
| (Stefanini et al, 2020) | (Xiao et al, 2020) | (Cai et al, 2020) |
| (Lacour et al, 2020) | (Le Berre et al, 2020) | (Poggiali et al, 2020) |
| (Harsch et al, 2020) | (Beccara et al, 2020) | (Vulliamy et al, 2020) |
| (de Barry et al, 2020) | (Zhou et al, 2020) | (Cui et al, 2020) |
| (Bellosta et al, 2020) | (Giacomelli et al, 2020) | (Ignat et al, 2020) |
| (Farina et al, 2020) | (La Mura et al, 2020) | (Varga et al, 2020) |
| (Norse et al, 2020) | (Bianco et al, 2020) | (Diago Gomez, 2020) |
| (Diago Gomez, 2020) |  |  |

In the 34 studies, the distribution of thromboembolic/ischaemic events per patient was similar to Figure 7, being skewed towards the mode value of 1 thromboembolic/ischaemic event per patient with a range of 1-8. The results are shown in Tables 8-12. In the limb ischaemia group, eight of the patients had been reported from one study where the data had been published selectively for patients that had died (i.e. individual data was not available for patients that had not died).

The 34 papers included 119 patients with COVID-19 and thromboembolic/ischaemic events and the overall results are shown in Table 15. All of the studies are based in a hospital setting. There are a number of patients with multiple thromboembolic/ischaemic events but the numbers are counted for each type of event and the sum is therefore greater than the number of patients. 38% of the patients in this group died. The percentage of deaths varied from just under 8% in those with stroke-related thromboembolic events to just under 77% in those with acute limb ischaemia.

**Table 15.** Results from Analysis of Cases from 34 Papers with Individual Data.

|  |  |
| --- | --- |
| Number of Patients | 119 |
| Average Age of Patients | 67 (2 s.f.) |
| Number of Male Patients | 82 |
| Number of Female Patients | 37 |
| Number of Deaths (%) | 45 (38% (2 s.f.)) |
| Number of Thromboembolic/Ischaemic Events | 205 |
| Average Number of Events Per Patient | 1.72 (2 d.p.) |
| Range of Number of Events Per Patient | (1-8) |
| Mode of Number of Events Per Patient | 1 |
| Number of Patients with Aortic Thromboembolic Events (%) | 9 (7.6 (1 d.p.)) |
| Deaths in Patients with Aortic Thromboembolic Event (%) | 3 (33%) |
| Number of Patients with Stroke Related Events Including Carotid Arteries (%) | 33 (27.7%(1 d.p.)) |
| Deaths in Patients with Stroke Related Events (%) | 9 (7.6%(1 d.p.)) |
| Number of Patients with Cardiac Thromboembolic/Ischaemic Events (%) | 42 (35%) |
| Deaths in Patients with Cardiac Thromboembolic/Ischaemic Events (%) | 15 (36% (2 s.f.)) |
| Number of Patients with Pulmonary Emboli (%) | 16 (13% (2s.f.)) |
| Deaths in Patients with Pulmonary Emboli (%) | 4 (25%) |
| Number of Patients with Venous Thromboembolic Events excluding PE (%) | 19 (16% (2 s.f.)) |
| Deaths in Patients with Venous Thromboembolic Events excluding PE (%) | 5 (26%(2 s.f.)) |
| Number of Patients with Thromboembolic Events in Splanchnic Arteries (%) | 15 (12.6%(1 d.p.)) |
| Deaths in Patients with Thromboembolic Events in Splanchnic Arteries (%) | 9 (60%) |
| Number of Patients with Acute Limb Ischaemic Events (%) | 14 (12%(2 s.f.)) |
| Deaths in Patients with Acute Limb Ischaemia (%) | 11 (78.6%(1 d.p.)) |

The results from the analysis of the female cases in the 34 papers is shown in Table 16. 32% of the patients in this group died. The deaths associated with each type of thromboembolic/ischaemic event ranged from 21% with cardiac thromboembolic/ischaemic events to 100% with acute limb ischaemia. The results from the analysis of the male cases in the 34 papers is shown in Table 17. There were just over twice as many males as females in the 34 papers and 40% of the patients in this group died. There were eight deaths reported in one of the studies where the data was not available for those who survived. The deaths associated with each type of thromboembolic/ischaemic event ranged from 18% with pulmonary emboli to 75% with acute limb ischaemia.

**Table 16.** Analysis of Female Cases in 34 Papers with Individual Data.

|  |  |
| --- | --- |
| Number of Patients | 37 |
| Average Age of Patients | 66 |
| Number of Deaths (%) | 12 (32% 2 s.f.) |
| Number of Thromboembolic/Ischaemic Events | 55 |
| Average Number of Events Per Patient | 1.48 (2 d.p.) |
| Range of Number of Events Per Patient | (1-6) |
| Mode of Number of Events Per Patient | 1 |
| Number of Patients with Aortic Thromboembolic Events (%) | 2 (5.4% (1 d.p.)) |
| Deaths in Patients with Aortic Thromboembolic Event (%) | 1 (50%) |
| Number of Patients with Stroke Related Events Including Carotid Arteries (%) | 12 (32%) |
| Deaths in Patients with Stroke Related Events (%) | 4 (33%) |
| Number of Patients with Cardiac Thromboembolic/Ischaemic Events (%) | 14 (38% 2 s.f.) |
| Deaths in Patients with Cardiac Thromboembolic/Ischaemic Events (%) | 3 (21% (2 s.f.)) |
| Number of Patients with Pulmonary Emboli (%) | 5 (14% 2 s.f.) |
| Deaths in Patients with Pulmonary Emboli (%) | 2 (40)% |
| Number of Patients with Venous Thromboembolic Events Excluding PE (%) | 7 (19% 2 s.f.) |
| Deaths in Patients with Venous Thromboembolic Events (%) | 2 (28.6% (1 d.p.)) |
| Number of Patients with Thromboembolic Events in Splanchnic Arteries (%) | 3 (8% 1 s.f.) |
| Deaths in Patients with Thromboembolic Events in Splanchnic Arteries (%) | 2 (67% 2 s.f.) |
| Number of Patients with Acute Limb Ischaemic Events (%) | 2 (3.6% (1 d.p.)) |
| Deaths in Patients with Acute Limb Ischaemia (%) | 2 (100%) |

**Table 17.** Analysis of Male Cases in 34 Papers with Individual Data.

|  |  |
| --- | --- |
| Number of Patients | 82 |
| Average Age of Patients | 67 |
| Number of Deaths (%) | 33 (40% 2 s.f.) |
| Number of Thromboembolic/Ischaemic Events | 150 |
| Average Number of Events Per Patient | 1.83 (2 d.p.) |
| Range of Number of Events Per Patient | (1-8) |
| Mode of Number of Events Per Patient | 1 |
| Number of Patients with Aortic Thromboembolic Events (%) | 7 (8.54% 2 d.p.) |
| Deaths in Patients with Aortic Thromboembolic Event (%) | 2 (28.6% (1 d.p.)) |
| Number of Patients with Stroke Related Events Including Carotid Arteries (%) | 21 (25.6%(1dp)) |
| Deaths in Patients with Stroke Related Events (%) | 5 (23.8% (1 d.p.)) |
| Number of Patients with Cardiac Thromboembolic/Ischaemic Events (%) | 28 (34% (2 s.f.)) |
| Deaths in Patients with Cardiac Thromboembolic/Ischaemic Events (%) | 12 (42.9% 1 d.p.) |
| Number of Patients with Pulmonary Emboli (%) | 11 (13%(2 s.f.)) |
| Deaths in Patients with Pulmonary Emboli (%) | 2 (18% (2 s.f.)) |
| Number of Patients with Venous Thromboembolic Events Excluding PE (%) | 12 (14.6%(1 d.p.)) |
| Deaths in Patients with Venous Thromboembolic Events (%) | 3 (25%) |
| Number of Patients with Thromboembolic Events in Splanchnic Arteries (%) | 12 (14.6%(1 d.p.)) |
| Deaths in Patients with Thromboembolic Events in Splanchnic Arteries (%) | 7 (58% (2 s.f.)) |
| Acute Limb Ischaemic Events (%) | 12 (14.6%(1 d.p.)) |
| Deaths in Patients with Acute Limb Ischaemia (%) | 9 (75%) |

We also compared the results in the 34 papers for those who died and those who survived. The results for the cases of those who died are shown in Table 18 where the average age is 73. The four main types of thromboembolic/ischaemic events in this group in increasing percentages were thromboembolic/ischaemic events in the splanchnic arteries (20%), stroke-related events including the carotid arteries (24%), acute limb ischaemia (24%) and cardiac thromboembolic/ischaemic events (33%).

**Table 18:** Calculating Difference between proportions using Z-test: F – female, M – male, FP – female proportion of sample, MP – male proportion of sample, F PC – female proportion of cases, M PC – male proportion of cases, D – difference between proportions (smaller value subtracted from larger), SP – sample proportion for test statistic calculation, SE – standard error, TS – test statistic, p – p-value

|  | F | M | F P | MP | F PC | M PC | D | SP | SE | TS | p |
| --- | --- | --- | --- | --- | --- | --- | --- | --- | --- | --- | --- |
| Number of Deaths | 12 | 33 | 0.3243 | 0.4024 |  |  | 0.078115 | 0.378151 | 0.096038 | 0.813377 | 0.20897 |
| Number of cases of aortic thrombembolism | 2 | 7 | 0.0541 | 0.0854 |  |  | 0.031312 | 0.07563 | 0.052364 | 0.597958 | 0.2776 |
| Number of deaths with cases of aortic thrombembolism | 1 | 2 |  |  | 0.5 | 0.2857 | 0.2143 | 0.02521 | 0.031046 | 6.902618 | 0.00003 |
| Number of cases of stroke | 12 | 21 | 0.3243 | 0.2561 |  |  | 0.06823 | 0.277311 | 0.088659 | 0.769574 | 0.07353 |
| Number of deaths with cases of stroke | 4 | 5 |  |  | 0.3333 | 0.2381 | 0.095205 | 0.07563 | 0.052364 | 1.818124 | 0.03515 |
| Number of cases of cardiac thrombembolism | 14 | 28 | 0.3784 | 0.3415 |  |  | 0.036915 | 0.352941 | 0.094643 | 0.390044 | 0.34827 |
| Number of deaths with cardiac thrombembolic events | 3 | 12 |  |  | 0.2143 | 0.4286 | 0.214284 | 0.12605 | 0.065733 | 3.259938 | 0.00058 |
| Number of cases with pulmonary emboli | 5 | 11 | 0.1351 | 0.1341 |  |  | 0.000989 | 0.134454 | 0.067561 | 0.01464 | 0.49601 |
| Number of deaths with pulmonary emboli | 2 | 2 |  |  | 0.4 | 0.1818 | 0.218182 | 0.033613 | 0.035694 | 6.112534 | 0.00003 |
| Number of cases with DVT | 7 | 12 | 0.1892 | 0.1463 |  |  | 0.042848 | 0.159664 | 0.072543 | 0.59065 | 0.2776 |
| Number of deaths with DVT | 2 | 3 |  |  | 0.2857 | 0.25 | 0.035714 | 0.042017 | 0.039733 | 0.898845 | 0.18673 |
| Number of cases with splanchnic arterial thrombembolism | 3 | 12 | 0.0811 | 0.1463 |  |  | 0.06526 | 0.12605 | 0.065733 | 0.99281 | 0.16109 |
| Number of deaths with splanchnic arterial thromboembolism | 2 | 7 |  |  | 0.6667 | 0.5833 | 0.083336 | 0.07563 | 0.052364 | 1.591462 | 0.00592 |
| Number of cases with acute limb ischaemia | 2 | 12 | 0.0541 | 0.1463 |  |  | 0.092291 | 0.117647 | 0.063808 | 1.446386 | 0.07493 |
| Number of deaths with acute limb ischaemia | 2 | 9 |  |  | 1 | 0.75 | 0.25 | 0.092437 | 0.057362 | 4.358259 | 0.00003 |

The results of the analysis of the patients that survived in the 34 papers is shown in Table 20. The average age in this group was 74 and there were twice as many men as women in this group. The four main types of thromboembolic/ischaemic events in this group in increasing percentages were pulmonary emboli (16%), venous thromboembolic events not including pulmonary emboli (19%), cardiac thromboembolic/ischaemic events (26%) and stroke-related events including the carotid arteries (32%).

**Table 19.** Analysis of Cases Where Patients Died in 34 Papers with Individual Data.

|  |  |
| --- | --- |
| Number of Patients | 45 |
| Average Age of Patients | 73 (2 s.f.) |
| Number of Male Patients | 33 |
| Number of Female Patients | 12 |
| Number of Thromboembolic/Ischaemic Events | 84 |
| Average Number of Events Per Patient | 1.87 (2 d.p.) |
| Range of Number of Events Per Patient | (1-8) |
| Mode of Number of Events Per Patient | 1 |
| Number of Patients with Aortic Thromboembolic Events (%) | 3 (6.7% (1 d.p.)) |
| Number of Patients with Stroke Related Events Including Carotid Arteries (%) | 11 (24% (2 s.f.)) |
| Number of Patients with Cardiac Thromboembolic/Ischaemic Events (%) | 15 (33% (2 s.f.)) |
| Number of Patients with Pulmonary Emboli (%) | 4 (8.8% (1 d.p.)) |
| Number of Patients with Venous Thromboembolic Events (not including Pulmonary Emboli) (%) | 5 (11% (2 s.f.)) |
| Number of Patients with Thromboembolic Events in Splanchnic Arteries (%) | 9 (20%) |
| Number of Patients with Acute Limb Ischaemic Events (%) | 11 (24.4% (1 d.p.)) |

**Table 20.** Analysis of Cases Where Patients Survived in 34 Papers with Individual Data.

|  |  |
| --- | --- |
| Number of Patients | 74 |
| Average Age of Patients | 62.8 (1 d.p.) |
| Number of Male Patients | 49 |
| Number of Female Patients | 25 |
| Number of Thromboembolic/Ischaemic Events | 121 |
| Average Number of Events Per Patient | 1.635 (3 d.p.) |
| Range of Number of Events Per Patient | (1-7) |
| Mode of Number of Events Per Patient | 1 |
| Number of Patients with Aortic Thromboembolic Events (%) | 6 (8.1% (1 d.p.)) |
| Number of Patients with Stroke Related Events Including Carotid Arteries (%) | 24 (32% (2 s.f.)) |
| Number of Patients with Cardiac Thromboembolic/Ischaemic Events (%) | 32 (26% (2 s.f.)) |
| Number of Patients with Pulmonary Emboli (%) | 12 (16% (2 s.f.)) |
| Number of Patients with Venous Thromboembolic Events Not Including Pulmonary Emboli (%) | 14 (19% (2 s.f.)) |
| Number of Patients with Thromboembolic Events in Splanchnic Arteries (%) | 6 (8% (1 s.f.)) |
| Number of Patients with Acute Limb Ischaemic Events (%) | 3 (4% (1 s.f.)) |

### D-Dimers

One paper reported the units (Fibrinogen equivalent units or D-Dimer units) and there were 27 values which cited a laboratory reference range for interpretation of the results. The other values in the studies did not include the units or reference range and so our analysis was limited to the 27 values which cited the reference range. We therefore expressed the results as multiples of the reference range and the results of the analysis are shown in Table 22.

**Table 21:**
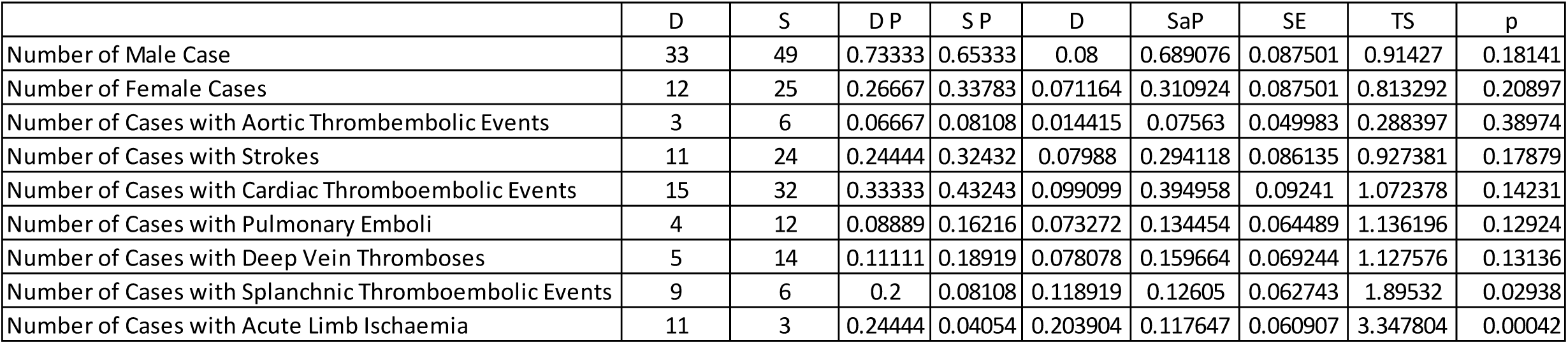
D – number who died, S - number who survived, DP – proportion of sample who died, SP – proportion of sample who survived, D – difference between proportions with smaller value subtracted from larger value, SaP – sample proportion for test statistic calculation, SE – standard error, TS – test statistic, p – p-value

**Table 22.** Benjamini–Hochberg procedure applied to values in Tables 18 and 21 with exception of deaths in cases of acute limb ischaemia. F – female, M – male, D – Deaths, S – Survival, p – p-value, (i/m)Q rank/number of values x false discovery rate (0.25)

|  | F | M | D | S | p | Rank | (i/m)Q |
| --- | --- | --- | --- | --- | --- | --- | --- |
| Number of Deaths (M/F) | 12 | 33 |  |  | 0.20897 | 15 | 0.163 |
| Number of Cases of Aortic Thromboembolism (M/F) | 2 | 7 |  |  | 0.2776 | 16 | 0.174 |
| Number of Deaths with Cases of Aortic Thromboembolism (M/F) | 1 | 2 |  |  | 0.00003 | 1 | 0.01 |
| Number of Cases of Stroke (M/F) | 12 | 21 |  |  | 0.07353 | 6 | 0.065 |
| Number of Deaths with Cases of Stroke (M/F) | 4 | 5 |  |  | 0.03515 | 5 | 0.054 |
| Number of Cases of Cardiac Thromboembolism (M/F) | 14 | 28 |  |  | 0.34827 | 17 | 0.185 |
| Number of Deaths with Cardiac Thromboembolic Events (M/F) | 3 | 12 |  |  | 0.00058 | 2 | 0.0217 |
| Number of Cases with Pulmonary Emboli (M/F) | 5 | 11 |  |  | 0.49601 | 19 | 0.207 |
| Number of Deaths with Pulmonary Emboli (M/F) | 2 | 2 |  |  | 0.00003 | 1 | 0.01 |
| Number of Cases with DVT (M/F) | 7 | 12 |  |  | 0.2776 | 16 | 0.174 |
| Number of Deaths with DVT (M/F) | 2 | 3 |  |  | 0.18673 | 14 | 0.152 |
| Number of Cases with Splanchnic Arterial Thromboembolism (M/F) | 3 | 12 |  |  | 0.16109 | 11 | 0.12 |
| Number of Deaths with Splanchnic Arterial Thromboembolism (M/F) | 2 | 7 |  |  | 0.00592 | 3 | 0.032 |
| Number of Cases with Acute Limb Ischaemia (M/F) | 2 | 12 |  |  | 0.07493 | 7 | 0.076 |
| Number of Deaths with Acute Limb Ischaemia (M/F) | 2 | 9 |  |  | 0.00003 | 1 | 0.01 |
| Number of Male Cases (D/S) |  |  | 33 | 49 | 0.18141 | 13 | 0.14 |
| Number of Female Cases (D/S) |  |  | 12 | 25 | 0.20897 | 15 | 0.163 |
| Number of Cases with Aortic Thromboembolic Events (D/S) |  |  | 3 | 6 | 0.38974 | 18 | 0.2 |
| Number of Cases with Strokes (D/S) |  |  | 11 | 24 | 0.17879 | 12 | 0.13 |
| Number of Cases with Cardiac Thromboembolic Events (D/S) |  |  | 15 | 32 | 0.14231 | 10 | 0.109 |
| Number of Cases with Pulmonary Emboli (D/S) |  |  | 4 | 12 | 0.12924 | 8 | 0.087 |
| Number of Cases with Deep Vein Thromboses (D/S) |  |  | 5 | 14 | 0.13136 | 9 | 0.098 |
| Number of Cases with Splanchnic Thromboembolic Events (D/S) |  |  | 9 | 6 | 0.02938 | 4 | 0.043 |

**Table 22.** Analysis of D-Dimer results expressed as multiples of upper limit of laboratory reference range.

| D-Dimers as multiples of upper limit of reference range | Value |
| --- | --- |
| Number of values | 27 |
| Mean | 21.6 (1 d.p.) |
| Standard Deviation | 41.5 (1 d.p.) |
| Standard Error of Mean | 7.98 |
| 95% Confidence Interval | 21.6 ± 15.6 (1 d.p.) |

### Fever

The papers in which the confirmation or exclusion of fever were reported are shown in Table 26. From these papers we identified 150 patients and these are summarised in Table 25.

**Table 23.** Analysis of Fibrinogen results expressed as multiples of upper limit of laboratory reference range.

| Ratio of Fibrinogen level to upper limit of reference range | Value |
| --- | --- |
| Number of values | 13 |
| Mean | 1.29 (2 d.p.) |
| Standard deviation | 0.277 (3 d.p.) |
| Standard error of mean | 0.077 (3 d.p.) |
| 95% Confidence Interval | 1.286 ± 0.15 |

**Table 24.** clinical correlates of 10 highest (D-Dimer/Reference range) values.

| Case | Paper | D-Dimer as multiple of reference range | Clinical Presentation |
| --- | --- | --- | --- |
| 1 | (La Mura et al, 2020) | 202 | 72 y/o male, Parkinson's disease, vascular dementia, E.Coli sepsis with hypotension as well as COVID-19, acute Portal Vein thrombosis, total occlusion of left portal venous system and branches of right portal |
| 2 | (La Berre et al, 2020) | 69 | 71 y/o male, previously healthy, free-floating aortic thrombus, thrombosis of right posterior tibial vein, pulmonary embolism |
| 3 | (Harsch et al, 2020) | 64.79 | 66 y/o female, bilateral pulmonary emboli, atrial fibrillation - unclear if this was new in onset, discharged |
| 4 | (Poggiali et al, 2020) | 48 | 82 y/o female, right common femoral vein DVT, acute renal failure, improved |
| 5 | (Vuilliamy et al, 2020) | 47 | 60 y/o male, bilateral acute lower limb ischaemia with no stenosis and minimal calcification, thromboembolectomy with improvement |
| 6 | (Zayet et al, 2020) | 38 | 84 y/o male, ischaemic stroke in multiple vascular areas, history of diabetes, hypertension, peripheral arterial disease, atrial fibrillation, patient died |
| 7 | (Oxley et al, 2020) | 27.6 | 44 y/o male, undiagnosed diabetes, left middle artery stroke, admitted to stroke unit |
| 8 | (Morassi et al, 2020) | 15.5 | 64 y/o male, acute myocardial infarct, acute renal failure, multiorgan failure, mechanical ventilation, multiple cortical and splenic infarcts, pulmonary embolism |
| 4 | (Poggiali et al, 2020) | 14 | Case 4 at 3 days of heparin infusion |
| 1 | (La Mura et al, 2020) | 8 | Case 1 at time of portal vein thrombosis diagnosis |

**Table 25.** Patients with fever.

|  |  |
| --- | --- |
| Number of patients where fever is excluded | 59 |
| Number of patients where fever is confirmed | 91 |

**Table 26.** Studies in which the exclusion or confirmation of fever is reported.

|  |
| --- |
| (Fara et al, 2020) |
| (Zayet et al, 2020) |
| (Gunasakeren et al, 2020) |
| (Escalard et al, 2020) |
| (Wang et al, 2020) |
| (Benussi et al, 2020) |
| (Morassi et al, 2020) |
| (Yaghi et al, 2020) |
| (Deliwala et al, 2020) |
| (Valderrama et al, 2020) |
| (Barrios Lopez et al, 2020) |
| (Co et al, 2020) |
| (Tunc et al, 2020) |
| (Viguier et al, 2020) |
| (Oxley et al, 2020) |
| (Hughes et al, 2020) |
| (Xiao et al, 2020) |
| (Cai et al, 2020) |
| (Lacour et al, 2020) |
| (La Berre et al, 2020) |
| (Poggiali et al, 2020) |
| (Harsch et al, 2020) |
| (Beccara et al, 2020) |
| (Vulliamy et al, 2020) |
| (de Barry et al, 2020) |
| (Zhou et al, 2020) |
| (Giacomelli et al, 2020) |
| (Qian et al, 2020) |
| (Bozzani et al, 2020) |
| (Bouaziz et al, 2020) |
| (Farina et al, 2020) |
| (La Mura et al, 2020) |
| (Varga et al, 2020) |
| (Bianco et al, 2020) |
| (Chan et al, 2020) |

The results show that there were 1.5 times as many patients with COVID-19 and thrombo-embolic events that were reported to have fever compared to those without fever.

### Reported Characteristics of Thromboembolic Events in COVID-19

The 56 identified papers were analysed and a thematic analysis was undertaken relating to the clinical thromboembolic/ischaemic features reported in patients with COVID-19. The themes are summarised in Table 27.

**Table 27.** Percentage of Papers Containing Comments on Specified Clinical Features of Covid-19.

| Reported Clotting Characteristic of Covid19 | Percentage of Papers Reporting Characteristics |
| --- | --- |
| No known risk factors | 19.60% |
| Thromboembolic event despite anticoagulation/antiplatelet therapy | 17.90% |
| High In Hospital Mortality | 16% |
| Asymptomatic prior to thromboembolic event | 12.50% |
| Cryptogenic/Without any source of Thromboembolism | 10.70% |
| Multiterritory stroke | 7% |
| Rethrombosis | 5.40% |
| Mild symptoms prior to presentation | 3.60% |
| Minimal or no improvement after revascularisation for stroke | 3.60% |
| No recanalisation after one pass for Stroke | 3.60% |
| Clot fragmentation with embolisation with intervention | 1.80% |
| Unusual location of clots | 1.80% |
| Non-detachable residual clots | 1.80% |
| Desert Foot | 1.80% |
| Low rate of successful revascularisation for ALI | 1.80% |
| Thrombosis of a graft | 1.80% |
| Clotting of medical devices | 1.80% |

#### A. No Known Risk Factors

This was the most commonly reported characteristic of thromboembolic events in COVID-19 in the papers. (Escalard et al, 2020) report on two patients under the age of 50 with developed stroke without risk factors. (Fara et al, 2020) report one previously healthy 33-year old lady who developed a thrombus in the common carotid artery extending to the internal carotid artery and associated with a middle cerebral artery thrombus. (Wang et al, 2020) report on two patients with stroke and no underlying medical risk factors including a patient in their 40’s. (Gunasakeren et al, 2020) report on a 40-year old lady without prior medical history with a large right middle cerebral artery stroke. (Yaghi et al, 2020) report on several patients who developed stroke with no significant medical history including two patients in their 40’s who died. (Barrios-Lopez e al, 2020) found no aetiology for stroke in two of their patients apart from hypercoagulation and systemic inflammation which were assumed to be COVID-19 related. (Vigueir et al, 2020) report on a 73-year old man with common carotid artery thrombosis with ischaemic stroke but no medical history or vascular risk factors.

(Stefanini et al, 2020) present a case series of STEMI which includes four patients between the ages of 45 and 67 with no medical risk factors. (Xiao et al, 2020) present a 62-year old man with inferior wall MI but without medical risk factors. (Poggiali et al, 2020) report on a 64-year old man without significant medical history who presented with a combination of deep vein thrombosis and subsegmental pulmonary embolism. (Diago-Gomez et al, 2020) present a previously healthy 50-year old who developed aortic thrombosis with acute limb ischaemia, DVT and stroke and a 69-year old male with no significant medical history who developed aortic thrombosis and pulmonary embolism.

#### B. High in-Hospital Mortality

(Escalard et al, 2020) report 60% mortality in their series of ten patients with large vessel stroke. (Wang et al, 2020) present a series of five patients with stroke with an average age of 52.8 years and a mortality of 60%. (Benussi et al, 2020) report a high in-hospital mortality in a COVID-19 cohort although not distinguishing between those with stroke and without. (Morassi et al, 2020) report 83% mortality in their case series of six patients with stroke. (Yaghi et al, 2020) compared their series of thirty two patients with COVID-19 and stroke with a control group and found an increased mortality in the COVID-19 group after adjusting for age and NIHSS score with an odds ratio of 64.87 (95% CI 4.44-987.28). (Middledorp et al, 2020) presented their findings in 199 patients with COVID-19 who were admitted to hospital. They found that venous thromboembolism was associated with high mortality and calculated a hazards ratio of 2.7 (95% confidence interval 1.3-5.8).

(Cui et al, 2020) report 40% mortality in twenty patients with COVID-19 and lower limb deep vein thrombosis. (Bellosta et al, 2020) report 40% mortality in their series of twenty patients with acute limb ischaemia. (Kaafarani et al, 2020) report on 141 critically ill patients with COVID-19 of which there were four cases of mesenteric ischaemia and one case of hepatic necrosis. Although they do not distinguish between the ischaemia and non-ischaemic pathology in the mortality, overall they report a mortality of 40% in those requiring surgery.

#### C. Thromboembolic Events Despite Anticoagulant or Antiplatelet Treatment

(Escalard et al, 2020) report in their case series that four out of ten patients with large vessel stroke were prescribed anticoagulant or antiplatelet treatment and one patient was on a combination of anticoagulant and dual antiplatelet treatment prior to the stroke. (Zayet et al, 2020) report on a patient who was being treated with apixaban for atrial fibrillation and subsequently developed ischaemic stroke in multiple areas. (Morassi et al, 2020) in their case series describe a patient with a previous myocardial infarct who was being treated with dual antiplatelet therapy and developed multiple ischaemic strokes and pulmonary embolism. They also report a man in his 80’s who was on aspirin but developed multiple ischaemic strokes and in hospital whilst receiving treatment with aspirin, clopidogrel and enoxaparin he developed another stroke. They further report on a lady in her 70’s who despite treatment with aspirin and warfarin develops multiple ischaemic strokes. They report on a man in his late fifties who despite treatment with Enoxaparin develops a dural sinus thrombosis but also a cerebral haemmorhage. (Barrios Lopez et al, 2020) report on a patient who develops ischaemic stroke whilst on bemiparin (a low weight molecular heparin) and another patient with known atrial fibrillation taking acenocoumarol prior to an ischaemic stroke.

(Lodigiani et al, 2020) draw attention to the high rate of arterial and venous thromboembolic events in patients with COVID-19 in their study (8%) despite anticoagulant prophylaxis and they believe that the figure may be higher due to undetected cases. (Lacour et al, 2020) report on a patient in his late 60’s with an anterior STEMI who is started on dual antiplatelet therapy and a bolus of heparin. Despite this he experienced another thrombus in the left anterior descending artery and after several interventions including IV unfractionated heparin experiences another thrombosis in the left anterior descending artery and dies. (Middledorp et al, 2020) in their cohort study reported on the development of venous thromboembolism in twenty-five patients (13% of the cohort) despite thromboprophylaxis.

(Klok et al, 2020) report the case of a lady in her 80’s who was started on low molecular weight heparin and who went on to develop a deep vein thrombosis. (Giacomelli et al, 2020) report on a 67-year old man with an aortic graft and aspirin prophylaxis who was commenced on Enoxaparin prophylaxis after admission. Despite this the graft occluded and he later died.

#### D. Asymptomatic prior to thromboembolic event

(Escalard et al, 2020) report two patients (20% of their sample) who were asymptomatic prior to large vessel stroke. (Wang et al, 2020) present a case series of five patients all of whom were normal two-and-a-half hours prior to presentation with stroke. (Yaghi et al, 2020) present five patients with ischaemic stroke who were asymptomatic prior to presentation. (Oxley et al, 2020) report two patients with no COVID-19 symptoms prior to presentation with large vessel stroke. (Stefanini et al, 2020) present twenty-four patients who presented with STEMI as the first clinical feature of COVID-19. (Xiao et al, 2020) report on two patients who present with sudden chest pain in the absence of other symptoms and confirmed acute myocardial infarct. (Lacour et al, 2020) present the case of a man in his sixties who is asymptomatic apart from chest pain with confirmation of myocardial infarct and is similarly asymptomatic prior to re-presenting with stent thrombosis.

#### E. Cryptogenic/Without any source of Thromboembolism

(Fara et al, 2020) report two cases of patients without obvious sources of thromboembolism including unremarkable echocardiography and no patent foramen ovale. (Yaghi et al, 2020) report cryptogenic strokes in most of their patients (21/32 (65.6%)). (Valderrema et al, 2020) report on a case of middle cerebral artery stroke with internal carotid artery thrombosis in the absence of patent foramen ovale or a cardiac embolus. (Barrios-Lopez et al, 2020) could not identify any source of emboli in two patients with ischaemic stroke and determined that this was secondary to a COVID-19-induced hypercoagulable state. (Stefanini et al, 2020) report in their case series that 11 (39.3%) patients did not have evidence of obstructive coronary artery disease. (Diago-Gomez et al, 2020) report on four cases of aortic thrombosis without atrial fibrillation or previous pro-thrombotic disease and attributed this to a COVID-19-induced hypercoagulable state.

#### F. Multi-territory Stroke

In their case series, (Escalard et al, 2020) report that fifty-percent of the patients with COVID-19 and stroke had multi-territory stroke involving the middle cerebral artery plus posterior or anterior cerebral artery involvement. (Zayet et al, 2020) describe two cases with ischaemic strokes affecting multiple vascular territories. (Wang et al, 2020) report one case with both anterior and posterior circulation ischaemic stroke. (Morassi et al, 2020) report multiple bilateral ischaemic strokes and suggest an embolic aetiology.

#### G. Rethrombosis

(Escalard et al, 2020) report four patients (40%) with reocclusion within twenty-four hours in their case series of ischaemic stroke. (Wang et al, 2020) describe their experience with mechanical thrombectomy in ischaemic stroke patients with COVID-19. They report two cases in which recanalisation with a stent retriever was followed by reocclusion within minutes and which they attributed to a hypercoagulable state. (Bellosta et al, 2020) report twenty cases of acute limb ischaemia with revascularisation. They identify a high rate of technical and clinical failure and attribute this to a hypercoagulable state.

#### H. Mild Symptoms Prior to Initial Presentation

(Escalard et al, 2020) report fifty percent of patients in their case series as presenting with mild symptoms at stroke onset. (Fara et al, 2020) present one case of a lady who was coughing prior to stroke but otherwise had no symptoms.

#### I. Minimal or No Improvement After Revascularisation for Stroke

(Escalard et al, 2020) report no significant neurological improvement in any of their patients 24 hours after mechanical thrombectomy for stroke. (Benussi et al, 2020) reported worse neurological outcome for COVID-19 patients with stroke compared to a control group without COVID-19.

#### J. No Recanalisation After One Pass for Stroke

The ability to recanalise an occluded blood vessel at first pass is associated with better outcome. (Escalard et al, 2020) reported an absence of first-pass effect for recanalisation in their series of ten patients with COVID-19 and ischaemic stroke. (Wang et al, 2020) report an average of just under three passes with the stent-retriever to achieve recanalisation.

#### K. Clot Fragmentation with Embolisation with Intervention

(Wang et al, 2020) report on the fragmentation of clots with intervention in a case series of patients with ischaemic strokes. They report on the intervention in one patient where aspiration was used initially with resulting embolisation from the carotid bulb thrombus distally. After using a stent-aspiration approach there was further embolisation of the thrombus into the middle cerebral artery. They report another case involving stent-aspiration of a thrombus in the internal carotid artery which embolised to the anterior cerebral artery. They report on another patient where a thrombus in the basilar artery was treated with a combination of balloon-guide catheter and aspiration resulting in embolisation to the posterior cerebral arteries. In another case they describe embolisation of fragments of a thrombus from the middle cerebral artery following stent-aspiration.The authors confirmed distant emboli in 100% of their cases. They also confirm embolisation into a different vascular territory in 40% of their cases which they contrast with a rate of 4.5% in a study involving patients without COVID-19 (Jovin et al, 2015).

#### L. Unusual Location of Clots

(Vigueir et al, 2020) report a case of a floating thrombus in the common carotid artery. They note that this is an unusual location for strokes resulting from occlusion within the cervicocephalic arteries and particularly in the absence of atheroma or dissection. They cite evidence that this location occurs in less than 1% of strokes involving the cervico-cephalic arteries (Singh et al, 2020).

#### M. Non-detachable Residual Clots

(Bellosta et al, 2020) describe the need for an additional surgical procedure in the treatment of acute limb ischaemia due to the occurrence of residual non-detachable clots.

#### N. Desert Foot

Desert foot refers to the occlusion of all of the main arteries of the foot. (Bellosta et al, 2020) refer to several cases of desert foot in their case series of 20 patients with acute limb ischaemia.

#### O. Low Rate of Successful Revascularisation of Acute Limb Ischaemia

(Bellosta et al, 2020) report a low rate of successful revascularisation in their case series of acute limb ischaemia. They note that patients receiving intravenous heparin did not undergo reintervention and a low oxygen saturation was significantly associated with unsuccessful revascularisation.

#### P. Thrombosis of a Graft

(Giacomelli et al, 2020) report on a case of a man in his late sixties with an abdominal aortic aneurysm that had been repaired with an aortic graft six years previously. At admission to hospital, the graft was patent. His overall condition deteriorated and nine days after admission, the graft was completed occluded with a thrombus and the patient died before revascularisation was possible.

#### Q. Clotting of Medical Devices

(Helms et al, 2020) report on 150 patients with COVID-19 who were admitted to four intensive care units in France. They report circuit clotting with the use of renal replacement therapy. They also report the thrombotic occlusion of the centrifugal pumps in patients receiving extracorporeal membrane oxygenation (ECMO). The centrifugal pump needed replacing after between 4 and 7 days. They also report that the average lifespan of the renal replacement therapy circuit was reduced by 50%. The authors hypothesised that the occlusion of the centrifugal pumps was due to a combination of ultrafiltration and high fibrinogen levels.

### Results of Exploratory Literature Search

We screened over 12,000 references from the clinical and scientific literature (see supplementary data) as well as references from prior and successive searches. The search strategy was determined from the specified aetiology unless there was sufficient evidence from the existing material.

### COVID-19-Related Coagulopathy Aetiologies Suggested in the 56 Main Papers

We identified 50 COVID-19 coagulopathy-related mechanisms suggested in the 56 main papers and have listed them in Table 28. Some of the mechanisms were mentioned by single authors whilst others such as a hypercoagulable state were mentioned by most of the authors.

**Table 28.** Coagulopathy-Related Aetiologies in COVID-19 Suggested in 56 Main Papers.

| Coagulopathy Related Aetiologies Suggested in 56 Main Papers |
| --- |
| **A. Hypercoagulable State** |
| Hypercoagulability/thrombophilic state |
| **B. Receptor Mediated** |
| ACE-2 mediated pathways via lung and heart binding |
| **C. Immune Mediated** |
| 1. Hyperinflammatory State |
| 2. SIRS mediated by IL-6 |
| 3. ARDS |
| 4. Secondary to septic shock |
| 5. Sepsis-associated Disseminated Intravascular Coagulopathy |
| 6. Cytokine storm/cytokine |
| 7. Immune mediated hyperviscosity |
| 8. Neutrophil extracellular traps |
| 9. Sepsis induced coagulopathy |
| 10. Immunothrombosis hypothesis |
| 11. Thromboinflammation/thrombogenesis |
| 12. Systemic inflammation with plaque disruption |
| 13. Lymphopenia |
| **D. SARS-Like Mechanisms** |
| **E. Direct Invasion of Tissues** |
| **1. Central Nervous System** |
| a. Direct virus nervous system injury |
| **2. Cardiovascular** |
| a. Cardioembolism 2nd to cardiac injury |
| b. Secondary to cardiogenic shock |
| c. Cardiovascular compromise |
| d. Myocarditis due to direct infection |
| **3. Gastrointestinal** |
| a. Viral enteroneuropathy |
| **4. Respiratory** |
| a. Respiratory pathology |
| b. Respiratory failure leading to myocardial mismatch perfusion |
| **5. Endothelial** |
| a. Microangiopathy leading to ischaemia |
| b. Endotheliatis |
| c. Endothelial Dysfunction |
| d. Vasculitis or vasculitis-like mechanism |
| e. SARS COV2 induced small vessel thrombosis |
| **E. Iatrogenic** |
| 1. Pharmacological adverse events |
| 2. Mechanical Ventilation |

**Table 28 (Continued)**
|  |
| --- |
| **F. Immune Mediated Syndromes** |
| 1. Antiphospholipid syndrome |
| 2. Type 1 interferonopathy |
| **G. General Factors** |
| 1. Critical illness encephalopathy |
| 2. Hypotension |
| 3. Hypertension |
| 4. Lymphopenia |
| 5. Atrial Fibrillation |
| 7. Immobilisation/Bed rest |
| 8. Hypoxia/hypoxia with vasoconstriction |
| 9. Stress cardiomyopathy |
| 10. Dehydration due to fever, diarrhoea |
| 11. Metabolic/electrolyte disturbance |
| 12. Low platelet count |
| 13. Plaques unstable due to mononuclear infiltrates, hypoxia, turbulence |
| **H. Secondary Infection** |
| 1. Septic embolisation with bacterial superinfection |
| 2. Secondary bacterial and fungal infection |
| 3. Stroke risk with infection |
| **I. Autopsy/Biopsy Finding Related Mechanisms** |
| 1. Pyroptosis |
| 2. Apoptosis |

#### A. Hypercoagulable/Thrombophilic State

In most of the main papers, the authors suggest that the coagulopathy results from a hypercoagulable state. Also termed a thrombophilic state, the hypercoagulable state is one which there is an increased likelihood of clotting or else a severe clotting response (Senst et al, 2020). A hypercoagulable state is one of the three components of Virchow’s triad and refers to the properties of the blood. Understanding the hypercoagulable state depends on an understanding of changes in the blood in COVID-19. In this section we treat this primarily as resulting from a change in the components of the blood although more general properties such as viscosity are considered in other sections.

Many studies have reported on the haematological, biochemical and immune parameters in COVID-19. (Henry et al, 2020) in their meta-analysis examined 21 studies (n=3377). They identified 27 laboratory values that were altered in severe or fatal COVID-19. These included an elevated prothrombin time, elevated D-Dimer, elevated CRP, IL-6 and myoglobin, cardiac troponin I and decreased platelet count. In a meta-analysis of coagulation dysfunction in COVID-19 (Jin et al, 2020) analysed 22 studies (n=4889) and found higher D-Dimer levels and prolonged prothrombin time in patients with more severe COVID-19. Non-survivors were more likely to have higher D-Dimer levels, decreased platelet count and increased prothrombin time compared to survivors.

(Di Minno et al, 2020) completed a meta-analysis looking at sixty subjects with 5487 patients with severe COVID-19 and 9670 patients with mild COVID-19. As with (Jin et al, 2020) they found evidence of higher D-Dimer levels in non-survivors compared to survivors while non-survivors had lower platelet levels. In another meta-analysis of patients with COVID19, 34 studies were included (n=6492) (Zhu et al, 2020). Their findings were similar to the previous meta-analyses. They found that patients with severe COVID-19 had lower platelet count, higher D-Dimer levels, higher fibrinogen, longer prothrombin time and shorter activated partial thromboplastin time. Again non-survivors were more likely to have higher D-Dimer levels. (Ibañez et al, 2020) found evidence of elevated D-Dimers and hypofibrinolysis in a prospective cohort study with COVID-19 in an ICU setting and suggest that the lungs are the source of the elevated D-Dimers. Increased levels of tissue factor in the blood in response to endothelial damage would fit with both explanations and is just one example of mediators of hypercoagulability in response to endothelial damage.

In summary, there is strong evidence of elevated fibrinogen and D-Dimers in severe COVID-19 compared to milder expressions of the illness. There is also evidence of decreased platelet count in COVID-19. Fibrinogen, D-Dimers and platelet count have an established role in clotting independent of COVID-19 (as per the DIC scores) and this may simply be a marker of the activation of the clotting pathways rather than providing insight into the aetiology. Thus there is the possibility that these changes are secondary to the vascular pathology including endothelial involvement that would that would form another aspect of Virchow’s triad. Increased levels of tissue factor in the blood in response to endothelial damage would fit with both mechanisms and is just one example of mediators of hypercoagulability in response to endothelial damage. We will consider other components of the blood and plasma that may contribute to hypercoagulability in subsequent sections.

#### B. Receptor-Mediated - ACE-2 receptor-mediated pathways

(Siddamreddy et al, 2020) suggest that SARS-COV2 can cause myocardial injury via the ACE-2 receptor. (Ciaglia et al, 2020) suggest that a reduced expression of ACE-2 receptors in older adults may increase susceptibility to more severe COVID-19.

The Renin-Angiotensin System (RAS) plays a central role in fluid homeostasis. There are a number of molecules, receptors and enzymes involved in the RAS (see Figure 9). One of the key functions is the tonic control of blood pressure which can be contrasted with the actions of the baroreceptors which drive changes more rapidly. Aldosterone release is an end-result of the action of Angiotensin II on the AT1 receptor which causes the retention of sodium in exchange for potassium. In hyperaldosteronism there is elevated blood pressure with hypokalaemia.

**Figure 9.**
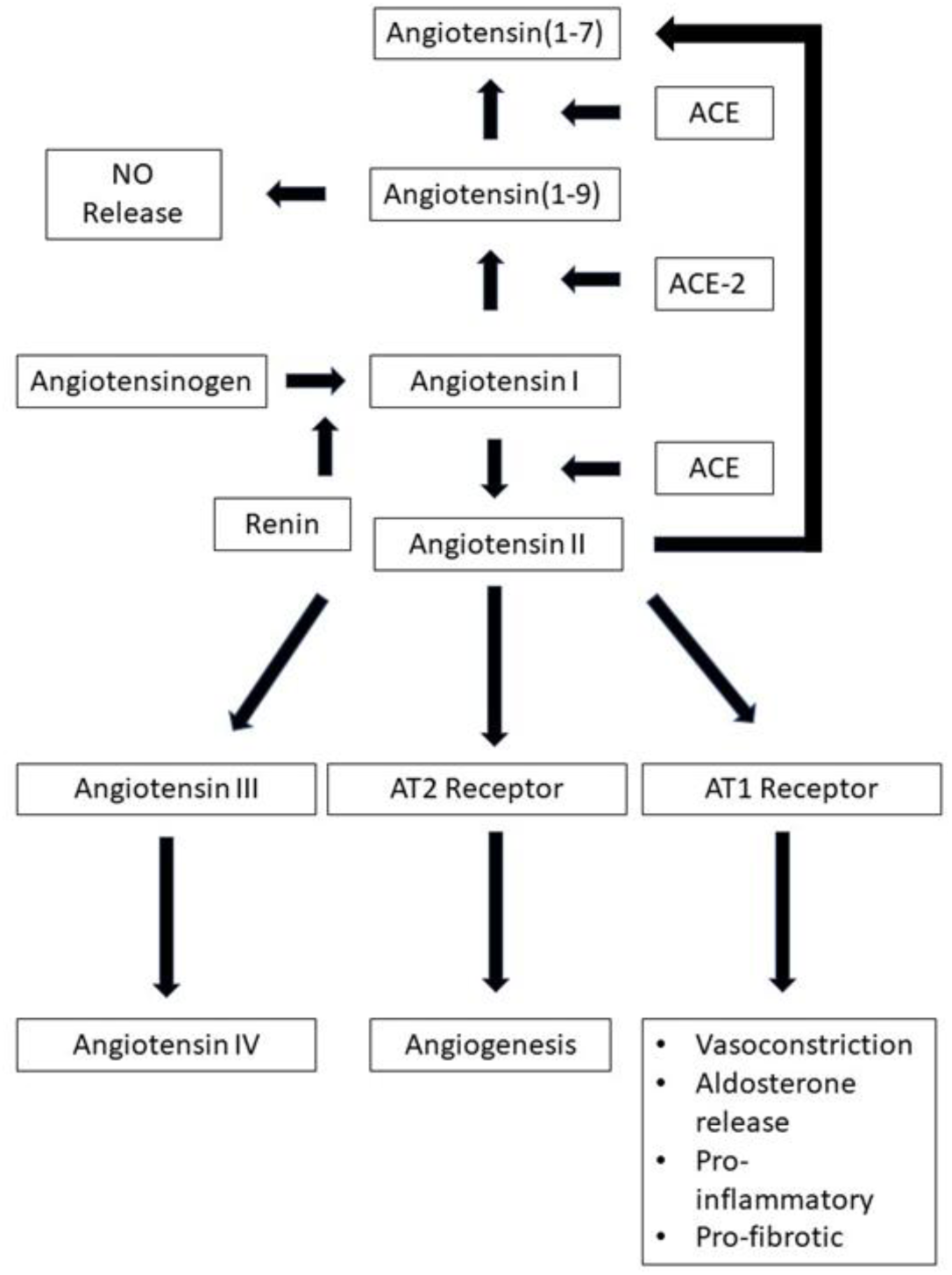
Overview of Renin-Angiotensin-Aldosterone System (RAAS)

(Miesbach, 2020) describes a reduction in ACE-2 receptors with SARS-COV2 infection and cites evidence of increased Angiotensin II levels in patients with COVID-19. (Miesbach, 2020) further outlines the mechanisms by which an increase in Angiotensin II levels could predispose to coagulation. (Miesbach, 2020) draws attention to the role of Angiotensin II in promoting smooth muscle cell proliferation which can contribute to atherosclerotic plaques, that Angiotensin II promotes the expression of Tissue Factor which in turn initiates coagulation and also promotes the expression and release of Plasminogen Activator Inhibitor-1 (PAI-1) which in turn inhibits fibrinolysis and promotes thrombus formation. (Senchenkova et al, 2019) provide evidence of a role for IL-6 and T-Cells in Angiotensin II-induced thromboinflammation. (Spillert et al, 1994) demonstrated an in-vitro procoagulant effect of Angiotensin II using a modified recalcification time test. (Lamas-Barreiro et al, 2020) note that the relationship between ACE-2 receptor activity and angiotensin II levels varies between organs. (Bryson et al, 2020) have in a model shown a protective effect from Angiotensin II-induced hypertension through antagonism of a prostaglandin receptor. Prostaglandins can inhibit platelet aggregation (Friedman et al, 2015) and it has been suggested that Eicosanoids such as prostaglandins may play a key role in COVID-19 (Hammock et al, 2020).

(Nguyen et al, 2013) Suggest that Angiotensin II’s actions may be mediated in part through reactive oxygen species (ROS) signalling. The evidence for a more ubiquitous presence of RAS is outlined by (Labandeira-Garcia et al, 2020). (Veras et al, 2020) found evidence of ACE-2 receptor and serine protease involvement in activation of NET’s. (Fang and Schmaier, 2020) review the role of the MAS receptor and the relationship between kallikrein/kinin and the RAA system in thrombosis.

In terms of ACE-2 receptors, a recent NICE review found no evidence to suggest that ACE-inhibitors or Angiotensin Receptor Blockers either increased the risk of contracting COVID-19 or else lead to a more severe manifestation of COVID-19 (NICE, 2020).

In their case study (Wang et al, 2020) report their findings in a man with COVID-19, ARDS and septic shock who experienced a marked response to the administration of angiotensin II. The authors discuss these findings whilst noting that a similar response has been found in non-COVID-19-related sepsis. (Liu et al, 2020) found elevated plasma angiotensin II levels in patients with severe COVID-19 compared to a control group with COVID-19.

(Garvin et al, 2020) analysed gene expression data from bronchial lavage specimens in patients with COVID-19 and used the Summit supercomputer to analyse the results. Their findings support the role of a bradykinin storm, an amplifying circuit of bradykinin production in response to the infection and mediated by RAS. They also found that the levels of hyaluronic acid were elevated and note that hyaluronic acid is associated with thrombosis. They suggest that the hyaluronic acid produces a gel in the lungs which interferes with the oxygenation of blood and thereby predisposes to hypoxaemia. (Garvin et al, 2020) looked at mRNA levels and found a reduction in ACE mRNA expression as well as an upregulation in ACE2 mRNA expression which may be expected to reduce the production of Angiotensin II. The mRNA levels do not necessarily correlate strongly with protein levels. The correlation between mRNA expression and protein levels (R^*2*^) was 0.4 across species in one study (de Sousa et al, 2009). This would suggest that the protein levels should be measured in preference to the mRNA or else in conjunction with this as (de Sousa et al, 2009) suggest that as much as 70% of the variance between mRNA and protein levels is accounted for by a combination of measurement accuracy and post-translation factors such as protein degradation.

(Kusadasi et al, 2020) also note the inter-relationship of the Renin-Aldosterone-Angiotensin system, complement system, Kinin-Kallekrein system and coagulation system as well as the relationship to ACE-2 while (Urwyler et al, 2020) report their initial findings with the use of Conestat Alfa In COVID-19, which targets the Kallikrein-Kinin system. (Curran et al, 2020) provide a model of pathology in COVID-19 involving a number of systems including the RAAS system and suggest that COVID-19 disrupts regulatory networks. (Wiese et al, 2020) hypothesise that pathology in COVID-19 arises from an upregulation of the classical arm of the RAS pathway and a downregulation of the ‘protective arm’ of the RAS pathway. (Czick et al, 2020) suggest a role for RAS imbalance in multiple aspects of COVID-19.

(Akoumianakis et al, 2020) note the relationship between obesity and dysregulation of the RAAS axis as well as myocardial and lung injury and suggest that this relationship may be relevant in COVID-19. In the Dyhor-19 Study (Villard et al, 2020) demonstrate a correlation between CRP and Aldosterone levels and COVID-19 severity. (Dudoignon et al, 2020) found that half of patients with COVID-19 and ARDS had acute kidney injury and this was significantly associated with activation of the RAAS with patients having high levels or renin and aldosterone on admission.

(Santamarina et al, 2020) provide evidence of ventilation/perfusion (V/Q) mismatch in the lungs and draw attention to two findings – well perfused areas of damaged lung and poorly perfused areas of healthy lung. They suggest this may explain the benefit of the proning position in patients with COVID-19 and ARDS. To partially explain this they suggest that Angiotensin II in COVID-19 results in vasoconstriction in the lungs and disrupts the ventilation/perfusion matching resulting in areas of healthy lung that are well perfused. (Lang et al, 2020) found evidence of pulmonary perfusion abnormalities in COVID-19 which supports the notion of disrupted perfusion/ventilation matching in COVID-19 secondary to RAA system disruption.

In summary, SARS-COV2 gains entry to cells via the ACE-2 receptor which in turn can lead to the downregulation of ACE-2 receptors and to an increase in Angiotensin II. There is evidence of elevated Angiotensin II levels in COVID-19. Angiotensin II in turn can lead to an increased risk of thrombosis. This can be considered as a component of the blood which leads to hypercoagulability. The relationship may not be so straightforward and may be organ-specific with the results of (Garvin et al, 2020) hinting at this complexity.

#### C. Sepsis-Related

##### 1. SIRS via IL-6

(Valderrema et al, 2020) suggest the septic inflammatory response syndrome (SIRS) mediated by IL-6 as one of the mechanisms that predisposes to ischaemic stroke in COVID-19. (Barrios Lopez et al, 2020) cite evidence that severe inflammation occurs during the acute phase of COVID-19. The difference between sepsis and SIRS as we saw from the introduction is one of organ dysfunction and a dysregulated immune response in sepsis in contrast with SIRS. If we consider a thromboembolic event then organ dysfunction is a function of the location of the event. The key question we should consider is whether there is a dysregulated immune response and in asking this question it becomes clear that SIRS cannot lead to a clotting event as this cannot be considered a *healthy response*. The utility of SIRS is lost due to semantics but we can still consider the associated mechanisms and the role of IL-6. However from a clinical perspective, the new sepsis consensus definition has removed the construct of SIRS, although there is an argument for the utility of SIRS (Sprung et al, 2016). We will not consider this further although we will add a section for IL-6 separately below. Also we consider septic shock on the continuum with sepsis and refer back to the introduction for discussion of the procoagulant mechanisms including DIC.

##### 2. IL-6

The suggestion of a key role for IL-6 in COVID-19 pathology is a basis for the recommendation for trialling IL-6-related therapies in COVID-19 (Ascierto et al, 2020). (Kuppalli and Rasmussen, 2020) suggest a potentially important role for IL-6 in the host response to SARS-COV2 and cite evidence for a reduced type-1 helper T-cell (Th1) antiviral response. However (Leisman et al, 2020) as well as (Sinha et al, 2020) outline the evidence against a central role for IL-6 in COVID-19.

(Liu et al, 2020) found a correlation between IL-6 levels and disease severity. (Luo et al, 2020) found a correlation between both IL-6 levels and CD8^+^ T cell counts and mortality. (Zhao et al, 2020) found IL-6 to be elevated in the later stages of severe COVID-19 but found that RANTES, a chemokine, was elevated earlier in the course of illness. (Mansouri et al, 2020) present a case of COVID-19 in which IL-6 and other parameters normalised after treatment with Colchicine and this was accompanied by a rapid improvement in presentation.

In a prospective comparative study (Giamarellos-Bourboulis et al, 2020) compared the immune responses of patients with COVID-19, influenza or bacterial sepsis. They found that in COVID-19 there was a more marked deterioration and they attributed this to immune dysregulation. This immune dysregulation was characterised by a disruption in antigen-presentation combined with lymphopenia but with monocytes producing IL-6 and TNF-α. IL-6 was elevated in patients with immune dysregulation. Immune dysregulation was characterised by a number of factors including absolute numbers of molecules of human leukocyte antigen on CD14 monocytes.

(Varhana and Wolchok, 2020) review the evidence for the immune response in COVID-19 and identify an accentuated innate immune response which results in elevated IL-6 levels. They suggest that a cytokine response syndrome could mediate pathology in COVID-19. However, they advise caution with this interpretation on the basis of significant differences between the typical description of the cytokine response syndrome and the features of COVID-19. They also identify a reduced adaptive response with T-cell exhaustion.

(Gubernatorova et al, 2020) suggest a key role for IL-6 in COVID-19 and provide a detailed review of the evidence. They note that IL-6 has properties which both promote and counter viral infections and they advise that further study of the role of IL-6 in COVID-19 is warranted. (Chatterjee et al, 2020) suggest that elevated IL-6 and hypoxia in COVID-19 could both lead to a reduction in Protein S and subsequent coagulopathy. In the non-COVID-19 literature (Hotter et al, 2019) demonstrated an association between IL-6 levels and size of infarcts in ischaemic strokes.

In summary, there are multiple lines of evidence to suggest that IL-6 plays a key role in COVID-19 but many details remain to be characterised and there is also evidence against IL-6 having a central role. There is a well-established evidence base for the role of IL-6 in infections independent of COVID-19 and there is also an association between IL-6 and strokes although there are questions about the direction of causality.

##### 3. Cytokine storm

Many of the authors in the 56 main papers have suggested that a COVID-19-related cytokine storm mediates the COVID-19-related coagulopathy. (Yaghi et al, 2020) cite evidence of cytokine storm association with COVID-19 (Chen et al, 2020). (Jain et al, 2020) suggest that a subset of patients with COVID-19 may have a cytokine storm. (Barrios-Lopez et al, 2020) again suggest that a small subset of patients with COVID-19 may experience a cytokine storm and cite evidence that this is associated with high mortality (Zhou et al, 2020).

The cytokine storm is a dysregulated elevation of cytokines that results in pathology. (Tisoncik et al, 2012) review the cytokine storm and note the first description in a graft-versus-host response paper in 1993. (Tisoncik et al, 2012) also outline the three main types of resulting pathology – endothelial dysfunction, pulmonary fibrosis and inflammatory responses and note that the lung injury can progress to ARDS. They also note a number of viruses associated with a cytokine storm in a number of the early descriptions (see Table 29).

**Table 29.** Infectious agents associated with cytokine storm response adapted from (Tisoncik et al, 2012)

|  |
| --- |
| Cytomegalovirus |
| Epstein-Barr virus-associated hemophagocytic lymphohistiocytosis |
| Group A streptococcus |
| Influenza virus |
| Variola virus |
| SARS-CoV |

(Shimizu, 2019) outlines the features of the cytokine storm syndromes including those resulting from IFN-γ, TNF, IL-6, IL-1β and IL-18 and note the features of cytokine storm syndrome including fever, hepatomegaly and splenomegaly.(Van der Poll et al, 2000-2013) note the procoagulant effects of IL-1, IL-2, IL-6, IL-12 and TNF.

(Barnes et al, 2020) provide an overview of the elevated cytokines in COVID-19 with reference to the literature (see Table 30 but also the earlier section on IL-6). (Coperchini et al, 2020) provide an overview of the cytokine storm in COVID-19. They suggest that ARDS in COVID-19 results from a cytokine storm. (Manjili et al, 2020) make a case for COVID-19 as an acute inflammatory disease and provide evidence of an association between elevated cytokine levels and more severe illness.

**Table 30.** Elevated cytokines in COVID-19 (Barnes et al, 2020)

|  |
| --- |
| IL1Beta |
| IL2 |
| IL6 |
| IL7 |
| IL8 |
| IL10 |
| IFNGamma |
| IFN-inducible protein 10 |
| Monocyte chemoattractant protein 1 |
| G-CSF |
| Macrophage inflammatory protein 1 alpha |
| TNF alpha |

In summary, there is strong evidence for the phenomenon of cytokine storms and there are clinical syndromes correlating with the relevant cytokines and evidence for a procoagulant action depending on the cytokine. There is also evidence of elevated cytokine levels in COVID-19 and a relation to the severity of illness. We have considered IL-6 separately as a special case.

##### 4. Immune-mediated Hyperviscosity

(Valderrema et al, 2020) suggest the IL-6 immune mediated response as mediating hyper-viscosity and cite evidence for this (Chen et al, 2020). Hyperviscosity in turn can increase the risk of stroke. Waldenström macroglobulinemia represents an example of an immunemediated plasma hyperviscosity (Gertz, 2019) resulting from elevated levels of IgM although there is evidence for a role for IL-6 (Poulain et al, 2008). (Maier et al, 2020) found evidence of hyperviscosity in COVID-19 but were not clear on what caused this although suggesting fibrinogen. In summary, we conclude that there is evidence of hyperviscosity but it is unlikely to result from elevated IL-6.

##### 5. Immunothrombosis hypothesis/thromboinflammation/hyperinflammation

(Valderrema et al, 2020) suggest an immune-mediated thrombogenic mechanism as do (Escalard et al, 2020) who in turn cite (Varga et al, 2020). (Klok et al, 2020) suggest thromboinflammation as a mediator of coagulopathy and in turn cite (Connors and Levy, 2020) who review the evidence for this. (Connors and Levy, 2020) cite evidence for a strong correlation between IL-6 and fibrinogen in COVID-19.

The immunothrombosis hypothesis is at the simplest level a contemporary restatement of the inflammatory theory of thrombosis promoted by Cruvielhier. (Schattner et al, 2020) note the origin of the term thromboinflammation in 1994 to describe arterial stent restenosis-related platetet-neutrophil interactions. A key paper in the modern restatement of the immunothrombosis hypothesis is (Nieswant et al, 2011). (Nieswant et al, 2011) have outlined the basis for a thrombotic cascade leading to ischaemic stroke. In their model, the thrombotic cascade begins when the platelet Gp1 receptor binds to von Willebrand factor released from granules in the endothelium after vessel wall injury. This binding is transient and slows down the platelets sufficiently so that Gp6 can bind with collagen at the site of the vessel wall injury. Following this there is upregulation of integrin receptors which facilitate platetet aggregation. The Gp6 also activates Factor XII leading both to the coagulation cascade and also the activation of the kallikrein-kinin system leading to inflammation and to the end-product bradykinin. There is evidence that the Gp1-VWF-Gp6 pathway is not involved in acute ischaemic stroke injury however and instead it has been suggested that this is mediated by interactions with a damaged vessel wall (Denorme et al, 2019).

(Mitchell, 2020) discusses the role of thrombin in thromboinflammation, driving the coagulation cascade and the immune system and activating leucocytes and other cells to release cytokines. (Bray et al, 2020) discuss a potential role for thromboinflammation in microvascular thromboses. They note that the definition of microvessels can vary but in their paper they refer to the arterioles, capillaries and venules in this category. They also note a vast increase in shear stress in the arterioles compared to the venules sufficient to impact on the dynamics of blood flow as well as endothelial cell morphology.

Platelets interact with a number of immune cells including neutrophils, leucocytes, monocytes and macrophages in a diverse range of immune-related functions (Rayes et al, 2020). Platelets contain α-granules with a variety of over 300 proteins, lysosomes containing proteolytic enzymes and dense granules containing molecules including serotonin, calcium and magnesium which mediate platelet activation (Stegner et al, 2019). (Guo and Rondina, 2019) distinguish between immunohemostasis (trapping pathogens with a combination of coagulation and inflammation and involving blood vessel injury without pathology) and thromboinflammation (vasculature-based pathological responses in response to infectious and immune triggers). (Guo and Rondina, 2019) associate thromboinflammation with DIC, viraemia, stroke, DVT and myocardial infarct. There are a number of diseases where thromboinflammation has been suggested to play a significant role including Behçet’s syndrome (Becatti et al, 2019) and delayed ischaemia after cerebral haemmorhage (McBride et al, 2017).

Thromboinflammation has been suggested as a mechanism which mediates the effects of snake venoms (Teixeira et al, 2019). The relationship between platelets and complement are outlined in (Eriksson et al, 2019). The role of platelets in mediating thromboinflammation in myeloproliferative disorders is examined in (Marin and Heller, 2019). (Senchenkova et al, 2019) provide evidence that IL-6 mediates angiotensin-II mediated thrombosis and that IL-6 potentiates platelets to activate Gp6 receptors.

There has been a suggestion that the von Willebrand Factor/ADAMTS13 axis plays a pivotal role in thromboinflammation and has been a focus for exploring therapeutic interventions (Gragnango et al, 2017). There is evidence that endothelial cell-derived von Willebrand factor plays a role in post-ischaemic stroke related thrombinflammation (Dhanesha, 2016). The lectin pathway has also been suggested to play a role in thromboinflammation (Kozarcanin et al, 2016). (Jayarangaiah et al, 2020) present a thrombinflammatory model of COVID-19-related coagulopathy.

We have included the term hyperinflammation here and would consider that the relation with a coagulopathy is mediated via a number of mechanisms including cytokines which is considered in a previous section and through the mechanisms of thromboinflammation. We note however that (Webb et al, 2020) have proposed and validated criteria for a COVID-19-related hyperinflammatory syndrome. (Lima et al, 2020) report a case of hemophagocytic syndrome in which there was a hyperinflammatory response with macrophage activation. (Cole et al, 2020) suggest that low grade inflammation with diabetes and adipose tissue dysfunction in obesity is a vulnerability in people who develop COVID-19. (Giamerellos et al, 2020) found evidence of two types of hyperinflammatory response in COVID-19 with severe respiratory failure – immune dysregulation and macrophage activation syndrome. They found that IL-6 results in immune dysregulation with monocytes producing excessive inflammatory cytokines and also resulting in CD4 lymphopenia and B-cell lymphopenia. They also found that IL-6 elevation was associated with a reduction in CD14 monocyte HLA-DR expression and that the HLA-DR expression increased in convalescence. They also found that IL-1β levels increased and resulted in macrophage activation syndrome.

In summary, there is evidence of thromboinflammation in COVID-19 and independent of COVID-19, platelets have a central role in thromboinflammation.

##### 6. Systemic inflammation with plaque disruption

(Morassi et al, 2020) discuss the association between sepsis and rupture of plaques. In the pre-pandemic literature (Maillet et al, 2012) present a case of infective aortitis. The case is characterised by a bacterial infection of the aortic intima which is an unusual location. The authors suggest that the atherosclerotic plaques may have resulted in a vulnerability to aortic infection enabling direct seeding or alternatively that the bacteria infiltrated via the vasa vasorum. The case demonstrates a possible relationship between arterial pathology and susceptibility to infection of the vessel wall which is relevant to the question of SARS-COV2 and vascular pathology. However we would group this under both secondary infection and vulnerabilities in COVID-19.

##### 7. Neutrophil extracellular traps

(Escalard et al, 2020) suggest neutrophil extracellular traps (NETs) as one possible explanation for the absence of reperfusion with recanalisation in their case series of thrombectomy for ischaemic stroke. They reference (Barnes et al, 2020) and (Varga et al, 2020) in relation to the combination of NETs and endothelial involvement. (Varga et al, 2020) identified widespread endothelial involvement together with neutrophil as well as mononuclear infiltration of a large arterial vessel in COVID-19. (Barnes et al, 2020) present the case for NETs as a driver of the pathology in COVID-19. NETs are a mechanism by which neutrophils eject intracellular material including DNA and enzymes such as neutrophil elastase which form an extracellular web which can destroy pathogens but can also cause collateral damage to other cells.

(Barnes et al, 2020) cite evidence for more severe disease with both neutrophilia and a high neutrophil-to-lymphocyte ratio and suggest that neutrophilia may be relevant to the role of NETs. They cite evidence of the relationship between NETs and ARDS, the regulation of neutrophils by cytokines and the relevance to the cytokine storm, NET-mediated thrombosis and a similarity between the mucous production in the airways in both COVID-19 and Cystic Fibrosis for which they cite evidence of NET involvement in the latter.

(Zuo et al, 2020) found evidence of NETosis in patients with COVID-19 in a hospital setting while (Cicco et al, 2020) suggest NETs and Damage-Associated Molecular Patterns (DAMPs) as potential treatment targets. (Radermecker et al, 2020) found evidence of NETs in the airways, interstitium and vascular spaces in the lungs in COVID-19 postmortems cases (n=4). (Middleton et al, 2020) provide evidence of NETs triggering thromboinflammation in COVID-19 in a prospective cohort study while (Beswick et al, 2020) find a role for NETs in ARDS-associated thromboinflammation in COVID-19. (Wang et al, 2020) identified a correlation between neutrophil activation in 55 patients with COVID-19 and 17 NET-associated genes. (Leppkes et al, 2020) found evidence of pulmonary microvessel occlusion with NETs. (Tomar et al, 2020) suggest NETs as a source of necroinflammation leading to endothelial cell death and promoting thrombosis.

(Liu et al, 2020) found that neutrophil-to-lymphocyte ratio (NLR) was prognostic for critical illness in COVID-19 in their prospective cohort study (n=61). Using a cut-off of NLR of 3.13 they found that 50% of patients aged over 50 with an NLR > 3.13 developed critical illness. (Lian et al, 2020) demonstrated prognostic significance of the neutrophil-to-lymphocyte ratio. (Moutchia et al, 2020) found four predictors of a more severe course of COVID-19: high markers of innate immunity (e.g neutrophils, neutrophil-to-lymphocyte ratio), low markers of adaptive immunity (e.g. lymphocytes), high markers of tissue and organ damage (e.g. LDH and markers of renal function).

In summary, there is evidence of the involvement of neutrophils in COVID-19 and there is an evidence base for NET involvement in a number of pathologies relevant to COVID-19.

##### 8. SARS-Like Mechanisms

(Deliwala et al, 2020) note a similarity between infections with SARS and SARS-COV2. They note a common association with stroke and that the SARS viral load plays a role in the SARS-related coagulopathy and endothelial dysfunction. SARS is a useful model for COVID-19 as it is similar to SARS-COV2, the clinical features of SARS infection have an overlap with COVID-19 and there is a body of research on SARS. The number of papers on COVID-19 including pre-prints however exceeds that of SARS. Additionally a number of clinical features in SARS are less clear as the overall number of cases was smaller than in COVID-19.

Both ACE-2 and TMPRSS2 are required for the entry of both SARS (Heurich, 2014) and SARS-COV2. (Franks and Galvin, 2013) review the evidence on SARS and note that watery diarrhoea occurs in 70% of cases and fibrin thrombi may also be present. (Cleri et al, 2010) provide an overview of the clinical presentation of SARS. (Singh et al, 2003) describe a case of SARS without the typical findings and reference evidence that SARS presentations range from asymptomatic to severe and fatal. (Wan et al, 2007) found evidence of a correlation between radiographic lung lesions and clinical presentation.

(Zarhariadis et al, 2006) demonstrated coinfection in SARS. (Ng et al, 2005) report a case of pulmonary artery thrombosis and cite the evidence for both pulmonary emboli and deep vein thrombosis in SARS. (Chen et al, 2004) found evidence of a history of stroke, hypertension and diabetes in patients who developed ARDS in SARS. (Wei et al, 2006) found evidence of thyroid involvement in SARS. (Vijayanand et al, 2004) review the evidence of SARS and identify a number of factors associated with worse prognosis including older age, lymphopenia, high peak LDH and comorbid conditions.

(Ding et al, 2003) report on an autopsy series in SARS and find localised fibrinoid necrosis and small vein thrombosis. (Mazzulli et al, 2004) found evidence of SARS-COV in lung tissue in post-mortem and also found a relationship between viral load and duration of illness. (Farcas et al, 2004) reported on the autopsy findings in people with SARS finding a distribution of virus in the lungs in 100% of cases, in the bowel in 73% of cases, the liver in 41% of cases and the kidneys in 38% of cases with the highest viral loads being in the lungs and bowel respectively. (Hwang et al, 2005) reported on an autopsy series of 20 patients with SARS and found evidence of damage to the vascular endothelium of small and medium pulmonary vessels as well as vascular fibrin thrombi with pulmonary infarcts. (Gu and Korteweg, 2007) review the pathology and pathogenesis of SARS.

(Frieman and Baric, 2008) review the mechanisms of severe acute respiratory syndrome pathogenesis and innate immunomodulation. (Gralinski et al, 2018) suggest SARS pathogenesis as predominantly immune-mediated in their review of the role of the complement pathway. (Chng et al, 2005) identified the course of the haematological parameters in SARS. (McBride and Fielding, 2012) provide an overview of the role of the accessory proteins in SARS pathogenesis. (Venkataraman and Frieman, 2017) review the evidence for the role of epidermal growth factor receptor signaling in pulmonary fibrosis in SARS.

The mechanisms of evasion of the immune system are outlined in (Perlman and Netland, 2009). (Jones et al, 2003) identified prolonged dysregulation of the cytokine response in patients who had experienced SARS. (Tan et al, 2007) outline the evidence for apoptosis and necrosis induced by SARS and also identify three of the accessory proteins which lead to apoptosis. (Lau and Peiris, 2005) suggest a lack of a type-1 interferon response in SARS and note that lymphopenia with a decrease in both CD4+ and CD8+ T-cells is a common occurrence. (Nicholls et al, 2003) suggested similarities between SARS and H5N1 with haemophagocytosis possibly related to cytokine dysregulation and lymphopenia together with white-pulp atrophy of the spleen. (van den Brand et al, 2014) review the pathogenesis of SARS in this paper and comment on the role of immunosenescence to account for the vulnerability of the older adult population. (Totura and Baric, 2012) review the role of the innate immune system in response to SARS COV pathogenesis. (Thiel and Weber, 2008) review the interferon and cytokine responses to SARS-coronavirus infection.

In summary, the literature on SARS is well established with an outline of the host response to infection. This knowledge is not directly relevant to the COVID-19 model we have developed but is useful as an analogous model for comparison. There are many similarities between SARS and SARS-COV2 including the mechanism of entry to the cell and to various aspects of the course of the infection and the clinical findings. The similarities include the following: the clinical picture varies from asymptomatic to fatal, with ARDS, pulmonary emboli and DVT described, SARS was identified in the lungs, bowel, liver, kidneys and vascular endothelium. Age, lymphopenia and LDH levels are prognostic and the host-immune response plays an important role in pathogenesis.

##### D. Direct Invasion of Tissues

###### a. Direct virus nervous system injury

In the 56 main papers (Morassi et al, 2020) note encephalopathy preceding stroke and could not exclude direct CNS invasion as a possibility. (Valderrama et al, 2020) also suggest the possibility of direct CNS invasion in relation to neurological involvement in COVID-19.

(Kishfy et al, 2020) report on a case of Posterior Reversible Encephalopathy Syndrome and suggest direct endothelial involvement. (Agagholi et al, 2020) cite evidence of encephalitis, meningoencphelitis and detection of SARS-Cov2 in the CSF. In their review of the literature, (Ellul et al, 2020) report evidence of encephalopathy, encephalitis, meningoencephalitis and disseminated encephalomyelitis in COVID-19. They also discuss the possibility of a trans-olfactory route into the central nervous system. (Ashraf et al, 2020) report two cases of seizures with COVID-19 and draw attention to evidence in other research of SARS-COV2 inclusions in nervous tissue samples.

The question arises of whether a direct invasion of the nervous system is responsible for strokes or else there are other factors including those relating to coagulopathy which are responsible. In the 56 main papers, we identified a number of cases with thrombi, sometimes extensive, arising in the carotid arteries and typically involving the middle cerebral arteries and being sufficient to account for the strokes. The role of direct invasion of the tissue has played a role in ischaemic damage in a case of necrotising encephalopathy (Poyiadji et al, 2020) and so this is a mediator for ischaemic damage but significantly less so than ischaemic stroke resulting from thromboembolism in the large vessels. Indeed (Kato et al, 2020) in their comparison of neurological manifestations of SARS-COV2 with MERS and SARS suggest that strokes in COVID-19 most likely result from a combination of a hypercoagulable state and endothelial dysfunction. Nevertheless the findings of (Morassi et al, 2020) raise the possibility of direct CNS invasion predisposing to stroke risk.

###### a. Direct Invasion of the Myocardium

(Valderrama et al, 2020) suggested that SARS-COV2 may directly invade the myocardium and lead to cardiac thrombi, atrial tachyarrythmias and ACE-2 receptor-mediated mechanisms. A recent study identified abnormalities in 55% of echocardiograms of 667 patients supporting the notion of cardiac involvement in COVID-19 (Dweck et al, 2020). (Momtazmanesh et al, 2020) provide strong evidence of myocardial involvement in COVID-19 in their systematic review and meta-analysis. (Hendren et al, 2020) describe a COVID-19 related acute cardiovascular syndrome which can include acute coronary syndrome (STEMI or NSTEMI), acute myocardial injury without obstructive coronary artery disease, arrhythmias, heart failure with or without cardiogenic shock, pericardial effusion with or without tamponade and thromboembolic complications.

In summary there is strong evidence for myocardial pathology in COVID-19, although the papers we identified for this section did not provide evidence of direct viral invasion of the myocardium although this is revisited in a subsequent section. Therefore we can say that there is strong evidence of an association between SARS-COV2 infection and myocardial pathology. The consequences of myocardial pathology are considered separately in subsequent sections.

###### b. Secondary Effects on the Myocardium: Stress Cardiomyopathy

(Yaghi et al, 2020) have proposed stress cardiomyopathy as a potential mediator of coagulopathy in COVID-19. In stress cardiomyopathy, physical or emotional stressors place additional demands on the heart leading to a cardiomyopathy. (Sattar et al, 2020) report a case of COVID-19 presenting with Takotsubo Cardiomyopathy with atrial fibrillation. (Jabri et al, 2020) report a significant increase in presentations of stress cardiomyopathy in patients compared to the pre-pandemic period. As with atrial fibrillation, the additional stresses placed on the heart may be a marker of the critically unwell state of a patient and so this can also be considered under the heading of general factors. (Okura, 2020) reviews the diagnostic criteria for Takotsubo Syndrome and the occurrence in COVID-19.

In the non-COVID-19 literature (El-Battrawy et al, 2020) found no difference in the rate of thromboembolic events including stroke between recurrent and non-recurrent Takotsubo Syndrome groups although finding an incidence of between 1.32 and 3.3%.

In summary there is moderate evidence of an association between COVID-19 and stress cardiomyopathy but there is a well-established evidence base for the consequences of stress cardiomyopathy independent of COVID-19. The critically ill patient may provide a special case of stress cardiomyopathy representing a number of psychological and physical predisposing factors and also a substantial number of patients in terms of the scale of the pandemic.

###### c. Cardioembolism 2nd to cardiac injury

In the 56 main papers, there was evidence of cardiac thrombi with a left ventricular thrombus being described (Moyashedi et al, 2020). Other authors have similarly identified cardiac thrombi in patients with COVID-19. (Soltani and Mansour, 2020) report a highly unusual case of COVID-19 presenting with biventricular thrombi. (Sethi et al, 2020) report a case of clot-in-transit in the right ventricle and describe the implications for the pulmonary embolism response team. (Horowitz et al, 2020) report another case of clot-in-transit in the right ventricle in a proned patient with ARDS. (Sulemane et al, 2020) report a case of pulmonary embolus with intramural right ventricular thrombus. Although this is evidence of cardiac thrombi this does not differentiate between an in-situ origin resulting from myocardial invasion or else an embolic origin. In summary, there is evidence of cardiac thrombi in the form of case reports in COVID-19 but we have not identified evidence linking the thrombi either to direct myocardial invasion or cardiac injury. Nevertheless there is a robust evidence base for cardiac injury and cardioembolism independent of COVID-19 and there is also overlap with subsequent sections.

###### d. Secondary to cardiogenic shock

(Barrios-Lopez et al, 2020) suggest cardiogenic shock in COVID-19 as a potential mediator of ischaemic stroke. In their systematic review, (Shafi et al, 2020) provide evidence of cardiogenic shock in COVID-19. They cite evidence both of an association with high mortality and also with direct SARS-COV-2 invasion of the myocardium.

(Warkentin and Pai, 2016) suggest ‘shock liver’ as a mediator for disseminated intravascular coagulation occurring with cardiogenic shock. (Akkus et al, 2009) found evidence of increased plasminogen activator inhibitor-1 (PAI-1) in patients with cardiogenic shock after myocardial infarct. (Schiessler et al, 1994) report their findings in shock-associated coagulopathy in a group of patients receiving mechanical circulatory support and heart transplantation.(Hanaki et al, 1991) report increased levels of leucotoxin in two patients with infective endocarditis and cardiogenic shock. (Egbring et al, 1986) investigated infection-related cardiogenic shock and identify proteinase inhibitor complexes as an important mediator and discuss the diagnostic and therapeutic implications.

In summary there is evidence of an association of SARS-COV2 infection with cardiogenic shock. Independently of COVID-19, there is an established evidence base on cardiogenic shock-related coagulopathy.

###### e. Cardiovascular compromise

Cardiovascular compromise is a broad term which encompasses many pathologies with the potential for coagulopathy including cardiogenic shock and myocardial involvement. (Bansal, 2020) mentions altered myocardial demand-supply ratio in a review of cardiovascular disease and COVID-19 in addition to other pathologies which impact on cardiovascular function. At present we would consider this as a broad category meriting further characterisation.

###### f. Myocarditis due to direct infection

(Stefanini et al, 2020) reported 11/28 cases of STEMI in which no culprit lesion was found and suggested a number of mediating mechanisms including myocarditis resulting from SARS-COV2 infection. Myocarditis has been reported as an important feature of COVID-19 early in the pandemic (Inciardi et al, 2020). (Shafi et al, 2020) review the evidence for COVID-19 associated myocarditis in their paper.

(Antoniak et al, 2008) provide evidence that viral myocarditis is associated with a hypercoagulable state and that this is associated with expression of tissue factor in the myocardium. (Pickens and Catterall, 1978) describe a case of disseminated intravascular coagulation and myocarditis with Mycoplasma pneumoniae infection lending evidence to the association of viral myocarditis with a coagulopathy although this does not discount the involvement of other mechanisms described here. (Chimenti et al, 2020) report on a case of infarct-like myocarditis resulting from Epstein-Barr virus. (Salacki and Wysokiński, 2016) report on a case of acute myocardial ischaemia associated with myocarditis and antiphospholipid syndrome. There is an established literature on Chagas disease which is divided into acute and chronic phases and can lead to a coagulopathy (Woudstra et al, 2018).

In summary there is evidence of myocarditis in COVID-19. Independently of COVID-19 there is an established literature on the association of myocarditis with a coagulopathy and also with ischaemia. Thus the possibility of a myocarditis-like mechanism for COVID-19-related coagulopathy, particularly for myocardial events, is supported by some evidence but of an association rather than causality. Furthermore there is limited evidence of this relationship.

###### a. Viral enteroneuropathy

(Kaafarani et al, 2020) suggest viral enteroneuropathy as a possible mediator of the GI pathology they have described. However their case series encompasses different groups of pathology including disorders of gastrointestinal motility as well as mesenteric ischaemia. In the cited reference (Wells et al, 2017) enteroneuropathy is suggested as a mediator for pseudo-obstruction.

(Bostancıklıoğlu, 2020) suggests that various pathways between the gut and the central nervous system that could mediate spread of SARS-COV2. (Briguglio et al, 2020) discuss the potential role of the vagus nerve in transmitting infection from the enteric nervous system to the central nervous system whilst also noting that the physical barriers as well as the gut-associated lymphoid tissue afford protection to the enteric nervous system. Nevertheless just as with the ischaemic strokes, the authors in the 56 main papers provided evidence of thromboembolic events in the splanchnic arteries which could account for the events including small bowel necrosis. In summary, viral enteroneuropathy may be more relevant to gastric dysmotility rather than mesenteric ischaemia.

###### a. Respiratory pathology

This term is broad and we will refer to this as requiring further characterisation.

###### b. Respiratory failure and hypoxia leading to myocardial injury

(Siddameddy et al, 2020) report a case of STEMI with COVID-19 and suggest that respiratory failure with hypoxia can lead to myocardial injury. The myocardium is metabolically flexible (Pascuale and Coleman, 2016) although the association between hypoxia and cardiac arrest is well characterised (Parish et al, 2018). (Raad et al, 2020) investigated the pattern of cardiac injury in patients with COVID-19 and in their discussion suggest a number of factors that may lead to myocardial infarct type II in COVID-19 including hypoxia. Myocardial infarct can lead to complications including Atrial Fibrillation (Gorenek and Kudaiberdieva, 2012) which in turn predisposes to thromboembolism.

Hypoxaemia which can lead to hypoxia is a characterising feature of ARDS and many of the 56 main papers identified in our review provide examples of patients with COVID-9 and ARDS. (Raad et al, 2020) therefore provide indirect evidence of COVID-19 leading to hypoxia which in turn leads to myocardial damage as they identify evidence of an association between ARDS and myocardial injury expressed as high-sensitivity Troponin T levels.

In summary, there is strong evidence of hypoxaemia in COVID-19 and there is evidence of ARDS-associated myocardial injury in COVID-19 as well as established evidence on the relationship of hypoxia to myocardial injury. While strictly speaking the initial ischaemic injury does not result from a coagulopathy (e.g. NMDA-mediated excitotoxic cell death), coagulation disturbances occur secondary to hypoxia and tissue injury.

###### c. Hypoxaemia/hypoxia with vasoconstriction

Many of the authors in the main paper comment on the significance of hypoxia in relation to the COVID-19 related coagulopathy. (Jain et al, 2020) describe cases of hypoxia in their retrospective cohort study. (Meza et al, 2020) suggest hypoxia as one of many putative mediators of stroke in COVID-19. (Helms et al, 2020) suggest hypoxia as a mediator of increased stroke risk and provide evidence of the increased thrombosis seen in ARDS. ARDS is one of the most relevant examples of a COVID-19-related hypoxaemic state although the multitude of pathologies in the critically ill patient can complicate the interpretation. (Helms et al, 2020) suggest a number of mechanisms by which hypoxia can lead to thrombosis. Hypoxia can lead to vasoconstriction which predisposes to occlusion and they cite (Grimmer and Kuebler, 2017) who outlined the details of hypoxic pulmonary vasoconstriction (HPV). HPV is an efficient homeostatic mechanism which ensures oxygenated areas of the lung are well perfused. HPV is influenced by a number of pharmacological agents which are relevant in anaesthetics (Tarry and Powell, 2017). However as a homeostatic mechanism, this would divert blood away from hypoxic areas and would therefore not be relevant to the coagulopathy unless the mechanism is dysregulated.

Hypoxia-inducible factors (HIF’s) are transcription factors which alter gene expression including Tissue Factor and Plasminogen-activator inhibitor-1 (PAI-1). (Evans, 2019) suggests HIF’s as a mediator between sepsis and thrombosis. (Gomez-Arbeleaz et al, 2020) suggest a number of mechanisms for the increased risk of thrombosis in COVID-19 including increased viscosity of the blood secondary to hypoxia (sic).

In summary, there is a robust evidence base for hypoxaemia/hypoxia in COVID-19, particularly as a result of ARDS. There is also a robust evidence base for a causal link with coagulopathy.

###### a. Microangiopathy leading to ischaemia

(Fara et al, 2020) refer to a microangiopathy that may be associated with SARS-COV2 infection in their case series of macrothrombosis and stroke.

(Makatsariya et al, 2020) provide a useful overview of microangiopathy. They note that thrombotic microangiopathy is divided into primary and secondary. There are two main forms of primary thrombotic microangiopathy: Haemolytic uraemic syndrome (HUS) and thrombotic thrombocytopenic purpura (TTP). In TTP there is reduced activity of ADAMTS13, an enzyme which cleaves the procoagulant von Willebrand Factor (VWF) while in HUS there is excessive release of VWF into the circulation. Secondary thrombotic microangiopathies also exist and result from a number of causes including autoimmune disorders.

(Martinelli et al, 2020) reported on a retrospective study of 50 patients with COVID-19 and find evidence of a mild deficiency in ADAMTS13 suggesting that this is evidence of a secondary microangiopathy. They also found evidence of an inverse relationship with D-Dimer levels. (Baeck et al, 2020) investigate the possibility that chilblains reported in COVID-19 may result from a microangiopathy and conclude that there is no relationship. (Jhaveri et al, 2020) report a case of COVID-19 in which there is thrombotic microangiopathy together with alternative complement activation.

(Song and FitzGerald, 2020) suggest that microangiopathy in COVID-19 may be mediated by dysregulation of complement activation and draw comparisons with atypical haemolytic anaemia and paroxysmal nocturnal haemoglobinuria. (Ackermann et al, 2020) report on a case series of 7 autopsies in which they found widespread evidence of thrombosis with microangiopathy in the pulmonary vessels. They further identified vascular angiogenesis as a feature of COVID-19 which distinguishes it from severe influenza. (Gavriilaki and Brodsky, 2020) provide evidence of microangiopathy in COVID-19 and outline the pathological mechanisms.

In summary, there are multiple lines of evidence of microangiopathy in COVID-19 and of an association with alternative complement activation. However there is also evidence that this can occur without the more characteristic features of COVID-19-related coagulopathy where there are elevated D-Dimer levels.

###### b. Endotheliitis

A number of the authors in the main papers refer to either the possibility of endotheliitis or provide evidence of the same. (Escalard et al, 2020) in their case series of mechanical thrombectomy for ischaemic stroke suggest that endotheliitis may be a reason why reperfusion was not achieved despite recanalisation. (Varga et al, 2020) in their case series report evidence of direct viral invasion of the endothelium and widespread endothelial inflammation. They cite evidence of widespread expression of ACE-2 receptors on the endothelium as a facilitator of viral entry into endothelial cells. (Fara et al, 2020) in their case series note thrombosis in association with mild COVID-19 and discuss the possibility of endotheliitis and reference another paper providing evidence of this (Sardu et al, 2020). (Sardu et al, 2020) examine the evidence for endothelial involvement in COVID-19 although focusing on endothelial dysfunction.

(Reitsma et al, 2007) provide an overview of the endothelial glycocalyx noting that it is membrane-bound layer consisting of proteoglycans (predominantly), glycosaminoglycans and glycoproteins in dynamic equilibrium with soluble glycosaminoglycans and proteins in the plasma. Important components of the glycocalyx are hyuluronan and heparin sulfate. Hyuluronan has a significant ability to bind to water molecules and various clinical applications including as a dermal filler. (Semeraro and Colucci, 2000-2013) outline the evidence of anticoagulant properties of the endothelium involving the protein C pathway, Tissue Factor pathway inhibitor and heparin sulfate and other glycosaminoglycans (GAGS).

An example of how endothelial cells and the glycocalyx in particular are specialised is provided by (Sol et al, 2020). In their review they outline the contribution of the endothelium and the glycocalyx in particular to the glomerular filtration barrier (GFB) in the kidneys. Furthermore they suggest that the glycocalyx can be disrupted by processes such as inflammation and by such means can contribute to the development of focal segmental glomerulosclerosis (FSGS). Interestingly this is of particular significance in COVID-19.

(Goshua et al, 2020) provide evidence of endotheliopathy in COVID-19 with elevated markers, find a difference between critically ill and non-critically ill patients with COVID-19 and provide evidence of soluble thrombomodulin as a prognostic indicator. (Tong et al, 2020) found evidence of increased levels of serum endothelial cell adhesion molecules in severe COVID-19 which decreased with recovery.

(Yamaoka-Tojo, 2020) suggests endothelial glycocalyx damage as a mediator of systemic inflammatory microvascular endotheliopathy in COVID-19 and relates this to the clinical findings in COVID-19. (Fraser et al, 2020) found evidence of glycocalyx degradation in critically ill patients with COVID-19. (Cipolloni et al, 2020) from 2 autopsies found evidence of Factor VIII positivity and endothelial and alveolar damage which they suggest leads to different forms of ARDS. (Nikbakht et al, 2020) note the presence of endothelial cells in the blood-brain barrier and suggest that this is relevant to CNS involvement in COVID-19. In summary, (Varga et al, 2020) provide evidence of endotheliitis in COVID-19 as well as a putative mechanism by which this is initiated and there is strong evidence both for a role of the endothelium in coagulation independently of COVID-19 and also for endotheliitis in COVID-19.

###### c. Endothelial Dysfunction

Many authors in the main papers suggest endothelial dysfunction as a mediator of COVID-19-related coagulopathy. Endothelial dysfunction is well-characterised and describes a reduction in the availability of vasodilators such as nitrous oxide and an increase in endothelial-derived vasoconstrictors resulting from the action of various cardiovascular risk factors. Endothelial dysfunction is a precursor for atherosclerosis and predisposes to a number of pathologies including thrombosis (Hadi et al, 2005).

With regards to COVID-19, (Varga et al, 2020) provide evidence of apoptosis in association with endothelial dysfunction in their case series. As per the previous section (Sardu et al, 2020) examine the evidence for endothelial dysfunction in COVID-19. (Smadja et al, 2020) identified elevated Angiopoetin-2 and E-selectin on admission as prognostic for ICU admission in patients with COVID-19. They note that both are markers for endothelial dysfunction. Further they suggest that an intact endothelium is antithrombic and disruption of the endothelium is therefore prothrombotic and that an analogy can be drawn with pre-eclampsia.

In summary, there is evidence for a role for endothelial dysfunction in increasing the risk of COVID-19-related coagulopathy although we did not identify evidence to suggest that this results from COVID-19. We therefore treat this as a risk factor but not a COVID-19-mediated mechanism for coagulopathy. The evidence base for endothelial dysfunction independent of COVID-19 is well established and so we consider this as a vulnerability factor in COVID-19-related coagulopathy although the evidence base in COVID-19 is well developed in terms of prognosis rather than coagulopathy.

###### d. Vasculitis or vasculitis-like mechanism

(Morassi et al, 2020) in their case series of stroke with COVID-19 suggest vasculitis as a possible mediating mechanism. They cite research in both SARS and New Haven Coronavirus. In SARS there was evidence of fibrinoid necrosis and inflammatory cells in the vessel walls in three cases (Ding et al, 2003).

The relationship between vasculitis and thrombosis is well-established and (Emmi et al, 2015) provide an overview of the evidence for increased risk of thrombosis in a number of vasculitides. (Iba et al, 2020) suggest macrophage activating syndrome as a cause of coagulopathy in COVID-19-related vasculitis. (Tahir et al, 2020) report a case of cutaneous small vessel vasculitis (CSVV) secondary to COVID-19 after exclusion of other differentials. (Hussein et al, 2020) report a case of COVID-19 with pulmonary haemmorhage secondary to ANCA vasculitis although the association with COVID-19 was unclear.

(Becker, 2020) reviews the evidence for vasculitis in COVID-19 and provides an overview for the pathogenesis of vasculitis. (Becker, 2020) references the histopathological findings of endotheliitis.

In summary, there are multiple lines of evidence for vasculitis in COVID-19, the relationship with COVID-19 and also the relationship of vasculitis to thrombosis independent of COVID-19. As per (Becker, 2020) we will group vasculitis and endotheliitis together.

###### d. SARS-COV-2-induced small vessel thrombosis

(Kaafareni et al, 2020) suggest two mechanisms in particular that would merit further investigation one of which is the possibility of a SARS-COV2-induced small vessel thrombosis. They cite (Connors and Levy, 2020) who write about thromboinflammation and who in turn reference (Fox et al, 2020) whose autopsy findings include small vessel thrombosis. There are numerous studies identifying evidence of small-vessel thrombosis in COVID-19 and this appears to be a robust finding (Bösmüller H et al, 2020), (Fox et al, 2020)(Ackermann et al, 2020)(Lax et al, 2020). Many of these findings describe pathology in the lungs. (Wang et al, 2020) report two cases of COVID-19 with digital gangrene mediated by small-vessel thrombosis.

In summary, the association with coagulopathy is a defining feature of small vessel thrombosis and there are many lines of evidence to support the occurrence of small vessel thrombosis in COVID-19 particularly in the lungs. However we would include small vessel thrombosis under the heading of microangiopathy with thrombosis.

##### E. Iatrogenic

###### 1. Pharmacological adverse events

(Kaafareni et al, 2020) describe a case series of patients with COVID-19 and gastrointestinal symptoms and suggest a number of explanatory mechanisms including pharmacological side-effects although this could also cover non-coagulopathy-related clinical features. Nevertheless there are a number of pharmacologically-active agents with thrombophilic properties and this includes licensed medications and illegal drugs. (Girolami et al, 2016) have reviewed the subject and comment on the paucity of published literature. However they identify 21 classes of pharmacologically active agents with possible thrombophilic properties. The licensed medications include the antipsychotic Clozapine, oral contraceptives, Tamoxifen and Heparin which can lead to Heparin-induced Thrombocytopenia (HIT). They also cite a number of case reports of arterial thrombosis with the use of Marijuana. The possibility that there may be a synergistic thrombophilic effect involving thrombophilic pharmaceutical agents and COVID-19-specific thrombophilic mechanisms should be considered. An association between severe COVID-19 and polypharmacy was found in one pre-print paper (McKeigue et al, 2020). Osteonecrosis of the femoral head was reported in SARS and has been attributed to the use of steroids but not yet described in COVID-19 on the basis of our examination of the literature (Tang et al, 2020). In summary, there are a number of pharmacologically-active agents that are procoagulant and this evidence base is well developed independently of COVID-19. On the basis of our literature search we did not identify evidence of a causal relationship with COVID-19-related coagulopathy but that may be a reflection of the limits of the search and published evidence

###### 2. Mechanical Ventilation

Pulmonary coagulopathy occurs secondary to lung injury. The aetiology of lung injury is broad and includes mechanical ventilation. Indeed the treatment of mechanical ventilation-associated-pulmonary coagulopathy was evaluated in a meta-analysis (Glas et al, 2016). In summary, mechanical ventilation is used in a subset of patients in COVID-19 and there is an established literature on mechanical ventilation-associated coagulopathy.

##### F. Immune-Mediated Syndromes

###### 1. Antiphospholipid syndrome

(Zayet et al, 2020) reported elevated anticardiolipin antibodies in one case whilst noting that a diagnosis of antiphospholipid syndrome cannot be made unless positive antibodies have been present for several months. This has presented a challenge in the early stages of the pandemic.

There are a number of suggested mechanisms by which antiphospholipid antibodies can potentially mediate coagulopathy. (Salmon and de Groot, 2008) discuss a number of these mechanisms in their paper including inhibition of Proteins C activity, inhibition of Protein S activity, inhibition of antithrombin activity and Tissue Factor induction.

(Devreese et al, 2020) in their case series of 31 patients in ICU identified positive antiphospholipid antibodies in 10 patients at initial testing but these tests were negative at repeat testing one month later. (Xiao et al, 2020) report antiphospholipid antibodies in 47% of the critically ill patients in their study and arising at a median of 39 days after disease onset. Those with multiple antibodies experienced a higher incidence of ischaemic stroke. (Cavelli et al, 2020) argue that secondary antiphospholipid syndrome is the cause of the coagulopathy in COVID-19.

(Borghi et al, 2020) demonstrate a low prevalence of antiphospholipid antibodies in their study involving 122 patients with severe COVID-19. (Zuo et al, 2020) found evidence of antiphospholipid antibodies in their series of 172 patients with COVID-19 and correlated higher levels with the severity of respiratory disease and increased neutrophil activity with release of neutrophil extracellular traps. (Rodriguez et al, 2020) note that as well as antiphospholipid syndrome, other autoimmune conditions have been reported in COVID-19. (Iba et al, 2020) outline 5 different coagulopathies associated with COVID-19 including an antiphospholipid antibody syndrome-like coagulopathy amongst four other coagulopathies.

(Maria et al, 2020) report a case of flare-up of primary antiphospholipid syndrome with adrenal haemmorhage and acute limb ischaemia in a male patient with COVID-19 and suggest an interaction between COVID-19 and antiphospholipid syndrome. (Cavalli et al, 2020) note that antiphospholipid antibody syndrome can result from viral infections.

In summary, there is evidence of antiphospholipid antibodies during the acute phase of COVID-19 but follow-up studies from those that we identified the evidence supports an association with elevated antiphospholipid antibodies but this is not persistent and appears to be distinct from antiphospholipid syndrome. Nevertheless there may be a role for these antibodies in the pathogenesis of COVID19 in a unique way. We therefore distinguish between antiphospholipid antibody syndrome-like coagulopathy for which there is evidence but not for the antiphospholipid antibody syndrome.

###### 2. Type-1 Interferonopathy

(Bouaziz et al, 2020) noted similarities between the dermatological presentation of COVID-19 and type-1 interferonopathy. Chilblains for instance occur in Aicardi-Goutiéres syndrome (AGS) (Crow, 2016).

This hypothesis is further developed by (Günther et al, 2020) and (Damsky et al, 2020). (Damsky et al, 2020) suggest that sporadic pernio in COVID-19 reflects a strong host interferon-I response and that patients experiencing acral pernio or chilblains are experiencing a milder course of illness due to the immune response. Histopathological findings were presented in (Kolivras et al, 2020) as well as a hypothesis about the interferon-1 response.

(Muskardin and Niewold, 2018) review the role of type-1 interferons in rheumatic diseases including systemic lupus erythematosus (SLE) and also note 12 different type-1 interferons. The type-1 interferons contrast with the type-III interferons which are preferentially activated in the respiratory epithelium and other barrier surfaces (Wells and Coyne, 2018). Interestingly there is evidence to suggest that type-1 interferon can impair the response to secondary bacterial infections (Shaabani et al, 2018). (Nagafuchi et al, 2019) suggest the need for precision medicine in profiling the immune response in SLE. (Chen et al, 2020) review the relationship of type-1 interferons to outcomes with atherosclerosis and note a role in endothelial dysfunction. (Martin-Fernandez et al, 2020) identified type-1 interferon-induced necrotic skin lesions in their study of five families with a deficiency in ISG15.

(Rodero and Crow, 2016) review the type-I interferonopathies and outline the different groups of clinical features (see Table 31) as well as the associated genotypes (see Table 32) and their posited role. They suggest that interferon is potent in action but also difficult to detect. In their review they state that there is insufficient evidence to support a causal relationship between an upregulation of type-I interferon signalling and the associated clinical features at that time.

**Table 31.** Presentations of Type-I Interferonopathies as per (Rodero and Crow, 2016)

|  |
| --- |
| Lupus |
| Spastic paraparesis |
| Lung inflammation |
| Developmental Delay |
| Malignancy |
| Glaucoma |
| Skin vasculopathy |

**Table 32.**
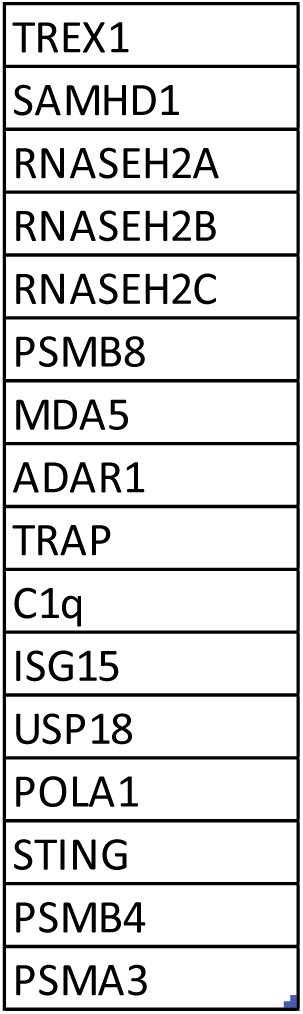
Genotypes of Type I Interferonopathies as per (Rodero and Crow, 2016)

|  |
| --- |
| TREX1 |
| SAMHD1 |
| RNASEH2A |
| RNASEH2B |
| RNASEH2C |
| PSMB8 |
| MDA5 |
| ADAR1 |
| TRAP |
| C1q |
| ISG15 |
| USP18 |
| POLA1 |
| STING |
| PSMB4 |
| PSMA3 |

(Schreiber, 2017) divides type-1 interferon signalling into robust and tunable and outline the details of the signalling cascade.

(Xia et al, 2020) found three SARS-COV2 proteins that functioned to evade the host’s type1 interferon response while (Gemcioglu et al, 2020) report a mild progression of COVID-19 in a patient treated with type 1 interferon for multiple sclerosis. Another perspective on interferon and COVID-19 is found in interventions. (Yu et al, 2020) in their systematic review and meta-analysis note that although there is a role for interferons for first-line therapy in coronavirus infections within local protocols there is also a need for robust trials.

In summary, there is evidence of a similarity between skin lesions in type-1 interferonopathies and COVID-19 and clinicians have hypothesised that a milder form of COVID-19 is experienced where there is an acute type-1 interferon response although offering a possible explanation for a range of skin lesions identified in COVID-19. While the biology of the interferonopathies is detailed, some of the authors have commented on the need for more evidence for causality in terms of clinical associations with the interferonopathies. The evidence for the involvement of type-1 interferons in COVID-19 is tentative but interestingly links in with other perspectives.

##### G. General Factors

###### 1. Critical illness encephalopathy

(Deliwala et al, 2020) note the association of critical illness encephalopathy, cytokines and stroke in their case report and literature review and suggest that this may represent a precursor state. (Aggarwal et al, 2020) investigate the relationship between pre-existing cerebrovascular disease and severity of COVID-19. (Helms et al, 2020) provide evidence of an association between severe COVID-19 with ARDS, critical illness encephalopathy and stroke although noting that they have not demonstrated a causal relationship. (Deliwala et al, 2020) present a case of encephalopathy associated with ARDS where the imaging does not initially reveal evidence of ischaemic stroke although this is subsequently identified. The relationship between encephalopathy and delirium has been outlined by (Slooter et al, 2020) who describe acute encephalopathy as a pathological brain process which may lead to a number of clinical conditions including delirium although having other associations also. In summary, we interpret this as a COVID-19-induced encephalopathy that may lead to an increased risk of ischaemic stroke although the evidence is limited and one of association rather than causality. The broader concept of a critical-illness encephalopathy has been developed in the literature and is a move to organise the findings in cases which may be complex and multifaceted. In such cases, other mechanisms including those discussed in this paper may be playing a role.

###### 2. Hypotension

(Zhou et al, 2020) in their case report of DVT and acute limb ischaemia note that hypotension is a risk factor for DVT. (Momtazmanesh et al, 2020) did not report hypotension as a finding in their systematic review and meta-analysis of 10,898 patients with COVID-19. (Wu et al, 2020) and (Song et al, 2020) identify a significant role for the order in which vasopressors are discontinued in the treatment of septic shock. Hypotension is however a cardinal feature of septic shock (Evans, 2018) and so this is best considered under the heading of septic shock. Additionally hypotension is a recognised cause of ischaemia (McDowall, 1985).

###### 3. Hypertension

(Yaghi et al, 2020) suggest relative hypertension secondary to mechanical ventilation leading to posterior reversible encephalopathy syndrome as a mechanism for ischaemic stroke in COVID-19. (Momtazmanesh et al, 2020) in their systematic review and meta-analysis note the significance of pre-existing hypertension as a risk factor for prognosis. (Jain and Yuan, 2020) in their systematic review and meta-analysis similarly identify hypertension as a comorbidity which was predictive for severity of COVID-19. In summary we will group this with mechanical ventilation associated mechanisms and also recognise hypertension as a risk factor for COVID-19 severity and suggest that this may relate to endothelial dysfunction and is best considered under the heading of vulnerabilities.

###### 4. Lymphopenia and Leucocytosis

(Zayet et al, 2020) describe lymphopenia and leukopenia in their cases of COVID-19 with stroke and relate this to the associated stroke risk. They cite a study demonstrating that the lymphocyte-to-monocyte ratio is a prognostic indicator in acute ischaemic stroke (Ren et al, 2017). (Urra et al, 2017) found an association between lymphopenia and ischaemic strokes involving the superior and middle temporal gyri. The phenomenon of post-stroke immune depression has been described (Liesz et al, 2013) but this temporal relationship would therefore not account for initial strokes in COVID-19. Nevertheless both post-stroke immune depression and infection (Klehmet et al, 2009) may be important in the characterisation of the consequences of COVID-19 coagulopathy in cases of stroke particularly in terms of the long term consequences of COVID-19 (Heneka et al, 2020). In a meta-analysis (Huang et al, 2020) found higher leukocyte counts and lower lymphocyte counts in patients with severe COVID-19 compared to patients with mild illness. In summary there is evidence of lymphopenia and leucocytosis in severe COVID-19 and there is an association between lymphopenia and stroke risk. In terms of modelling, this is best considered in terms of the thrombophilic state.

###### 5. Atrial Fibrillation

(Arrigo et al, 2020) in their review of Atrial Fibrillation note that this is a common condition in an ICU setting and they identify several mechanisms leading to atrial fibrillation: myocardial stretch, inappropriate oxygen delivery, electrolyte disturbances, inflammation, adrenergic overstimulation, endocrine disorders and hypothermia. They outline a number of treatment approaches depending on the underlying mechanism. (Lazzeri et al, 2020) report a series of 28 patients with COVID-19 and ARDS. Two of the patients presented with chronic atrial fibrillation while 11 of the remaining 26 presented with paroxysmal atrial fibrillation. (Holt et al, 2020) report a reduction in the incidence of atrial fibrillation during the early period of lockdown but suggest that this may be due to the effects of the pandemic on healthcare access. (Schnaubelt et al, 2020) report on a case of atrial fibrillation complicating a suspected cytokine storm. In summary there is evidence of atrial fibrillation in patients with COVID-19 albeit in a critical care setting where the prevalence of atrial fibrillation is expected to be increased due to a range of factors. Thus atrial fibrillation may be a non-specific feature of COVID-19 relating to the context of a critically unwell patient and this would not necessarily be restricted to the ICU setting given the rapid deterioration that is seen. There is a strong evidence base for atrial fibrillation as a cause of thromboembolic events which forms the basis for the practice of thromboprophylaxis.

###### 6. Immobilisation and Bed rest

Immobility and bed rest are well recognised risk factors for DVT and this relationship is an important contributor to the practice of thrombophrophylaxis in the hospital setting. (McLendon et al, 2020), (Zhou et al, 2020) and (Prevatali et al, 2011) review the risk factors for DVT (see Table 33) and cite evidence for immobility and bed rest as risk factors for DVT. In the 56 main papers, the authors note that patients experienced thromboembolic events despite thromboprophylaxis. There is no doubt that this is an important consideration in the ICU setting. (McLendon et al, 2020) note a hierarchy of DVT risk with immobility in order from highest to lowest: Hip, knee, ankle, shoulder, elbow.

**Table 33.** Deep Vein Thrombosis Risk Factors (McLendon et al, 2020)(Zhou et al, 2020)(Prevatali et al, 2011)

|  |
| --- |
| Age |
| Bed rest |
| Respiratory failure |
| Acute coronary syndrome |
| Congestive heart failure |
| Chronic kidney disease |
| Chronic disease |
| Indwelling catheters |
| Long distance travel |
| Major trauma |
| Non-infectious inflammatory conditions |
| Obesity |
| Varicose vein in lower extremity |
| Prior Venous Thromboembolism |
| Smoking |
| Stroke |
| Thrombophilias |
| Surgery |
| Immobility |
| Oestrogen use |
| Pregnancy |
| Initial postpartum |
| Family History |
| Infection |
| Impaired blood flow |
| Malignant tumour |
| Chemotherapy |
| Air Pollution |

The veins are valved which influences the dynamics of blood flow and periods of stasis can predispose to thrombosis. However the arterial anatomy is substantially different and in our findings the arterial ischaemic events significantly outnumbered those affecting the veins. (Reddy et al, 2016) describe a case of arterial ischaemia in the lower limb secondary to thrombosis involving both the deep and superficial veins - phlegmasia cerulean dolens. This mechanism may be of minor relevance in COVID-19 but in the majority of the 56 main papers describing acute limb ischaemia, deep vein thrombosis was not reported as a comborbidity.(Zhou et al, 2020) was the exception.

Air travel presents a special case of immobility and the occurrence of deep vein thrombosis has been well documented in this setting and the efficacy of interventions investigated (Clarke et al, 2016). (Sohail and Fischer, 2005) reiterate the finding of deep vein thrombosis in their review of air travel and health-related conditions but also cite cases of arterial thrombi although less frequent.

In summary, bed rest and immobility are associated with deep vein thrombosis independent of COVID-19 and this relationship is well-established. This association may result from the anatomy of the veins, the dynamics of blood flow in the returning venous circulation and the effects of gravity in pooling blood. Whilst arterial thrombi are described in the literature in association with immobility this occurs much less commonly. In COVID-19 particularly in the ICU setting, immobilisation and bed rest, combined with a thrombophilic state may predispose to DVT’s even with thromboprophylaxis and the smooth muscles cells in the walls of the veins express ACE-2 receptors.

###### 7. Dehydration due to fever, diarrhoea

(Zhou et al, 2020) suggest fever and diarrhoea as relevant risk factors for DVT in COVID-19. In a retrospective study of 25 patients with COVID-19 diarrhoea was reported in 8% of patients (Duan et al, 2020). (Parasa et al, 2020) in a systematic review and meta-analysis which looked at the prevalence of gastrointestinal symptoms found that 12% of patients with SARS-COV-2 infection experienced gastrointestinal symptoms including diarrhoea, nausea and vomiting. In another systematic review and meta-analysis looking at gastrointestinal involvement in COVID-19, the prevalence of diarrhoea was 10.4% (95% CI 7.7-13.9) (Rokkas, 2020). (Hajifathalian et al, 2020) identified a prevalence for diarrhoea of between 10 and 29% in patients with COVID-19 in their review of the literature. In another systematic review and meta-analysis, the authors looked at the outcomes in 61, 742 patients. In a subset of 18 studies they found a prevalence of diarrhoea of 8% (95% confidence interval 4.6-11.4) (Pormohammad et al, 2020). (Kaur et al, 2020) found a prevalence of diarrhoea of 9.2% in their systematic review.

In terms of the relationship between diarrhoea and dehydration, this is well-established and the importance of oral rehydration has been emphasised across a range of settings. (Giddings et al, 2016) provide an overview of traveller’s diarrhoea including the central role of oral rehydration therapy in addressing dehydration. Dehydration can increase the likelihood of coagulation. (Shi et al, 2019) found that dehydration mediated ischaemic stroke risk via activation of coagulation. (Borgman et al, 2018) demonstrated that dehydration leads to central hypovolaemia with increased clotting in a trial involving 11 volunteers. (Li et al, 2017) identified dehydration as a predictor for prognosis in ischaemic stroke over the long term.

Fever has been well characterised in COVID-19 as per the introduction and can lead to fluid loss through sweating thereby providing an additional mechanism which can lead to dehydration.

In summary there is strong evidence both for fever and diarrhoea in COVID-19 and this can occur in mild as well as severe presentations of COVID-19. Fever and diarrhoea can lead to dehydration for which there is robust evidence for a causal link with coagulopathy.

###### 8. Metabolic/electrolyte disturbance

(Kaafareni et al, 2020) in their description of gastrointestinal complications of COVID-19 including mesenteric ischaemia suggest a number of mechanisms including metabolic and electrolyte disturbances seen in critically ill patients. Numerous metabolic and electrolyte disturbances have been described in critically ill patients and may have a relationship with thromboembolic events.

In more general terms (Gawalko et al, 2020) suggest a number of mediators of atrial fibrillation including hypoxaemia, electrolyte imbalance, endothelial dysfunction, ACE-2-related pathways and the cytokine storm. Renal function is an important consideration for electrolyte disturbances and there are a number of findings on urea nitrogen levels (BUN) where BUN is approximately twice the value of serum urea levels. (Shao et al, 2020) undertook a meta-analysis (n=24527) of acute kidney injury in patients with COVID-19 and found that acute kidney injury was significantly associated with mortality and BUN and creatinine were significantly associated with mortality. (Post et al, 2020) report two cases of kidney infarcts associated with elevated LDH. (Liu et al, 2020) found that urea nitrogen (BUN) levels, D-Dimers and lymphocyte ratio (expressed as a percentage of white cell count) were prognostic in COVID-19. (Oussaleh et al, 2020) analysed the biochemical parameters in a prospective cohort study of 162 patients with severe COVID-19 and found that only urea Nitrogen > 0.42 g/L was prognostic for death. Thus there may be an indirect relationship with coagulopathy via critical illness and the associated risks.

In terms of sodium (Corwin and McIlwaine, 2019) note that in critically ill patients, hypernatraemia may be associated with reduced cerebral volume which can lead to mechanical stress on the cerebral blood vessels. However (Berni et al, 2020) found evidence of hyponatraemia in COVID-19 and that it was inversely correlated with IL-6 levels which they suggest is mediated by an IL-6-induced vasopressin release. (Post et al, 2020) cite evidence that low sodium levels may cause upregulation of the renal membrane bound ACE-2 receptors. In turn they suggest that this would increase the risk of SARS-COV2 binding to renal ACE-2 receptors. They also cite evidence of an association between hyponatraemia and severity of COVID-19 (Libbi et al, 2020). In another paper (Post et al, 2020) also cite evidence of geographical differences in sodium intake and suggest that dietary intake may play a role in illness severity. (Królicka et al, 2020) reviewed the evidence for hyponatraemia in infections including COVID-19. They cite evidence for hyponatraemia in COVID-19 and more generally identify hyponatraemia as a risk factor in infections for longer hospital stay and mortality. (Ghahramani et al, 2020) in their meta-analysis of 22 studies (n=3396) found evidence of a significant association of decreased sodium in patients with severe COVID-19 compared to non-severe COVID-19. Hyponatraemia is also noted in patients with stroke and suggested mediators include the syndrome of inappropriate ADH secretion and cerebral salt wasting syndrome (Saleem et al, 2014). Hyponatraemia occurred in 30% of patients in a retrospective observational cohort study (Frontera et al, 2020) with increasing severity correlated with mortality, encephalopathy and mechanical ventilation with lower levels being correlated with IL-6 levels.

(Liu et al, 2020) found evidence of hyponatraemia in 25.8% of 93 patients with COVID-19. (Tezcan et al, 2020) found that hyponatraemia was the most common electrolyte disturbance in their study of 408 hospitalised patients with COVID-19. In their study, hyponatraemia, hypochloraemia and hypocalcaemia were prognostic for outcomes. (Zimmer et al, 2020) in contrast with other studies found evidence of hypernatraemia in COVID-19 and this was associated with prognosis as well as hypokalaemia and hyperchloraemia. They suggest that this results from elevated angiotensin II levels. (Suso et al, 2020) report a case of IgA Vasculitis with Nephritis. (Carriazo et al, 2020) draw attention to the occurrence of SIADH in COVID-19. (Khan et al, 2020) present a case of hypovolaemic hyponatraemia in COVID-19. (Ata et al, 2020) present a case of hyponatraemia in COVID-19 and suspect that this reflects SIADH. (Kumar et al, 2020) present a case of hyponatraemia with adrenal infarction in COVID-19. (Habib et al, 2020) report a case of symptomatic hyponatraemia in COVID-19 which was determined to be secondary to SIADH after further investigation. (Ravioli et al, 2020) report 2 cases of SIADH in COVID-19. Thus there are consistent findings with the vast majority of studies identifying hyponatraemia but when hyponatraemia or hypernatraemia occur they are both prognostic in COVID-19. A key point here is that if sodium homeostasis is working effectively then hyponatraemia or hypernatraemia would be avoided and it is likely that this is related to a disruption in the RAA system.

(Frise and Salmon, 2019) note the relationship between hypokalaemia and cardiac arrythmias and these in turn may give rise to thromboembolic events (e.g. atrial fibrillation). (Chen et al, 2020) completed a cohort study looking at hypokalaemia in COVID-19 (n=175). They found that 85% of severely and critically ill patients had hypokalaemia and that patients with higher levels of hypokalaemia were more likely to have elevated creatine kinase, lactate dehydrogenase, C-reactive protein levels and higher body temperature. They suggest that hypokalaemia is difficult to correct because of ACE-2 degradation and that the hypokalaemia is mediated via the renin-angiotensin system. (Vena et al, 2020) in a retrospective cohort study in a tertiary care hospital (n=317) identified hypokalaemia in 25.8% of patients. (Yang et al, 2020) found a significant relationship between corticosteroid treatment and hypokalaemia in patients with COVID-19 in their meta-analysis of 15 studies (n= 5270). (Chen et al, 2020) found a high prevalence of hypokalaemia in COVID-19 which was responsive to potassium supplementation. They suggest that this may result from disruption of the renin-aldosterone system. (Moreno-P et al, 2020) found evidence of hypokalaemia in 30.7% of 301 patients with COVID-19 and this was associated with need for mechanical ventilation.

(Prabhu et al, 2020) present a case of BRASH syndrome (bradycardia, renal failure, AV nodal blockade, shock, and hyperkalemia) in COVID-19. (Chen et al, 2020) looked at the characteristics of 113 deceased patients compared to those who survived in their study (n=799) and found that renal failure, hyperkalaemia and alkalosis were more common in those who died. Thus in the case of potassium, there are mixed results. With the RAA system, sodium retention is accompanied by potassium excretion whereas hypokalaemia and hyponatraemia are commonly reported in studies. Taken together with the evidence about the RAA system, the most plausible explanation is that there is a disruption of sodium and potassium regulation due to a disruption in the RAA system.

(Rosengart, 2019) outlines the consequences of hypocalcaemia and hypercalcaemia in critically ill patients and the association of hypocalcaemia with hypotension is noted. Hypotension is independently associated with stroke risk (Eigenbrodt et al, 2000). The role of Vitamin D in the immune system is well documented (Charoenngam and Holick, 2020) and there is also a well-established causal link with hypocalcaemia in Vitamin D deficiency in keeping with the homeostatic functions of Vitamin D. (Chakhtoura et al, 2020) highlight the importance of Vitamin D in infections and suggest that Vitamin D may play a role in COVID-19 susceptibility.

(Bossoni et al, 2020) report a case of hypoparathyroidism in a lady with a thyroidectomy who developed COVID-19 and the authors suggest that the COVID-19 was the precipitant. (Elkattawy et al, 2020) report a case of hypoparathyroidism with normal calcium and hyperphosphataemia which they suggest as secondary to SARS-COV2 infection. (Liu et al, 2020) found evidence of hypocalcaemia in 62.6% of 67 patients with severe COVID-19 and was inversely correlated with D-Dimer and IL-6 levels. (Sun et al, 2020) found evidence that hypocalcaemia was prognostic in COVID-19 and associated with a higher incidence of septic shock and that calcium levels were correlated with Vitamin D levels.

(Liu et al, 2020) found that calcium levels were lower in severe COVID-19 and that calcium levels negatively correlated with CRP, D-Dimer and IL-6 levels. In a retrospective cohort study (n=241), 74.7% of patients with COVID-19 were hypocalcaemic on admission (Sun et al, 2020). Furthermore calcium was lower in those with severe illness and those with low calcium experienced higher 28-day mortality and were more likely to develop septic shock.

Another consideration for calcium homeostasis is medication. (Vila-Corcoles et al, 2020) found no significant reduction in the hazards ratio for developing COVID-19 in patients taking calcium channel blockers. There was a reduction for patients taking Angiotensin II receptor blockers but this was not statistically significant. Similarly (Sardu et al, 2020) found no significant effect of antihypertensive medication on prognosis in COVID-19 in their prospective cohort study. (Reynolds et al, 2020) looked at patients taking one of five classes of antihypertensives and found no significant reduction in incidence of COVID-19 or in severity in those with COVID-19. (Solaimanzadeh, 2020) did find a significant reduction in mortality as well as a significant reduction in intervention with mechanical ventilation and intubation in older adults admitted with COVID-19 who were receiving treatment with calcium channel blockers compared to those who were not.

(Rotman et al, 2020) report a case of a rare syndrome of calciphylaxis and COVID-19 in which calcium was deposited in the vasculature and associated in this case with ischaemic dermopathy. In retrospective cohort study in a tertiary-care hospital (Di Filippo et al, 2020) found hypocalcaemia in 82% of 531 patients with COVID-19. They also identified a correlation of hypocalcaemia with LDH levels and suggest a cytopathic viral lysis syndrome to account for this. Both (Kumar et al, 2020) and (Kumaran et al, 2020) report cases of pancreatitis with hypocalcaemia. (Demir et al, 2020) present a case of hypocalcaemia and hypoparathyroidism with COVID-19 presenting with Fahr’s syndrome.

Magnesium as a group 2 element in the periodic table, competes with calcium which is relevant physiologically. (Tang et al, 2020) review the role of magnesium in the management of different conditions and suggest a role for magnesium supplementation for specific indications in COVID-19.

(Kim et al, 2020) present two cases of acute hyperglycaemic crises in Diabetes in COVID-19, of which there are many other cases in the literature.

(Esen, 2019) outline the relationship between hypomagnesaemia and arrythmias in critically ill patients. Again this may similarly mediate thromboembolic events via atrial fibrillation. We did not identify evidence of hypomagnesaemia in COVID-19 in our literature search.

In summary there is evidence of hypocalcaemia, hyponatraemia and hypernatraemia, hypokalaemia and hyperkalaemia, hyperglycaemia as well as elevated BUN in COVID-19. There are also well-established links with hypotension, stroke and atrial fibrillation all of which are relevant to the central theme of COVID-19-related coagulopathy. For the purposes of modelling, this category can be included with the hypercoagulable/thrombophilic state.

###### 9. Low platelet count

(Meza et al, 2020) cite a number of factors which predispose towards a hypercoagulable state including low platelet count. One study which has been cited to support this assertion is (Liu et al, 2020). This paper references a low platelet count, relating this to the ACE-2 receptor and also to an increased risk of cerebral haemmorhage when combined with hypertension. This in turn relates to another paper (Chan et al, 2020). This paper described a family cluster of SARS-COV2 infection and is one of the earlier descriptions of the clinical features. (Levi and Opal, 2006) consider the low platelet count in the critical care setting and identify a number of mediators including disseminated intravascular coagulopathy, microangiopathy and sepsis.

Firstly looking at the relationship between platelets and thrombosis in the non-COVID-19 literature one of the conditions associated with both thrombocytopenia and thrombosis is heparin-induced thrombocytopenia (HIT). The relationship of heparin to thrombocytopenia is well-established and the experimental evidence dates back to 1942 (Arepally, 2017). The negatively charged heparin binds to the positively charged PF4 (which is released from platelet alpha-granules) forming a complex which due to the nature of the electrostatic interactions is dependent on the concentrations of heparin and PF4 (Arepally, 2017). The heparin-P4 complex is the target of antibodies (HIT). PF4 is expressed on the endothelium and is prothrombogenic. HIT is associated with thrombosis and more people develop antibodies when treated with heparin than develop thrombocytopenia or thrombosis (Arepally, 2017) suggesting a subclinical presentation also exists. The heparin/PF4/antibody complex binds to platelets and causes platelet activation via a spleen tyrosine kinase-mediated mechanism and procoagulant microparticles are released (Arepally, 2017). (Gollomp et al, 2018) provide experimental evidence to support several neutrophil-mediated prothrombotic mechanisms in HIT including formation of PF4/NET/HIT antibody complexes and neutrophil-binding to inflamed endothelium. (Perdomo et al, 2019) provide evidence that neutrophil activation and NET’s are predominantly responsible for the thrombosis in HIT. (Hsieh et al, 2013) report a case of pneumopathy associated with antiphospholipid antibodies, HIT and cerebral venous thrombosis. The absence of platelet alpha-granules is found in the grey platelet syndrome and is associated with a bleeding tendency (Gunay-Aygun et al, 2010).

Multivessel coronary artery thrombosis has been described in a case of idiopathic thrombocytopenic purpura (Yagmur et al, 2012). Antiphospholipid antibodies have been detected in primary immune thrombocytopenia even in the absence of antiphospholipid syndrome and thrombocytopenia is seen in primary antiphospholipid syndrome (Yang et al, 2011). (Katz et al, 2011) note an incidence of thrombocytopenia in intensive care settings of up to 50%.

They also note that thrombocytopenia occurs in up to 50% of cases of ARDS, is predictive of mortality in sepsis and septic shock, correlates with hypoxemic respiratory failure and is also seen in SARS. They suggest a range of causes may be responsible for the association of thrombocytopenia with critical illness including splenic sequestration.

(Donahue et al, 2010) reported on a novel approach to treating hypersplenism-associated thrombocytopenia in unresectable pancreatic cancer. They performed splenectomy and chemotherapy and they cite another study in which splenectomy enabled the continuation of interferon therapy in patients with hepatitis C, liver cirrhosis and portal hypertension (Kercher et al, 2004). DVT and pulmonary embolism occurred in a lady being treated for idiopathic thrombocytopenia purpura with intravenous immunoglobulin (Lee et al, 2007). (Hally et al, 2020) provide an overview of the platelet TOLL-like receptors including their response to PAMPS and DAMPS. They note that fibroblast stimulating lipopeptide-1 (FSL-1) is found to stimulate platelet production of IL-6 via the TLR2/6 receptors. (Li et al, 2020) note some of the ways in which platelets interact with microbes including sequestration.

(Hottz et al, 2020) identified platelet activation and platelet-mediated induction of Tissue Factor in monocytes in patients with severe COVID-19 compared to those with mild COVID-19. They provided evidence of mediation by integrin αIIb/β3 receptors and platelet p-selectin. (Manne et al, 2020) found evidence of increased p-selectin expression in platelets in people with COVID-19 compared to those without. There were found to be increased circulating aggregates of platelet-neutrophils, platelet-T-cells and platelet-monocytes in people with COVID-19. Platelet activation markers were not correlated with IL-6, IL-8, and TNF-α levels but demonstrated platelet hyperreactivity and there was evidence that this was related to both thromboxane generation and MAPK pathway activation.

(Becker et al, 2020) suggest that the megakaryocytes found circulating in the pulmonary microvasculature may be an important source of platelets in COVID-19. (Rapkiewicz et al, 2020) find evidence of platelet-rich thrombi in multiple organs and megakaryocytes.

In summary, there is an association between thrombocytopenia and thrombosis in specific conditions. However we would argue that confounding factors are responsible for the thrombosis with thrombocytopenia. Indeed the low platelet count is likely to reflect a period of increased activation of platelets and consumption which in turn is more likely to be related to thromboembolic events. The low platelet count is similar therefore to the D-Dimers and fibrinogen in reflecting the end result of pathway activation rather than the cause. The platelet count is included in the SIC score but here we are looking specifically at biologically plausible mechanisms for thromboembolic events. HIT is appropriate to consider and this can be included with iatrogenic mechanisms. More appropriate to a COVID-19 coagulopathy is the platelet activation that is central to the modern interpretation of thromboinflammation and it is in this context that we include platelet activation in the model of COVID-19-related coagulopathy.

###### 10. Plaques unstable due to mononuclear infiltrates, hypoxia, turbulence

(Cai et al, 2020) suggest that plaques may rupture in the context of COVID-19 associated hypoxia, haemodynamic turbulence and mononuclear infiltrates and thrombosis in microvessels. They cite two papers (Xu et al, 2020)(Restrepo et al, 2018). (Restrepo et al, 2018) provide evidence of 20 different cardiac-injury mechanisms resulting from organisms which cause pneumonia. This includes instability of coronary atheromas, acute coronary syndromes and thrombin formation. (Xu et al, 2020) report on post-mortem findings in a male patient with Covid-19 but found no evidence of cardiac pathology. Taken together these papers provide evidence of established mechanisms of cardiac injury by organisms that cause pneumonia but this evidence is not specific to COVID-19. We will consider this as best placed within the possible consequences of secondary infection in COVID-19 depending on the causative organisms.

##### H. Secondary Infection

###### 1. Septic embolisation with bacterial superinfection

(Yaghi et al, 2020) suggest a number of mechanisms that may mediate the increased risk of stroke seen in the cases they presented. They suggest that the critical care period has established associations including septic embolisation in the case of superinfections.

With reference to the non-COVID-19 literature, (Elsaghir and Khalil, 2020) review the evidence for septic embolisation with the most notable example being infective endocarditis. In an autopsy series (Grosse et al, 2020) found evidence of gram-negative and positive as well as fungal bronchopneumonia in cases of COVID-19. They also describe fungal colonisation with septic pulmonary thromboemboli.

Strictly speaking, septic embolisation is not coagulopathy-related and although there is strong evidence for ischaemic events resulting from septic emboli, we will not include this in the model of COVID-19-related coagulopathy.

###### 2. Secondary infection

(Meza et al, 2020) report 4 cases of diabetic ketoacidosis in COVID-19 without respiratory symptoms but with bacterial coinfection in 2 cases. (Zhou et al, 2020) present a case of DVT with acute limb ischaemia and discuss the possible cause of DVT in COVID-19 and include secondary bacterial and fungal infections as risk factors. Many of the risk factors overlap with those of (McLendon et al, 2020) mentioned previously including infection. DVT’s in turn can predispose to pulmonary emboli.

(Manna et al, 2020) provide an overview of viral infections with secondary bacterial infection. They cite evidence of an elevated risk of secondary bacterial infection with viral infection and also outline some of the mechanisms by which this may occur.

Given the size of the pandemic it is unsurprising that there is a significant evidence base for coinfection with COVID-19. (Verroken et al, 2020) found bacterial co-infection in 40.6% of patients admitted to ICU with COVID-19 with the main bacteria being Staphylococcus Aureus, Haemophilus Influenza and Moraxella Catarrhalis. (Kim et al, 2020) found that 20.7% of 116 specimens from patients with SARS-COV2 were positive for co-infection. Rhinovirus, enterovirus, respiratory syncytial virus and non-SARS-CoV-2 Coronoviridae were the most common co-infections. Interestingly they found that co-infections were more likely in samples without SARS-CoV-2 infection but this relationship was not significant for any of the individual organisms using a Chi squared test (P < 0.05).

(Lansbury et al, 2020) completed a systematic review and meta-analysis of coinfections in people with COVID-19 (n=3834). They found that 7% of patients with COVID-19 in hospital had bacterial infection and that this figure was higher in ICU compared to the wards and the most common bacteria were Mycoplasma Pneumonia, Pseudomonas Aeruginosa and Haemophilus Influenza while Respiratory Syncytial Virus and Influenza A were the most common viruses. Fungal coinfections were also identified. (Arastehfar et al, 2020) report on 35 published cases of COVID-19 associated pulmonary aspergillosis (CAPA). (Cucchiari et al, 2020) report on a case series of 5 patients with COVID-19 and pneumococcal pneumonia superinfection identified with urinary antigen. (Peddu et al, 2020) demonstrated viral and bacterial coinfection with SARS-COV2 using metagenomic analysis. (He et al, 2020) found that in severe COVID-19 with reduced T-cell count there was a higher risk of bacterial and fungal infection.

There are numerous case reports identifying coinfection and providing evidence of unusual coinfections, comorbidities or other insights. (Shah et al, 2020) report a case of Covid-19 with coccidioidomycosis. (Langerbeins et al, 2020) report a case of chronic lymphocytic leukemia with SARS-COV2 infection with parainfluenza coinfection. (Jose and Desai, 2020) report a case of COVID-19 with rapid deterioration following an E.Coli superinfection. (Menon et al, 2020) report a case of SARS-CoV-2 comorbid with Pneumocystitis Jirovecci and note that this is commonly seen when there is are deficiencies in T-cell immunity. (Lehmann et al, 2020) found a rate of coinfection of 3.7% of patients but this increased to 41% in those admitted to ICU. (Blaize et al, 2020) report a case of COVID-19 with invasive pulmonary aspergillosis. (Poignon et al, 2020) report a case of COVID-19 with invasive pulmonary fusariosis. (Castiglioni et al, 2020) report a case of superinfected pneumatocoles in a case of COVID-19. The pneumatoceles were suspected to have resulted from the SARSCOV2 infection. (Muller et al, 2020) report a case of Covid-19 comorbid with hepatitis C and HIV. (Konala et al, 2020) present 3 cases of coinfection with COVID-19 and influenza.

In summary, secondary infection can lead to DVT’s and there is an established evidence base for this independent of COVID-19. There is also an abundant evidence base for secondary infection with COVID-19 although this does not focus specifically on coinfection-mediated coagulopathy. Interestingly coinfection was actually less than in the control group in (Kim et al, 2020) although this does not negate the significance of coinfection in COVID-19. The other complicating factor is that identifying the organism still requires a clinical correlate at a time where COVID-19 may dominate the clinical presentation. (Castioglioni et al, 2020) report on a case of superinfected pneumatoceles provides direct evidence of a potential structural mediator of respiratory superinfection.

###### I. Stroke risk with infection

(Morassi et al, 2020) discuss the importance of the relationship between stroke and infection. They cite a paper examining this relationship in more detail (Emsley and Hopkins, 2008). (Emsley and Hopkins, 2008) suggest that up to one third of strokes are preceded by infections.

(Miller and Elkind, 2016) provide an overview of the relationship between infection and stroke. They identify 22 infectious organisms where evidence has been found of an association with stroke and which include viruses, bacteria and fungi. The pathological mechanisms include arachnoiditis, vasculitis, compression of large arteries by cysts, arterial occlusion by infected erythrocytes, cardioembolism, microembolic infarction, larval obstruction of small vessels, meningitis, arteritis, vascular necrosis, aneurysmal dilatation, opportunistic CNS infections, accelerated atherogenesis, enhanced platelet aggregation, prothrombotic state and chronic inflammation.

(Elkind et al, 2010) identified an increased risk of ischaemic stroke with increasing infection burden involving five common infections in the Northern Manhatten Study. (Benjamin et al, 2012) review the evidence for stroke in HIV infection noting that 1-5% of patients with HIV experience stroke and cerebral ischaemic lesions are found in up to 34% of autopsies. They suggest a number of possible mediators for stroke risk in HIV: HIV associated vasculopathy, vasculitis, accelerated atherosclerosis, opportunistic infections, neoplasia, bacterial endocarditis, marantic endocarditis, HIV-associated cardiac dysfunction, HIV-associated hyperviscosity and ischaemic heart disease. In a systematic review and meta-analysis of studies looking at Hepatitis C infection and stroke risk, (Huang et al, 2013) found an odds ratio of stroke with HCV infection of 1.97 (95% CI 1.64-2.30).

(Cowan et al, 2016) published their findings in the Atherosclerosis Risk in Communities (ARIC) cohort. The odds ratio for stroke following a hospitalised infection in the previous 14 day-period was 7.7 (95% CI: 2.1-27.3). (Erskine et al, 2017) completed a meta-analysis which included 7.9 million patients who had been infected with herpes zoster. They found increased odds of up to 40% of experiencing a cerebrovascular event at between 3 months and 1 year after the infection onset.

In summary there is a strong evidence base for an increased risk of stroke with infection with multiple mechanisms identified including coagulopathy. In terms of modelling this can be grouped with secondary infections in COVID-19.

##### I. Autopsy/Biopsy Finding Related Mechanisms

###### 1. Pyroptosis

Pyroptosis was described as a potential mechanism by (Varga et al, 2020). Pryoptosis is a form of programmed cell death which involves inflammation. (Li et al, 2020) noted that the pathways mediating pyroptosis are poorly understood and they investigated the signalling pathways involved in Pneumonia associated sepsis. They found an important role for IL-17 and outline details of the signalling pathway. (Yang et al, 2020) in their comparison of SARS and SARS-COV2 propose a role for pyroptosis in the pathogenesis of SARS-COV2. We have included this under the broader heading of sepsis.

###### 2. Apoptosis

Apoptosis was described in (Varga et al, 2020). Apoptosis is programmed cell death and is associated with characteristic histopathological findings. There is a suggestion that apoptotic pathways may be important in cases of severe sepsis. (Luan et al, 2015) characterised apoptotic pathways in immune cells in severe sepsis in their study. As with pyroptosis, we have included apoptosis with the broader topic of sepsis whilst also noting that there is a strong evidence base for apoptosis in COVID-19.

##### Additional Mechanisms

###### A. Alternative Complement Pathway

More recently evidence has emerged for an important role of the alternative complement pathway. The complement pathway is part of the innate immune response and is divided into three pathways – the classical, alternative and lectin complement pathways. (Brodszki et al, 2020) provide a guideline for the complement deficiencies and their management and review the evidence on complement deficiencies and the regulation of complement. They note a thrombotic microangiopathy associated with dysregulation of the alternative complement pathway. Furthermore there are a number of complement deficiencies that are associated with autoimmune disorders such as SLE.

(Chatzidionysiou et al, 2020) consider the question of whether COVID-19 is a complementopathy. (Rambaldi et al, 2020) report initial findings in an investigation of a lectin pathway inhibitor in patients with COVID-19-related ARDS and suggest the results support a role for the lectin pathway in COVID-19-related pathophysiology. (Magro et al, 2020) reported on a case series of 2 post-mortems and biopsies from 3 patients with skin involvement all with COVID-19. They found evidence from the lungs and skin of marked involvement of the lectin and alternative complement pathways. They suggest that a COVID-19-related generalised thrombotic microvascular injury results from complement activation. Numerous cases of collapsing glomerulopathy have been described in COVID-19 (Larsen et al, 2020)(Gaillard et al, 2020)(Peleg et al, 2020)(Nasr et al, 2020)(Magoon et al, 2020)(Jhaveri et al, 2020)(Kadosh et al, 2020). Collapsing glomerulopathy as a morphological variant of focal segmental glomerulosclerosis is in turn associated both with thrombotic microangiopathy (Buob et al, 2016) and alternative complement pathway activation (Thurman et al, 2015). C3 glomerulopathy is another condition involving the kidneys and resulting from a dysregulated alternative complement pathway activation (Caravaca-Fontán et al, 2020).

In summary, multiple lines of evidence, some indirect, suggest a role for the alternative complement pathway in COVID-19-related coagulopathy. The role of complement deficiencies in COVID-19 should also be borne in mind.

###### B. General Factors Relating to Arterial Thrombosis

(Lowe and Tait, 2009) identify a number of risk factors relating to arterial thrombosis and suggest that it is the role of all healthcare practitioners to reduce these events through pharmacological prevention and education. The risk factors they identify are traditional risk factors for cardiovascular disease and include dyslipidemia, psychosocial factors, diabetes, hypertension, smoking and abdominal obesity as well as protective factors including fruit and vegetables and exercise. Whilst smoking and obesity are also risk factors for DVT as per Table 25, dyslipidemia, psychosocial factors, diabetes and hypertension are not listed in Table 25 and suggest a more chronic aetiology and a role for endothelial dysfunction. This however is contrast with the acute nature of arterial thromboses in COVID-19.

###### C. Medical Device-Related Coagulopathy

In the 56 main papers, there were multiple examples of medical device-related coagulopathy (including catheters). We suggest a number of mechanisms are involved. Firstly catheters may result in subtle vascular wall damage that ordinarily would not present any problems but when combined with the vasculopathy of COVID-19 acts synergistically to increase the risk of thrombosis. Secondly in the case of the ECMO centrifuge, (Helms et al, 2020) suggest that this is due to a combination of ultrafiltration and high fibrinogen levels. (Kowalewski et al, 2020) compare coagulation and inflammation in COVID-19 and ECMO and note the pro- and anticoagulant properties of ECMO. Thirdly when the blood is in contact with artificial surfaces this can trigger the intrinsic pathway which amplifies the extrinsic pathway.

###### D. Vulnerabilities

There are a number of factors that act as modifiers of risk of mortality in COVID-19. (Williamson et al, 2020) looked at 10,926 COVID-19-related deaths in 17,278,392 adults using pseudonymised records from primary care. They identified a number of risk factors for death in COVID-19 (see Table 34).

**Table 34.** Risk Factors from (Williamson et al, 2020)

|  |
| --- |
| Diabetes |
| Haematological malignancy |
| Non-haematological malignancy |
| Age |
| Male |
| Black, South Asian & Mixed and Other ethnicity |
| Obesity |
| eGFR <= 60 ml/min per 1.73 m<sup>2</sup> |
| Chronic respiratory disease |
| Chronic liver disease |
| Stroke & Dementia |
| Other neurological disease |
| Conditions associated with immunosuppression |
| Rheumatoid arthritis, lupus or psoriasis |
| Hypertension up to age 70 |

(Marazeula et al, 2020) examined the endocrine and metabolic aspects of COVID-19. They suggest four ways in which obesity may worsen outcome in COVID-19 - reducing respiratory reserve, comorbidities, increased viral shedding and chronic inflammation. However a key factor for consideration with vulnerabilities is endothelial dysfunction and this relates to coagulopathy.

(Amraei and Rahimi, 2020) suggest that endothelial dysfunction is a common factor in a number of comorbidities that impact on COVID-19 severity including obesity, diabetes and cardiovascular disease including hypertension. They suggest that endothelial cell injury by SARS-COV2 both in the lungs and elsewhere may be an important consequence of infection and would explain the elevated Von Willebrand Factor levels. (Bansal et al, 2020) review the evidence for worse outcomes in COVID-19 with hypertension, diabetes and obesity. They further highlight the presence of endothelial dysfunction in diabetes type 2 and obesity. One study found an odds ratio of 1.67 for severe COVID-19 with obesity compared to non-obesity (95%CI: 1.43, 1.96; P<0.001)(Huang et al, 2020).

(Hayden, 2020) reviews endothelial activation and dysfunction in diabetes Type 2, metabolic syndrome and COVID-19. Type 2 diabetes is noted to affect a number of tissues including the lungs through a range of mechanisms including endothelial activation and dysfunction. Furthermore the blood gas barrier (BGB) is formed by the fusion of the basement of the type I pneumocytes and alveolar endothelial cells and plays a role both in oxygen exchange and after endothelial activation also plays a role in the immune response to SARSCOV2. The BGB endothelium is activated by cytokine storms, binding of SARS-COV2 to the ACE-2 receptors on the pulmonary endothelium and via a SARS-COV2 induced redox storm mediated via reactive oxygen and nitrogen species (RONS). A key finding is the loss of the pulmonary epithelial glycocalyx in both diabetes type 2 and the metabolic syndrome and it is suggested that this could account for the vulnerability of people with type 2 diabetes to more severe COVID-19.

Of note in (Williamson et al, 2020) splenectomy was associated with increased risk of mortality in COVID-19 although this did not reach statistical significance. Nevertheless postsplenectomy there is noted to be an increased risk of arterial thrombosis (Long et al, 2020). (Zhou et al, 2020) in their review cite evidence of destruction of secondary lymphoid tissue in autopsy findings including splenic atrophy and focal haemmorhagic necrosis. The possibility of a susceptibility to COVID19 related coagulopathy in this population would require further evidence.

(Falasca et al, 2020) report on a number of post-mortem findings including splenic congested red pulp and lymphoid hypoplasia whilst many of the remaining findings relate to apparent non-coagulopathy-related pathologies in COVID-19. (Qasim et al, 2020) report a case of splenic artery thrombosis and infarct in COVID-19. (Shaukat et al, 2020) report a case of atraumatic splenic rupture in COVID-19.

In terms of genetic susceptibility (Zeberg & Pääbo, 2020) identified a haplotype that presents increased risk for severe COVID-19.

In summary there are a number of vulnerabilities that include conditions that lead to endothelial dysfunction, genetic susceptibility, age, ethnicity, male gender. However these are more general susceptibilities not specifically related to COVID-19-related coagulopathy although they may be very relevant particularly endothelial dysfunction. We will include them cautiously in the model on the proviso that their relationship with the COVID-19-related coagulopathy requires further clarification.

###### E. Type-III Hypersensitivity Reaction

In the type III hypersensitivity reaction, antibody complexes are deposited in various tissues in the body. The complement pathway can be involved in the pathogenesis of the type III hypersensitivity reaction and so this can be considered together with the section on the alternative complement pathway. (Granger et al, 2019) review the evidence on the relationship between NET’s and autoimmune diseases. They note that with deficiency in DNAase 1 activity which results in elevated NET’s there is an association with active lupus and provide additional evidence of the relationship between NEs and lupus. They also note that difference in the components of NET’s in patients with and without SLE.

There is evidence that when senescent neutrophils are not cleared through apoptosis this is associated with the development of autoimmune disease (Kawano et al, 2018). (Tanaka et al, 2020) have demonstrated an association between apoptosis-associated release of autoantigens and SLE. (Yang et al, 2019) note that autoantigens and damage associated molecular patterns (DAMPS) are released after apoptosis, pyroptosis, necroptosis and by NET’s resulting in inflammation and with a potential for autoimmune reactions.

(Roncati et al, 2020) describe a case of ischaemic bowel and spleen. They describe the histopathological findings and demonstrate evidence of fibrinoid necrosis. They suggest that naїve helper T-cells switch the immune system to a type-2 helper T-cell response rather than a type-1 helper T-cell response which then escalates to a type-III hypersensitivity reaction with immune-complexes deposited in the blood vessel walls resulting in a systemic vasculitis.

(Mahdi, 2020) proposes a model of COVID-19-related type-III hypersensitivity. (Favelli et al, 2020) draw parallels between rheumatoid arthritis and COVID-19 and ask whether the latter can be consider an autoimmune condition. (Deshmukh et al, 2020) report a case of collapsing glomerulopathy with COVID-19 in which there were no pre-existing risk factors and they attributed the pathology to COVID-19. (Cossey et al, 2017) note that there are many causes of collapsing glomerulopathy. (Gupta et al, 2020) report two cases of collapsing glomerulopathy with podocytopathies and (Sharma et al, 2020) presents two cases of COVID-19associated collapsing focal segmental glomerulosclerosis. Further cases of collapsing glomerulopathy are discussed in the section on the alternative complement pathway.

In summary there is evidence of a type-III hypersensitivity reaction in COVID-19 with deposition of immune complexes in the skin, kidneys and vascular walls in association with vasculitis which in turn can lead to a coagulopathy. We think this is likely to occur in a minor proportion of cases of coagulopathy although further data would be needed to draw even initial conclusions on this.

## Towards a Model of COVID-19-Related Coagulopathy

In our paper, we draw together the findings from many studies to develop a testable model as per Table 35 and the diagrammatic mapping (see Figures 10-21). This is a simple model with several components and is described by causal relationships but without quantitative descriptions of those same relationships. The purpose of this model is to serve as a starting point for further enquiry and to enable other researchers to refute or confirm these relationships or to quantify and expand upon them, thereby improving the understanding of the COVID-19-related coagulopathy.

**Table 35.** A model of coagulopathy in COVID-19 colour coded according to Virchow’s triad: Purple – three elements of Virchow’s triad combined, yellow – hypercoagulability, blue – stasis, green – endothelial damage, black – not applicable, white – no category of Virchow’s criteria, red – combination of endothelial damage and hypercoagulability, General evidence – evidence for relationship with coagulopathy independently of COVID-19, Evidence in COVID-19 – evidence of relationship with coagulopathy in COVID-19.

| Revised Taxonomy for COVID-19-related Coagulopathy Mechanisms | General Evidence | Evidence in COVID-19 |
| --- | --- | --- |
| A. Viral Life Cycle |  |  |
| A1. Attachment |  |  |
| A2. Penetration |  |  |
| A3. Uncoating |  |  |
| A4. Gene Expression and Gene Replication |  |  |
| A5. Assembly |  |  |
| A6. Release |  |  |
| B. Host Immune Response |  |  |
| B1. Non-Pathological Response |  |  |
| B2. Cytokines |  |  |
| B2a. IL-6 |  |  |
| B2a1. IL-6-associated coagulopathy | Moderate | Weak |
| B2b. Cytokine storm | Strong | Moderate |
| B3. Innate immunity-mediated mechanisms |  |  |
| B3a. Alternative complement pathway activation | Moderate | Moderate |
| B3b. Neutrophil extracellular traps | Strong | Strong |
| B4. Adaptive immunity-mediated mechanisms |  |  |
| B5. Antiphospholipid Antibody Syndrome-Like Coagulopathy | Strong | Strong |
| B6. Immunothrombosis | Strong | Strong |
| B7. Type-III hypersensitivity | Strong | Moderate |
| B8. Sepsis-induced coagulopathy | Strong | Moderate |
| B9. Sepsis-associated DIC | Strong | Weak |
| C. Hypercoagulable state |  |  |
| C1. Hyponatraemia | Strong | Strong |
| C2. Hypokalaemia | Strong | Strong |
| C3. Calcium | Strong | Strong |
| C4. Dehydration | Strong | Nil |
| C5. Immune-mediated mechanisms (B2-B10) |  |  |
| C6. Angiotensin II | Strong | Moderate |
| C7. Clotting cascade | Strong | Strong |
| C8. Hypoxaemia | Strong | Strong |
| C9. Hyperviscosity | Strong | Moderate |
| D. System Involvement |  |  |
| D1. Cardiovascular |  |  |
| D1a. Vasculature |  |  |
| D1a1. Microangiopathy with thrombosis | Strong | Strong |
| D1a2. Endotheliitis | Strong | Strong |
| D1a3. Vasculitis | Strong | Strong |
| D1b. Myocardium |  |  |
| D1b1. Direct invasion of the myocardium |  |  |

**Table 35 (cont'd)**
| **Revised Taxonomy for COVID-19-related Coagulopathy Mechanisms** | **General Evidence** | **Evidence in COVID-19** |
| --- | --- | --- |
| D1b2. Cardioembolism secondary to cardiac injury | Strong | Weak |
| D1b3. Secondary to cardiogenic shock | Strong | Moderate |
| D1b4. SARS-COV2 myocarditis |  | Weak |
| D1b5. Stress cardiomyopathy | Strong | Strong |
| D1b6. Atrial fibrillation | Strong | Strong |
| D2. Respiratory |  |  |
| D2a. ARDS |  |  |
| D2a1. Respiratory failure with hypoxia leading to myocardial injury | Strong | Weak |
| D2a2. DIC | Strong | Strong |
| D2a3. Mechanical ventilation-mediated coagulopathy |  |  |
| D2a4. ARDS-related coagulopathy | Strong | Strong |
| D3. Central Nervous System |  |  |
| D3a. Direct nervous system injury leading to coagulopathy |  | Weak |
| **E. Factors Relating to mild COVID-19** |  |  |
| E1. Immobilisation/Bed Rest | Strong | Nil |
| E2. Dehydration due to fever, diarrhoea | Strong | Nil |
| **F. Factors Relating to severe COVID-19** |  |  |
| F1. Critical-illness encephalopathy | Weak | Weak |
| F2. Stress cardiomyopathy |  |  |
| F3. Atrial Fibrillation |  |  |
| F4. Metabolic/electrolyte disturbance | Strong | Strong |
| G. Receptor-mediated |  |  |
| G1. ACE2-receptor-mediated pathways | Strong | Moderate |
| H. Iatrogenic |  |  |
| H1. Pharmacological adverse events | Strong | Nil |
| H2. Medical Device-related clotting events | Strong | Strong |
| I. Vulnerabilities | Strong | Strong |

**Figure 10.**
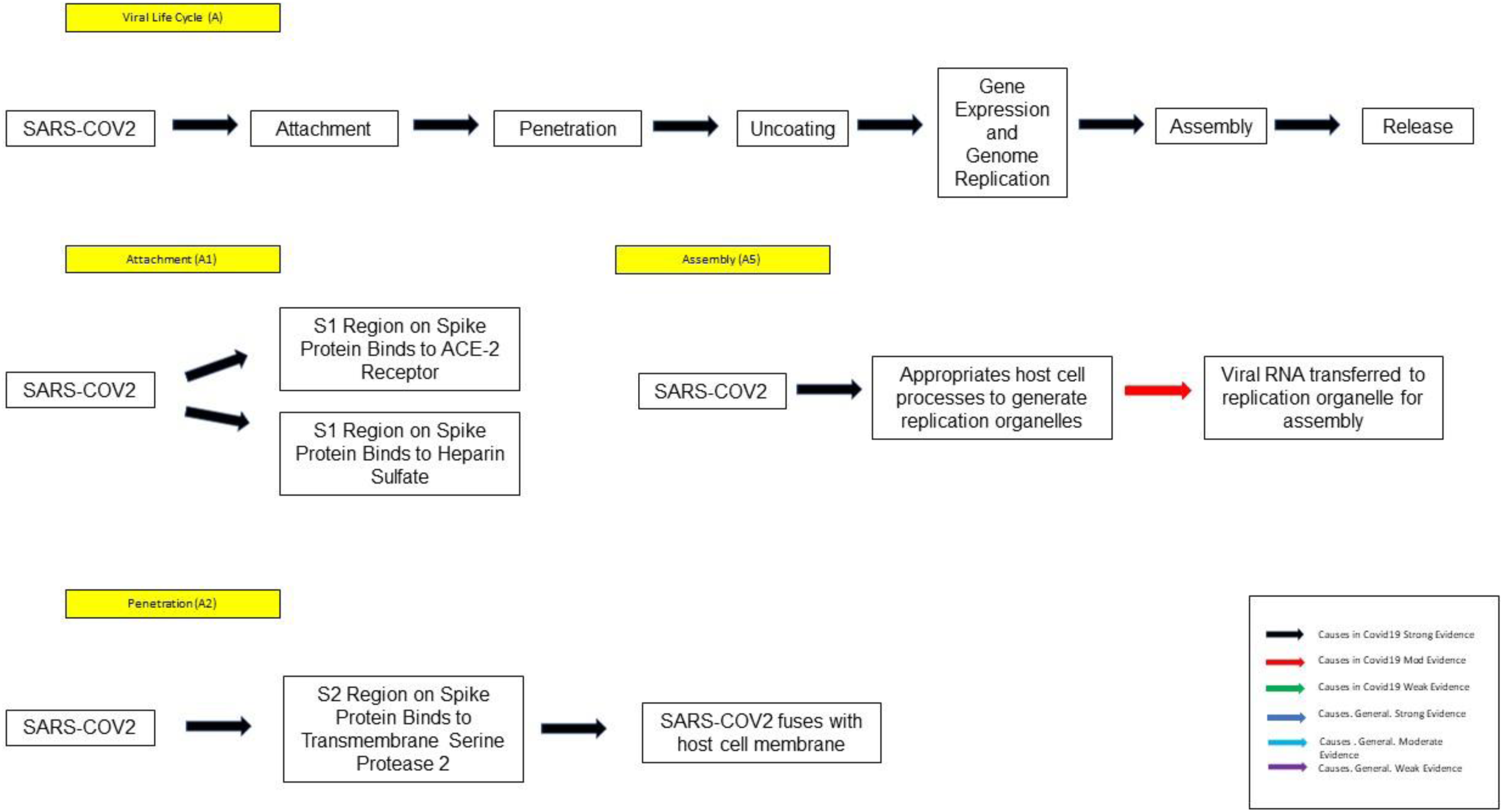
Viral Life Cycle.

**Figure 11.**
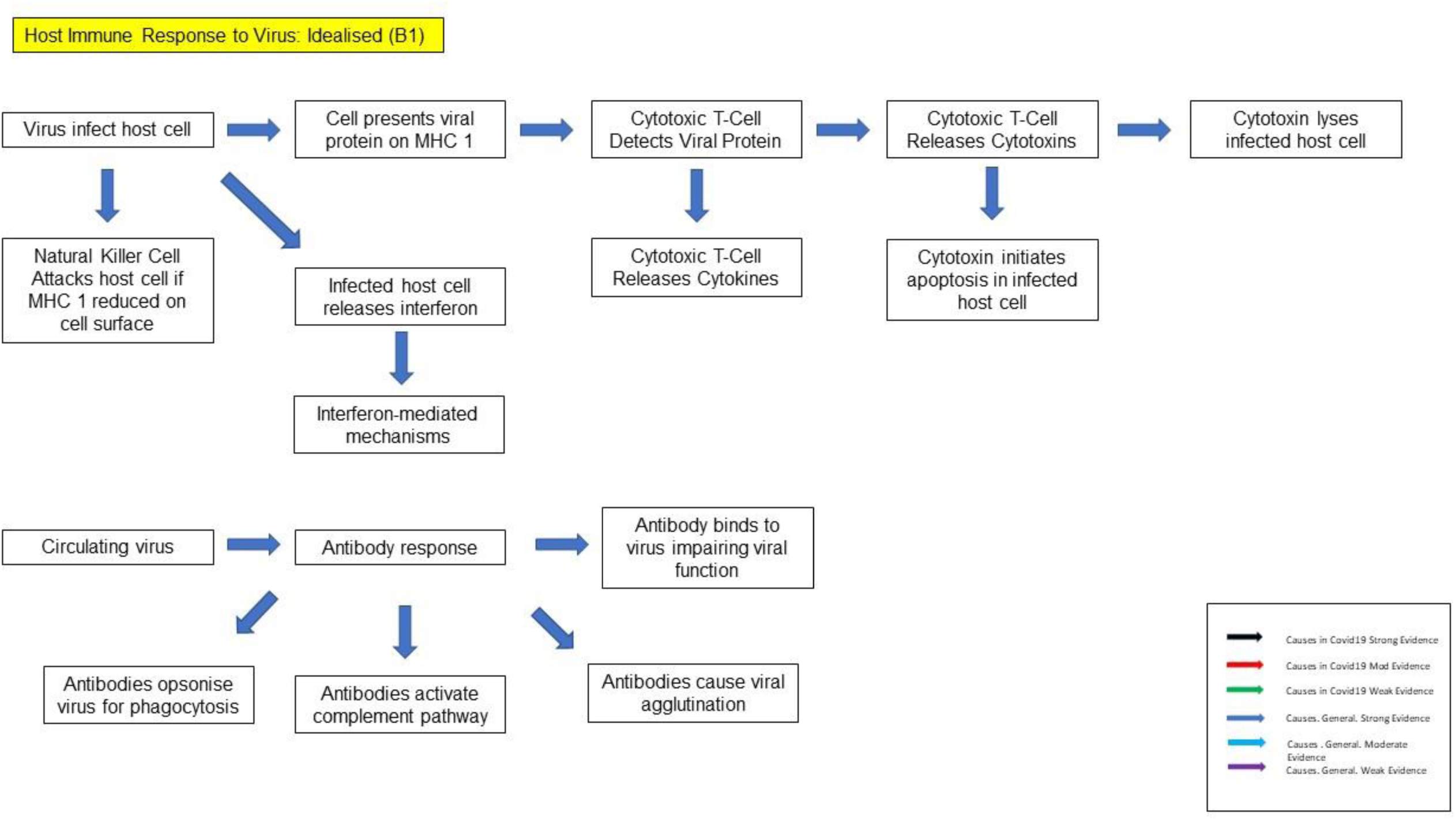
Idealised Viral Host Immune Response (Non-Pathological)

**Figure 12.**
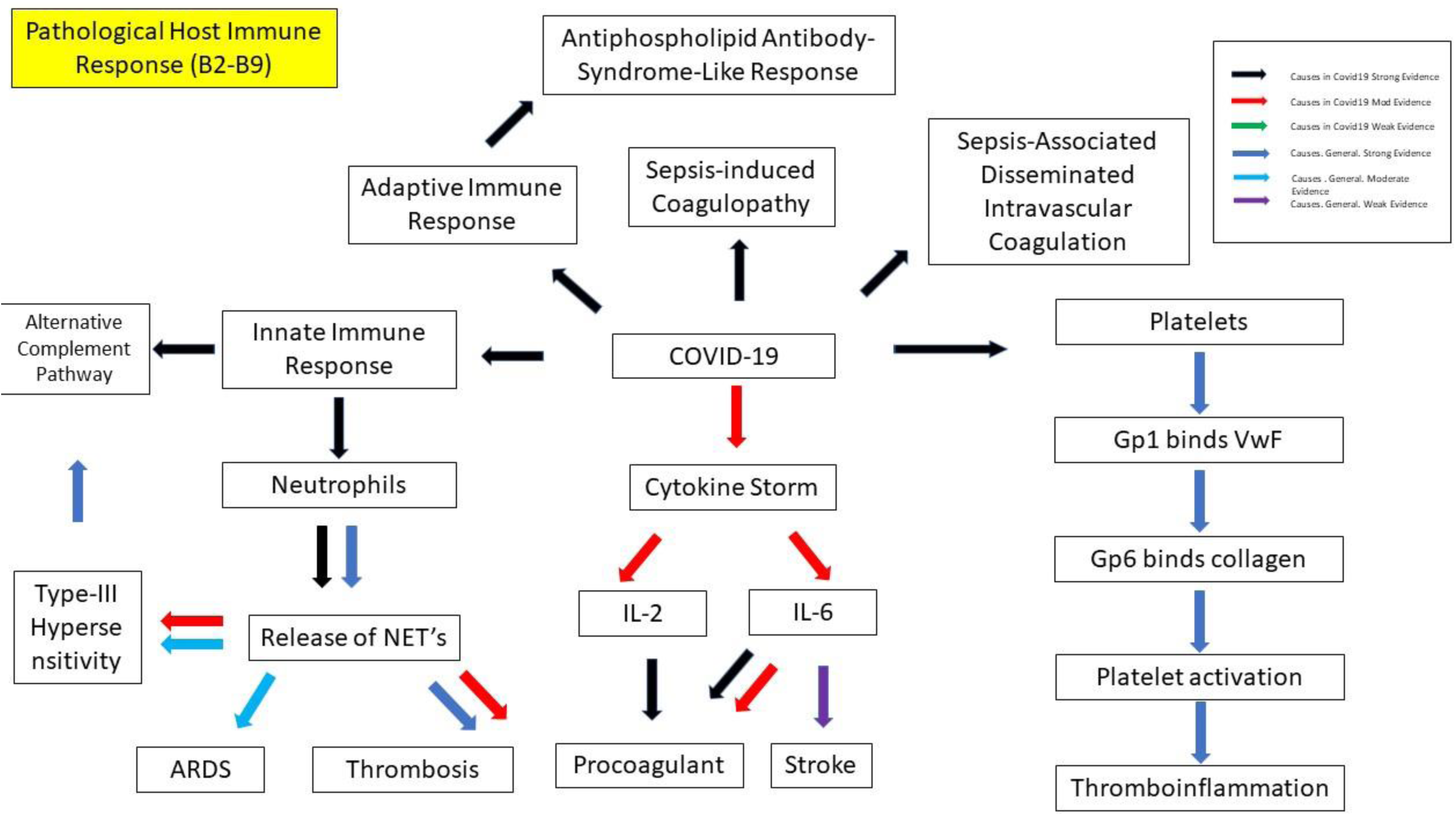
Pathological Host Immune Response in COVID-19.

**Figure 13.**
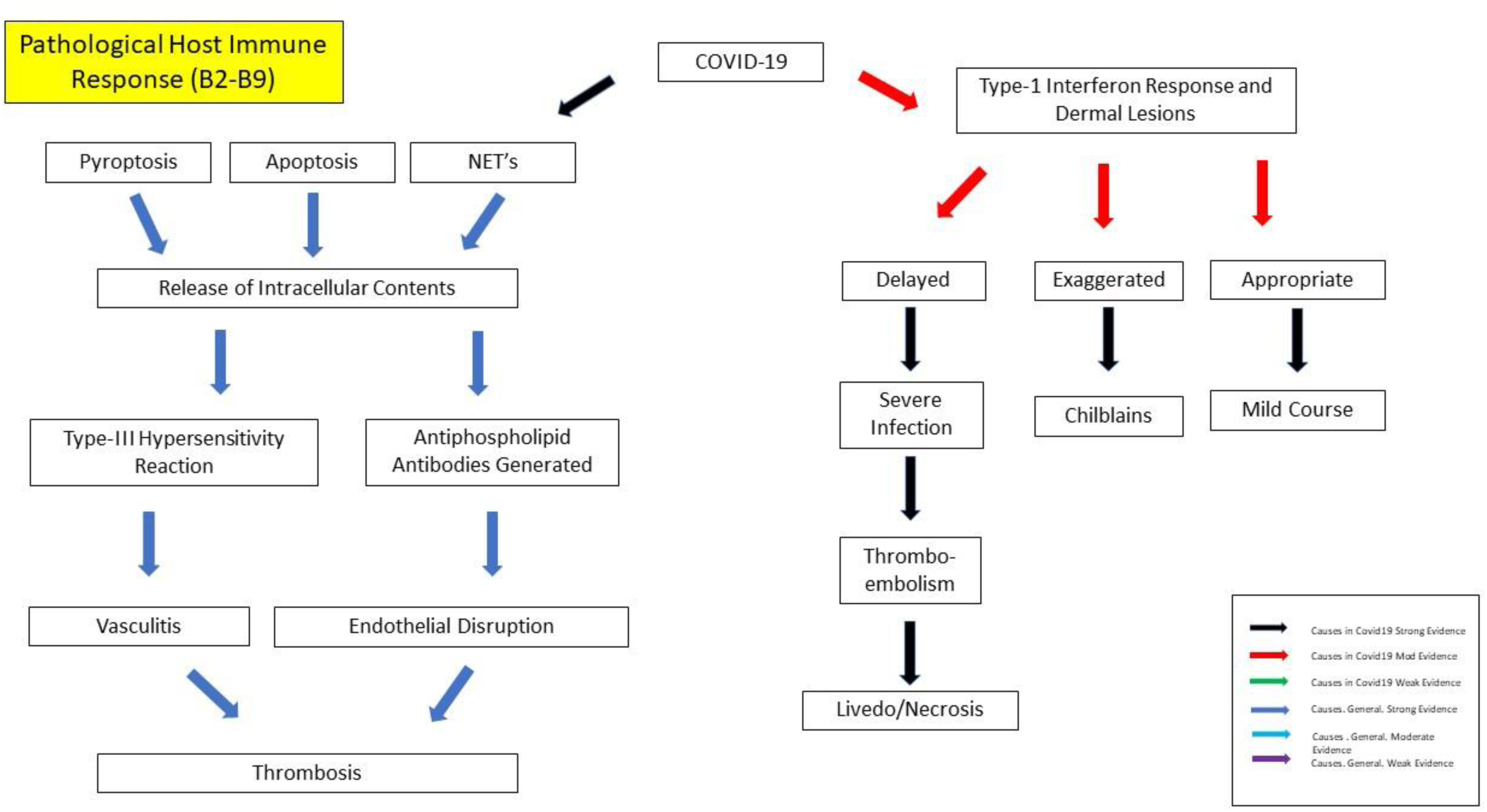
Pathological Host Immune Response in COVID-19: 2^nd^ Diagram.

**Figure 14.**
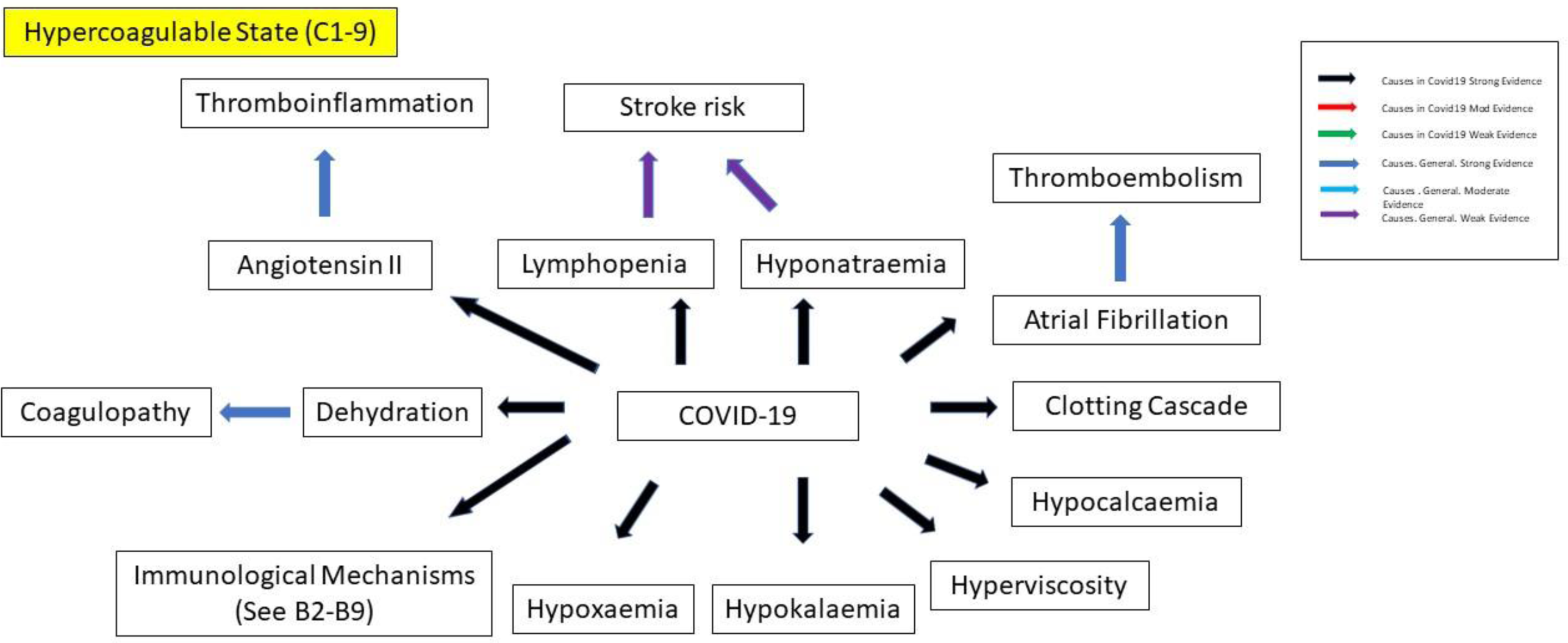
Hypercoagulable State in COVID-19.

**Figure 15.**
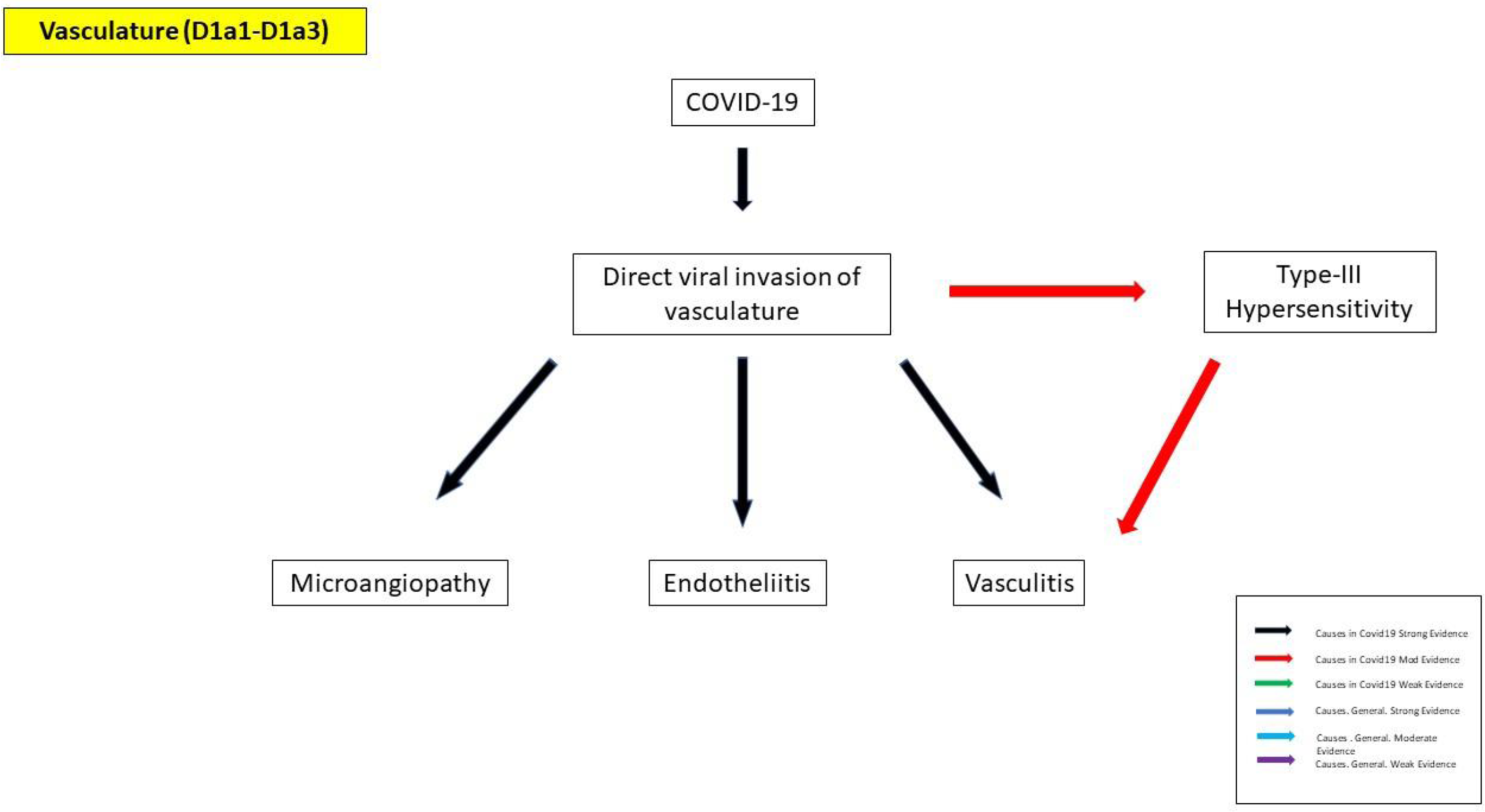
Involvement of the Vasculature in COVID-19.

**Figure 16.**
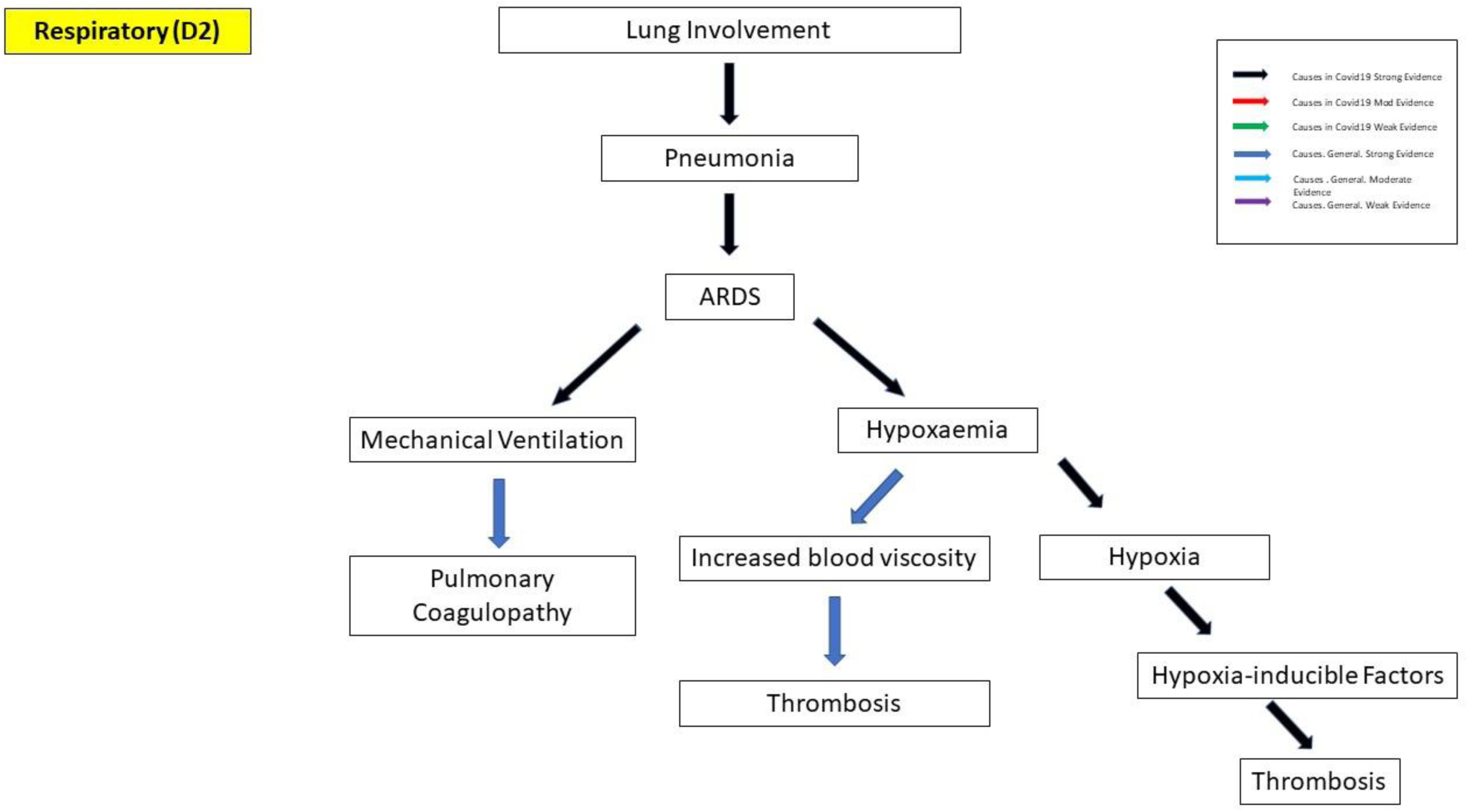
Respiratory Involvement in COVID-19.

**Figure 17.**
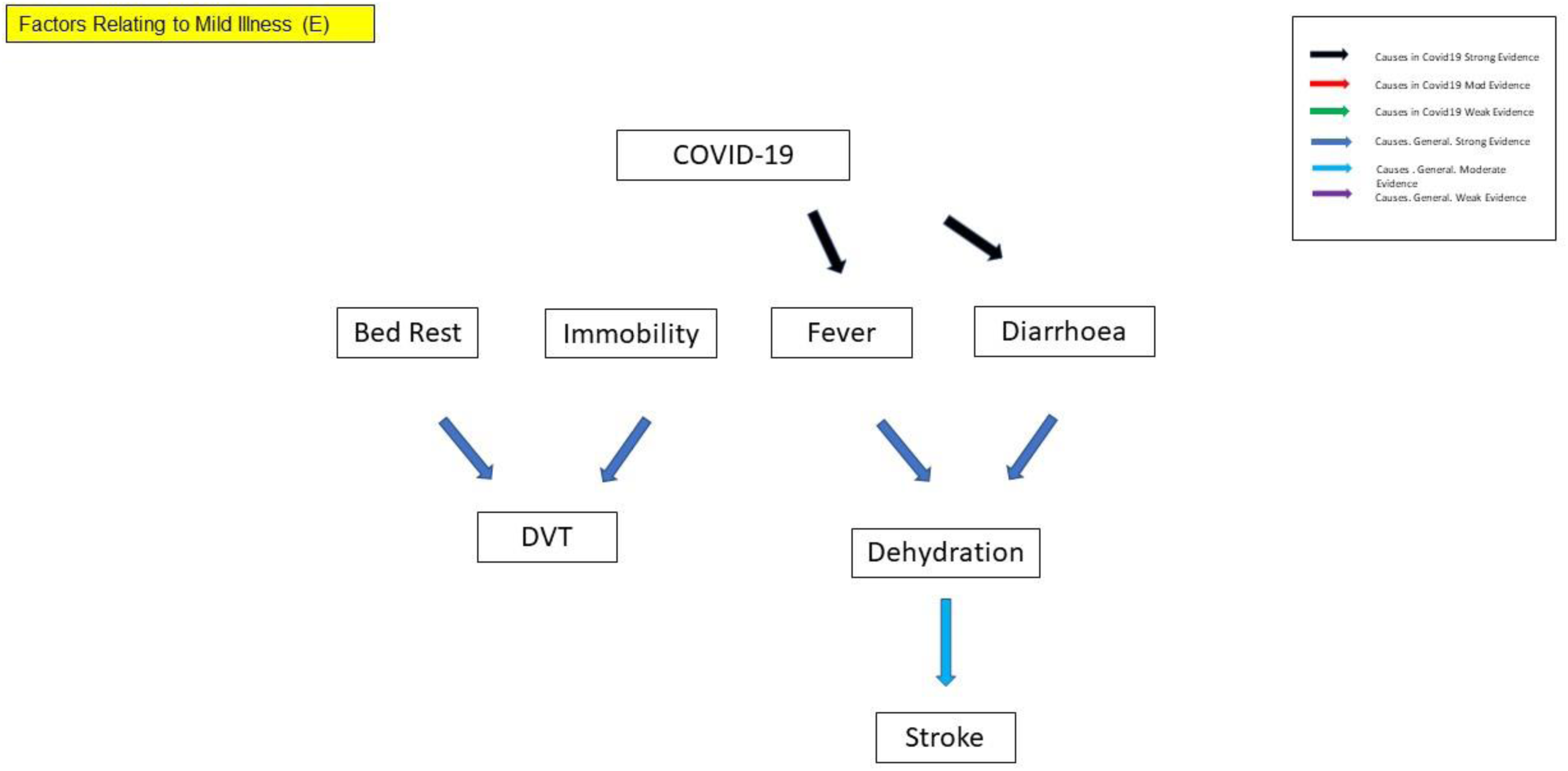
Factors Relating to Mild Illness in COVID-19.

**Figure 18.**
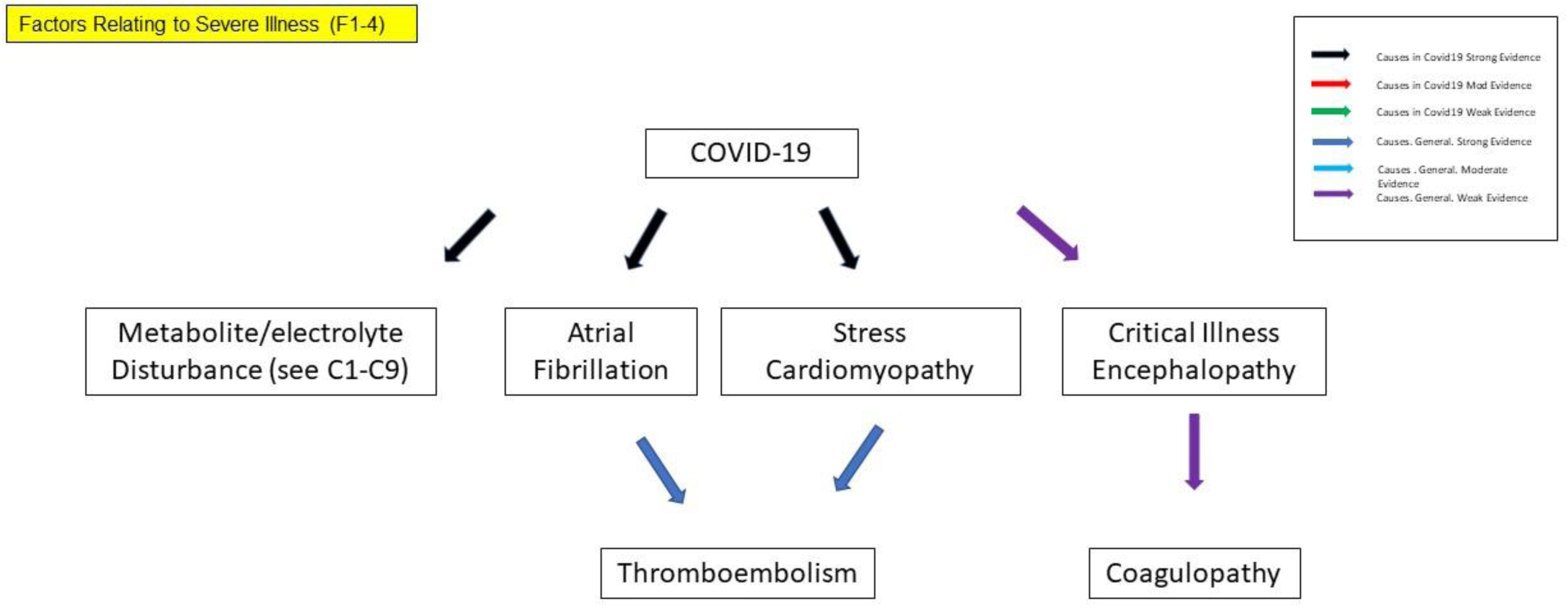
Factors Relating to Severe Illness in COVID-19.

**Figure 19.**
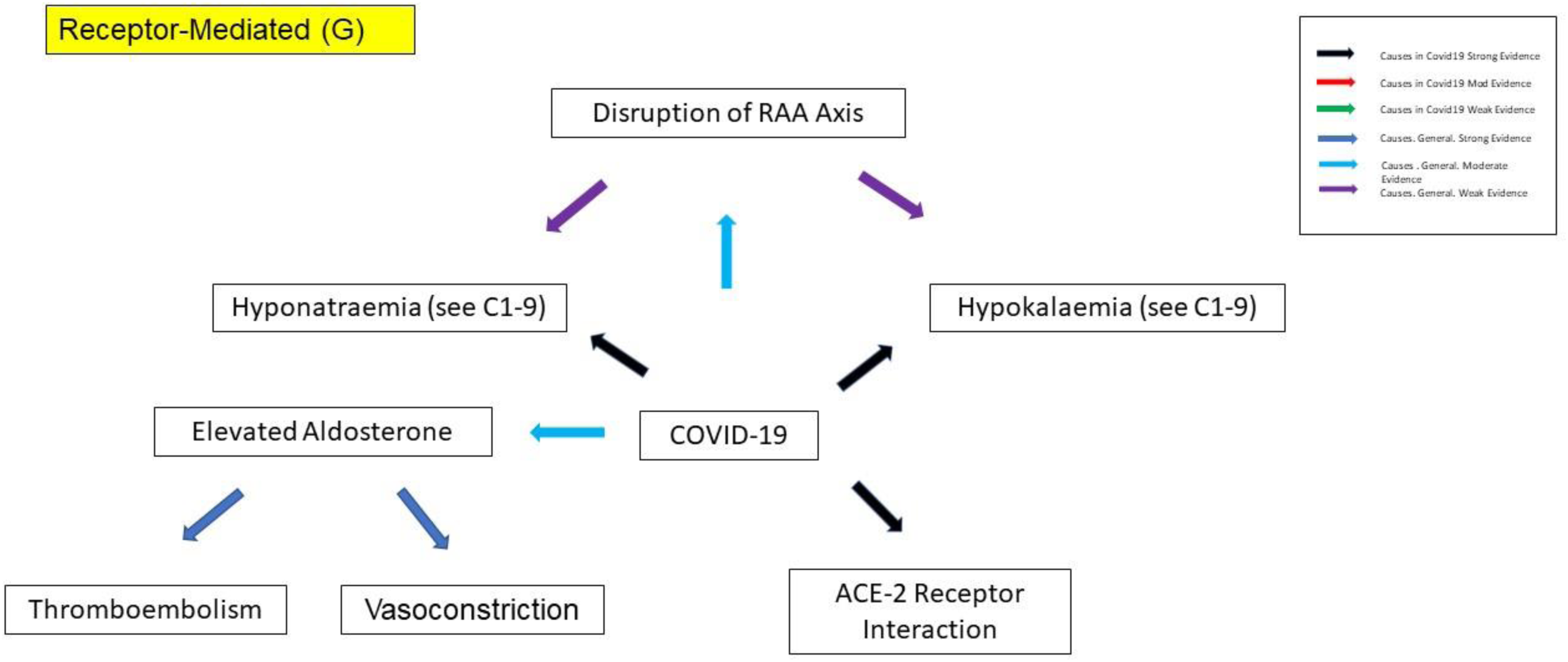
Receptor-mediated Mechanisms in COVID-19.

**Figure 20.**
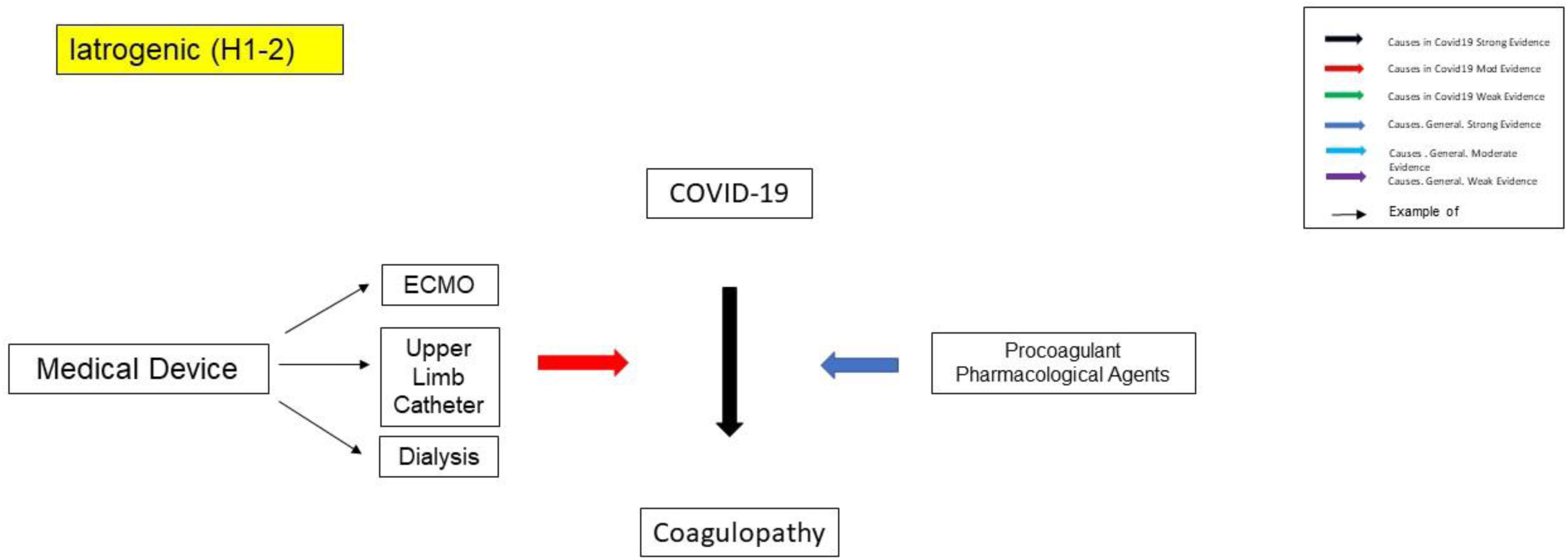
Iatrogenic Mechanisms in COVID-19.

**Figure 21.**
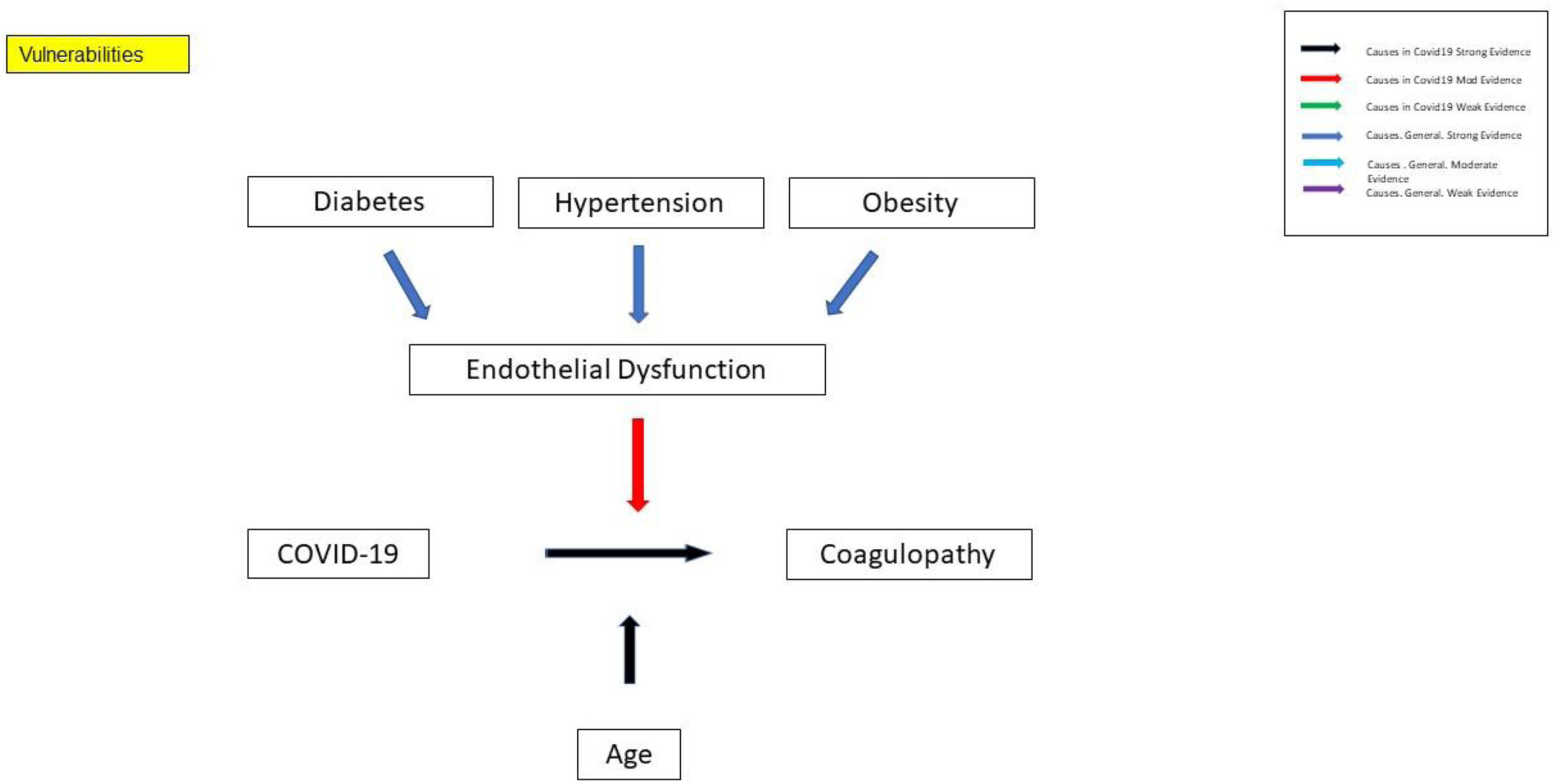
Vulnerability Factors in COVID-19.

### Revised Aetiologies for COVID-19

The revised aetiologies are shown in Table 35. The revised aetiologies have been organised into categories following the preceding analysis. The general evidence is then considered for each potential aetiology and categorised as weak, moderate or strong. The general evidence refers to the evidence for this aetiology leading to a coagulopathy independently of COVID-19 and this is contrasted with the evidence for this aetiology leading to a coagulopathy in cases of COVID-19. The revised aetiologies then form the basis for the development of the model together with the corresponding narrative descriptions

### A. Viral Life Cycle

Since our enquiry has focused on the coagulopathy, the model can be described as per the introduction section on the viral life cycle. From the literature we have reviewed, it is unclear which aspects of the life cycle lead to the various coagulopathy-related pathology and the details thereof. The mechanisms to be accounted for would be the steps leading to the immune response as well as the mechanisms of damage to the tissues themselves. We may infer empirically that there is a causal sequence of viral invasion of tissues and subsequent tissue damage and immune response and this is reflected in the diagrammatic mapping.

### B. Host Immune Response

#### B1. Non-pathological host immune response

Again since our enquiry focused on the coagulopathy, the model can be described as per the introduction section on the host response and this is reflected in the diagrammatic mapping. We have characterised the pathological response as one which may have led to the coagulopathy.

#### B2-9. Pathological host immune response

We have divided up the pathological host immune response into the general mechanisms common to both innate and adaptive immune response (cytokines), innate immune response (NETs and complement pathway), adaptive immune response (antiphophospholipid antibody syndrome-like coagulopathy), immunothrombosis, type III hypersensitivity and recognised coagulopathies – sepsis-induced coagulopathy and DIC although this can occur independently of the immune-response we have included it with sepsis in COVID-19.

##### B2a. IL-6

###### B2a1. IL-6-associated coagulopathy

In the non-COVID-19 literature, there is evidence of an association between elevated IL-6 levels and stroke as well as a suggested putative mechanism to mediate a coagulopathy. In COVID-19 there is controversy regarding elevation of IL-6 levels.

##### B2b. Cytokine storm

There is evidence of procoagulant effects of several cytokines in the non-COVID-19 literature. In COVID-19 there is a suggestion that the cytokine storm could lead to ARDS and there is an association between cytokine levels and severity of illness although a causal direction is unclear from the literature we reviewed.

##### B3. Innate immunity-mediated mechanisms

###### B3a. Alternative complement pathway activation

In the non-COVID-19 literature, there is evidence of involvement of the alternative complement pathway in type III hypersensitivity. There is evidence of alternative complement pathway activation in COVID-19 and evidence for vasculitis and collapsing glomerulopathy.

###### B3b. Neutrophil extracellular traps

In the non-COVID-19 literature there is evidence of NET involvement in ARDS and a causal relationship with thrombosis. There are multiple lines of evidence in COVID-19 to suggest a role for NET’s in COVID-19-related coagulopathy as well as indirect lines of evidence such as the prognostic neutrophil-to-lymphocyte ratio.

##### B4. Adaptive immunity-mediated mechanisms

###### B5. Antiphospholipid Antibody Syndrome-Like Coagulopathy

There is strong evidence of the thrombogenic potential of antiphospholipid antibody (Hughes) syndrome. In COVID-19 there is no evidence we have seen of persistently elevated antiphospholipid antibodies but instead there is strong evidence for transient elevation. There is also the possibility that this transient elevation may be thrombogenic and represents an antiphospholipid antibody-like syndrome.

##### B6. Immunothrombosis

There is strong evidence of immunothrombosis mediated by platelets both in the non-COVID-19 and COVID-19-related literature. In terms of Virchow’s triad, this mechanism provides an important relationship between the vasculature and coagulopathy.

##### B7. Type-III hypersensitivity

There is strong evidence in the non-COVID-19-related literature of a type-III hypersensitivity response manifesting as vasculitis which in turn is thrombogenic. There is strong evidence in COVID-19 of a vasculitis and a type III hypersensitivity reaction.

##### B8. Sepsis-induced coagulopathy

In the non-COVID-19-related literature there is strong evidence of a sepsis-induced coagulopathy and this is the basis for the SIC score. There is evidence of an elevated SIC score in COVID-19 although this still requires a clarification of the mechanisms unless there is overlap with other mechanisms described here.

##### B9. DIC

There is strong evidence in the non-COVID-19-related literature of a relationship between DIC and both ARDS and sepsis and we have provided evidence of this in COVID-19 from our analysis.

### C. Hypercoagulable state

#### C1. Hyponatraemia

In the non-COVID-19-related literature there is evidence of an association between hyponatraemia and stroke and it has been suggested that the syndrome of inappropriate ADH secretion is a mediator. There is significant evidence of hyponatraemia in COVID-19 and evidence of SIADH in COVID-19 together with likely disruption of the RAA axis. Aldosterone and ADH are typically secreted simultaneously.

#### C2. Hypokalaemia

In the non-COVID-19-related literature there is evidence of an association between hypokalaemia and cardiac arrhythmias which in turn can be thrombogenic. In COVID-19 there is evidence for hypokalaemia and many authors suggest the involvement of the RAA axis as an explanation.

#### C3. Hypocalcaemia

There is evidence in the non-COVID-19-related literature of an association between hypocalcaemia and stroke risk. There is also evidence of Vitamin D in COVID-19 as a vulnerability factor. There is evidence of an association between hypocalcaemia in COVID-19 and severity of illness and an association with D-Dimers.

#### C4. Dehydration

There is strong evidence in the non-COVID-19-related literature between dehydration and stroke risk or a coagulopathy. There is strong evidence in COVID-19 for fever and diarrhoea although we did not find direct evidence of dehydration. Indeed the hyponatraemia and evidence for SIADH as well as disrupted RAA axis suggests this relationship may be complex.

#### C5. Immune-mediated mechanisms (B2-B10)

#### C6. Angiotensin II

There is evidence in the non-COVID-19-related literature for a thrombogenic potential of Angiotensin II. In COVID-19 there is evidence of elevated Angiotensin II although the disruption of the RAA axis suggests that impairment of homeostatic mechanisms is likely to make this relationship less than straightforward particularly with the finding of hyponatraemia.

#### C7. Clotting cascade

The clotting cascade represents the central aspect of the coagulopathy and there is abundant evidence in COVID-19 of elevated fibrinogen and D-Dimers of which we have provided evidence. There is also evidence of elevated von Willebrand factor.

#### C8. Hypoxaemia

There is a strong evidence base for a procoagulant effect of hypoxaemia and hypoxia and strong evidence of this in COVID-19 particularly with ARDS.

#### C9. Hyperviscosity

There is an increased risk of thrombosis in the hyperviscosity syndrome of Waldenstrom’s macroglobulinemia independent of COVID-19 and there is some evidence of increased hyperviscosity of unknown aetiology in COVID-19.

### D. System Involvement

#### D1. Cardiovascular

##### D1a. Vasculature

###### D1a1. Thrombotic Microangiopathy

There is strong evidence of primary and secondary microangiopathy in the non-COVID-19-related literature. There is also strong evidence of microangiopathy in COVID-19.

###### D1a2. Endotheliitis

The vascular endothelium can be considered as antithrombic and disruption prothrombotic. There is indirect evidence of disruption of the glycocalyx in COVID-19 and there is also strong evidence of endotheliitis in COVID-19.

###### D1a3. Vasculitis

There is a well-established relationship between vasculitis and thrombosis and there is strong evidence of vasculitis occurring in COVID-19.

##### D1b. Myocardium

###### D1b2. Cardioembolism secondary to cardiac injury

There are various types of cardiac injury and these can predispose directly or indirectly to thrombogenesis including through arrhythmias such as atrial fibrillation. There are a number of cases of cardiac thrombi in COVID-19 we identified but we did not identify evidence to suggest a direct link with cardiac injury.

###### D1b3. Secondary to cardiogenic shock

There is evidence in the non-COVID-19 literature of an association between cardiogenic shock and coagulopathy. In COVID-19 there is evidence of cardogenic shock which is prognostic and associated with evidence of SARSCOV2 myocardial invasion.

###### D1b4. SARS-COV2 myocarditis

There is evidence in the non-COVID-19-related literature of an association of myocarditis with a hypercoagulable state and there is evidence of myocarditis in COVID-19.

###### D1b5. Stress cardiomyopathy

There is evidence of an increased incidence of thromboembolic events in Takotsubo syndrome in the non-COVID-19-related literature. In the COVID-19-related literature we identified a case report of Takotsubo syndrome in COVID-19 with atrial fibrillation.

###### D1b6. Atrial fibrillation

The relationship between atrial fibrillation and thromboembolism is well-established in the non-COVID-19-related literature. There is evidence of an increased incidence of atrial fibrillation in COVID-19 particularly in severe COVID-19.

##### D2. Respiratory

###### D2a1. Respiratory failure with hypoxia leading to myocardial injury

Hypoxic injury leading to myocardial injury is well established and there are various sequelae including arrhythmias that can lead to thromboembolic events. There is evidence of myocardial injury being associated with ARDS in COVID-19.

###### D2a2. DIC

DIC is by definition a procoagulant condition which is well established in the non-COVID-19-related literature and we have provided evidence of DIC in this paper.

###### D2a3. Mechanical ventilation-mediated coagulopathy

See H2.

###### D2a4. ARDS-related coagulopathy

There is a well-established ARDS-related coagulopathy in the non-COVID-19-related literature. In our analysis we have provided evidence of COVID-19 and non-COVID-19-related ARDS being associated with thromboembolic events although with a higher incidence in the former. There is a suggestion that ARDS in COVID-19 may be significantly different from non-COVID-19 ARDS, sufficiently so to merit a distinct construct but for the model we assume it is valid.

##### D3. Central Nervous System

###### D3a. Direct nervous system injury leading to coagulopathy

We found limited evidence for this in COVID-19 other than a case of necrotising encephalopathy and a case of encephalopathy preceding stroke.

### E. Factors Relating to mild COVID-19

#### E1. Immobilisation/Bed Rest

The relationship with thromboembolism particularly DVT is well-established in the non-COVID-19-related literature although we did not find specific evidence in COVID-19.

#### E2. Dehydration due to fever, diarrhea

See C4.

### F. Factors Relating to severe COVID-19

#### F1. Critical-illness encephalopathy

There is some evidence in the non-COVID-19-related literature with an association between critical-illness encephalopathy and coagulopathy but this is in the context of severe comorbidity and risk factors. We found limited evidence of an association in COVID-19 and there is overlap with D3a.

#### F2. Stress cardiomyopathy

See D1b5.

#### F3. Atrial Fibrillation

See D2f.

#### F4. Metabolic/electrolyte disturbance

See C1-C4.

### G. Receptor-mediated

#### G1. ACE2-receptor-mediated pathways

### H. Iatrogenic

#### H1. Pharmacological adverse events

There is well-established evidence for a procoagulant effect of certain medications although we did not find evidence of this occurring in COVID-19 in the literature we reviewed.

#### H2. Medical Device-related clotting events

There is well-established evidence for clotting events with ECMO and we found evidence of thromboembolic events occurring with the use of ECMO, dialysis, with a previously patent aortic graft and with upper limb catheters. This is likely to result from a combination of hypercoagulability particularly with fibrinogen being concentrated in the ECMO and also with disruption of the vascular surface with catheterisation.

### I. Vulnerabilities

There are well-established vulnerabilities in COVID-19 although they are with prognosis rather than with coagulopathy specifically. A number of risk factors relate to endothelial dysfunction and we found evidence of case reports of thromboembolism in conditions which are associated with coagulopathy independently of COVID-19 and we propose a two-hit hypothesis.

## Discussion

We will now discuss our results, dividing this up into a general discussion of the findings, gaps in knowledge, a model of COVID-19-related coagulopathy, drawbacks of the study and then the next steps followed by a summary.

### General Findings on the nature of the COVID-19-related Coagulopathy

In our mixed methods scoping review, we were able to identify clinical evidence of a COVID-19-related coagulopathy. We found evidence of ARDS, sepsis and disseminated intravascular coagulation in cases with thromboembolic complications. However we also found evidence of cases where ARDS, sepsis or disseminated intravascular coagulation had either not been confirmed or else had been excluded. Since ARDS, sepsis and disseminated intravascular coagulation have well-established causal links with thromboembolic events ARDS, disseminated intravascular coagulation and sepsis are likely to account for a subset of the thromboembolic events seen in COVID-19. Given the evidence for additional mechanisms associated with coagulopathy, we can demonstrate more generally that COVID-19 is a polycoagulopathy mediated by multiple coagulation mechanisms (MCM’s).

The mechanisms for sepsis-induced coagulopathy (SIC) were unclear although there are criteria for the identification of SIC. Nevertheless (Liao et al, 2020) found that sepsis-induced coagulopathy typically preceded DIC. The non-overt DIC criteria may similarly be useful in COVID-19. Pulmonary thromboinflammation has been suggested as one of the main mechanisms leading to respiratory failure in COVID-19 (Bhattacharyya et al, 2020) and although undoubtedly important, the relationship of the pulmonary thromboinflammation to the endothelial inflammation elsewhere is not yet clear although it most likely results from viraemia.

We found that pulmonary arterial embolism was one of the most frequent thromboembolic events. (Benito et al, 2020) found an incidence of PE’s in COVID-19 of 2.6% in a cohort of patients in Barcelona. (Mackman et al, 2020) report on the incidence of thrombosis in COVID-19 in various studies and find rates differ in ICU and non-ICU settings. In ICU settings, the rates are VTE (4.8-69%), PE (16.7-35%), DVT (0.5-69%), arterial thromboembolism (3.8%), ischaemic stroke (2.7%). In non-ICU settings the rates are VTE (0.9-6.5%), DVT (0-46%).

Thromboembolism is unlikely to spare any vascular territory in COVID-19 although the pulmonary vasculature is likely to be commonly affected given the transmission dynamics of SARS-COV2 together with the anatomical considerations of the pulmonary vasculature. (Natelello et al, 2020) found evidence of microthrombi and microhaemmorhages from analysis of nailfold capillaries in patients with COVID-19 or else recovered from the acute illness. (Borczuk et al, 2020) found the pulmonary vasculature was commonly involved in COVID-19 in their autopsy cohort and this include macro and microthrombi. (Patel et al, 2020) found evidence of reduced pulmonary perfusion in COVID-19 and suggested this as indirect evidence of thrombosis. (Chi et al, 2020) found 23.9% of patients in 11 cohorts developed VTE.

Our findings suggest that there is evidence that the thrombi formed in COVID-19 may have unusual properties which merit further investigation. (Kalinskaya et al, 2020) investigated patients with COVID-19-related coagulopathy and compared them to a control group. They used methods including rotational thromboelastometry to characterise the clots and found that in patients with COVID-19 compared to the control group they grew more quickly, to a larger size and were lysed more quickly also which fits with a number of the themes identified in the abridged thematic analysis.

We suggest the construct of ‘residual COVID’ to describe residual irreversible ischaemic damage from thromboembolic events. Given the degree of plasticity of organs such as the brain, the necessary investigation of this construct is required but is relevant in the context of the possibility of sub-clinical thromboembolism in the acute phase of the illness. The transience of the coagulopathy in COVID-19 also requires clarification. (Garg et al, 2020) report a case of a patient who had recovered from COVID-19 but developed pulmonary emboli despite treatment with warfarin although they suggested that the PCR result may have been a false negative.

### Virchow’s Triad in Relation to the COVID-19-related Coagulopathy

The application of the model of Virchow’s triad to the COVID-19-related coagulopathy highlights an important distinction between the arterial and venous system. Arterial thromboembolic events are usually a result of many decades of pathology, predominantly through the development of atherosclerotic plaques. Deep vein thromboses however can occur relatively acutely and more frequently and this reflects the importance of stasis. The veins are valved and contrast anatomically with the elastic, pulsatile arteries and the blood is more prone to coagulation in the venous system with reduced mobility. With COVID-19, we see this pattern is reversed and arterial thromboembolic events occur relatively frequently and acutely even in the absence of plaques.

There are two factors that are likely to account for this. Firstly there is the marked hypercoagulability. This has been commented on by nearly all of the authors in the 56 main papers and is epitomised by the clotting that occurs in the ECMO centrifuges and dialysis machines. Although clotting is seen in the ECMO centrifuge in non-COVID-19 cases, this was noted to far exceed the degree of clotting that is expected and cannot be attributed either to stasis or to endothelial involvement except via hypercoagulability. The effect of the hypercoagulability is to accentuate the usual coagulation processes. Secondly there is also involvement of the vasculature with combination of different pathologies that have been identified representing the endothelial involvement in Virchow’s triad.

Another finding from considering the COVID-19-related coagulopathy through the prism of Virchow’s triad is that cardiac involvement may represent an important aetiology for stasis. This can be mediated via atrial fibrillation as an example and the importance of myocardial involvement in COVID-19 is becoming apparent.

### The Significance of the ACE-2 Receptor

SARS-COV2 requires the ACE-2 receptor for entry into the cells. From our literature review, we identified evidence of a potentially significant role for the ACE-2 receptor in terms of the COVID-19-related coagulopathy. Firstly there is the location of the ACE-2 receptors which determines which tissues are directly involved. The involvement of the respiratory and cardiovascular systems are particularly important in this regards. The second point relates to the effect on Angiotensin-II which is thought to be upregulated. Angiotensin-II has many effects including vasoconstriction and is thrombogenic. Thus by gaining entry to the cells via the ACE-2 receptor, SARS-COV2 sets in process a chain of events leading to an increase in the levels of circulating Angiotensin thereby potentially contributing to hypercoagulability of the blood.

### Mortality

The in-hospital mortality although high is in keeping with other studies (Vena et al, 2020). The hospital sample likely represents the most severely unwell patients. We have thus characterised a syndrome that can occurs in severe cases although this does not negate this occurring in cases that are classed as less severe and this is discussed further in the next section. Whilst we had a small sample population from the subset of 34 studies there were a number of findings that would benefit from further replication studies. We found a particularly high mortality in association with acute limb ischaemia and it was not clear to us why this was. For other thromboembolic events such as strokes and STEMI’s there are well-established care pathways informed by evidence-based policies. Given the high infection rate of COVID-19 even rare consequences of the infection will result in high case numbers due to the size of the infected population.

### Characteristics of the Population

The sample in the 56 main papers were all hospitalised patients and given the high mortality it is reasonable to assume that they represent the most unwell patients with COVID-19. Not all severely ill patients will be hospitalised due a variety of factors depending on the local healthcare provision which varies significantly internationally. Not all patients seen in hospital may be severely ill. Additionally thromboembolic/ischaemic events may be subclinical. The question of how relevant the syndrome is to the wider population of all patients with SARS-COV2 infection would require additional information. This information could be gained retrospectively but ideally with a prospective cohort study with a comprehensive sequential assessment for thromboembolic events through the course of the acute illness.

### Abridged Thematic Analysis

Thematic analysis is a method used in the analysis of qualitative data which can utilise both qualitative and quantitative approaches. There are three main approaches to thematic analysis: Coding book approach, coding reliability and reflexive/organic (Braun and Clarke, 2013). All of these approaches have one thing in common – the coding step which is considered central to the contemporary thematic analysis approach. Although thematic analysis has been references as far back in the 1950’s in psychoanalysis (Winder and Hersko, 1958) and further back in other fields, the modern origins of thematic analysis are cited as ‘The Thematic Origins of Scientific Thought’ by Gerald Holton (Holton, 1973). In this work, Holton examines key scientific theories to understand how they were developed. Holton’s sense is that there are certain aspects of theory development that occur in the minds of scientists even before they begin their work and he developed an approach to try and understand these aspects. (Merton, 1975) notes that Holton appears to have used an inductive approach to the thematic analysis. Thematic analysis is a flexible method for analysis that has been used with a variety of different approaches including philosophical underpinnings.

(Yardley, 2000) draws attention to the variety of qualitative methodologies together with a diversity of philosophical underpinnings whilst also recognising the importance of pluralism and the risks around evaluation. Whilst (Yardley, 2000) points out that there are objections to a universal code of practice of qualitative research, (Greenhalgh and Taylor, 1999) maintain that qualitative research should utilise “explicit, systematic and reproducible methods”.

The abridged thematic analysis (ATA) solved an important problem in analysing the content of a large number of papers with limited resources. The absence of the coding depends on an intuitive understanding of the material and a trust in the rater to identify the salient themes. We based the removal of the coding step on the philosophy of pragmatism, epitomised in the works of Dr William James who graduated from medical school and later-founded the American school of Psychology as well as writing works on the philosophy of pragmatism (James, 1907). We argue that in a pandemic, there is a strong ethical imperative to utilise and make available a methodology for rapidly analysing clinical literature to gain invaluable insights and generate understanding and testable models. Ironically it is pragmatism that led to the school of behaviourism which in turn led to response items in psychological testing which in turn can be viewed as eliciting a reactionary approach in terms of the reflexive approach that can be used in thematic analysis. The coding approach becomes impractical in the analysis of large numbers of studies and sampling is not appropriate as we had to examine all of the identified studies to extract the themes we arrived at. Thus pragmatism has provided the philosophical underpinning for the carefully considered step of removing the coding step although there are further considerations in terms of ATA and Validated Abridged Thematic Analysis (VATA).

ATA is justified by analysing material which is linked to an established academic field. Whereas the usual aim of thematic analysis is to describe the themes within the material, our further aim is to gain insights into COVID-19 in the midst of a pandemic. This further aim provides resource constraints including time and also the limited resources we had available to undertake the analysis. The analysis of the clinical material is most likely to be undertaken by frontline clinicians who are treating patients and who in the absence of large scale research institution support would work with limited research resources. ATA is a suitable tool in these situations, meeting the need for clinicians to rapidly develop an expert clinical knowledge base to provide care for those with COVID-19. In a more recent study (Janardhan et al, 2020) confirmed a subset of the findings from our abridged thematic analysis including COVID-19-associated clots being refractory and susceptible to rethrombosis and reocclusions and these findings provide some validation of our approach. They also suggest the primacy of the coagulopathy in COVID-19.

Turning to VATA, we needed to pivot firstly from understanding the perspectives of clinical experts in various fields and then hold in mind that our aim was to understand a viral illness. We therefore needed to validate our taxonomy which is the structuring of the themes we identified and of the themes themselves against the wider perspectives of experts across diverse fields. Whilst this may seem an ambitious undertaking, there is a simple reality here which is that all frontline clinicians in a pandemic are faced with providing care for people affected by a novel infectious agent with potentially serious outcomes. In medicine this applies without exception to all medical practitioners (e.g. acute and general physicians, psychiatrists, general practitioners and surgeons). This also applies to nurses, allied health professionals and all of the other essential staff in hospital and community settings and it is imperative that clear and accurate information is provided to patients and the public. COVID-19 is a multi-system disorder which crosses isolated areas of specialist practice and presents not only a significant clinical challenge but an immensely complex theoretical challenge. This is a time when silos of knowledge and training must be restructured to allow for rapid joined-up learning to enable us as a health and social care community to face a common and serious challenge and further to allow us as a global community to face this challenge with the conversations and debates that are so essential. VATA enabled us to gain insights from a diverse range of clinical experts that we ourselves would never have gained otherwise.

### Gaps in Knowledge

There are many knowledge gaps identified from this scoping review and we have included a selection of specific knowledge gaps that may help to inform future studies.

#### A. What is the role of the microvasculature in COVID-19?

(Tibirica and De Lorenzo, 2020) suggest a number of approaches to investigate the microvasculature and microvascular flow in critically ill patients with COVID-19 including laser Doppler perfusion monitoring, hand-held sidestream dark field imaging and reactive hyperemia-peripheral arterial tonometry.

#### B. Does splenectomy predispose to increased severity of COVID-19 and if so is this mediated via coagulopathy?

The spleen is a secondary lymphoid tissue. From the (Williamson et al, 2020) study there was a suggestion that splenectomy is associated with a higher risk of death in COVID-19 although this did not reach significance. There is evidence that COVID-19 can result in pathological changes in the spleen. There is also evidence that post-splenectomy there is an increased risk of thrombosis. This suggests that understanding the role of the spleen in COVID-19 may shed further light on aspects of the COVID-19-related coagulopathy.

#### C. Is coagulopathy a feature of Long COVID-19?

Does coagulopathy persist in COVID-19 after the acute illness has ended?

#### D. Are different configurations of ECMO associated with different risks of thrombosis?

There are different permutations such as venous-arterial and venous-venous. Do the different configurations have different risks of thrombosis given the anatomical distinction between the veins and arteries and the nature of any COVID-19-related vasculopathy.

#### E. Do drugs that act on the renin-angiotensin-aldosterone system (RAS) reduce the state of hypercoagulability?

As there is a potentially significant role for Angiotensin-II in COVID-19 and in relation to hypercoagulability, is there evidence that drugs acting on the Renin-Angiotensin-Aldosterone system affect the relevant coagulation parameters?

#### F. What is the role of Endothelial stabilising agents in COVID-19?

(Weinbaum et al, 2020) review the role of glycocalyx in various vascular-related diseases and note that the glycocalyx is a target for influenza viruses as well as SARS-COV. They also review a number of therapeutic agents for stabilising the glycocalyx. (Wilson et al, 2020) note that Adrenomedullin plays an important role in stabilising the endothelium in infection and call for an investigation of the role of Adrenomedullin in COVID-19. One study found that Adrenomedullin RNA was increased in patients with severe COVID-19 compared to mild illness (Hupf et al, 2020).

#### G. Is there an association between mesenteric ischaemia and mesenteric fat?

(Kaafareni et al, 2020) have presented interesting findings in a case of mesenteric ischaemia which was accompanied by a pattern of yellow tissue overlying areas of ischaemia. This can be seen clearly in the paper by (Gartland and Velmahos, 2020).

A case report by (Pang et al, 2020) may be of relevance here with the association of a congenital band associated with mesenteric ischaemia where they had earlier identified strands of fat. (Stanifer et al, 2020) draw on experimental findings to suggest that intestinal cells are an active site of SARS-COV2 replication.

(Ha et al, 2020) refer to ‘creeping fat’ and note the different functions of adipose tissue. They note the description of ‘creeping fat’ in Crohn’s disease by Crohn in 1932 and note that it may not only be an extra-intestinal repository for mucosa-associated gut bacteria but also that ileal inflammation may increase intestinal permeability. This in turn would facilitate the translocation of intestinal bacteria to extraintestinal locations and may explain why ‘creeping fat’ is associated with underlying strictures and inflammation.

(Coffey et al, 2018) created a metric for mesenteric pathology which included ‘fat wrapping’ which is equivalent to creeping fat. They found a significant correlation between this metric (mesenteric disease activity index) and the Crohn’s disease activity index and argued for inclusion of mesentery in ileocolic resection for Crohn’s disease. There is evidence for the involvement of adipose tissue in inflammation and immunomodulation (Mao et al, 2019)(Eder et al, 2019).

This raises the question of whether Crohn’s disease with GI involvement is a special case of COVID-19 with a potential immunological role for the fat wrapping, whether this is consistently associated with mesenteric ischaemia and if there is passage of the virus from the intestine into the peritoneal cavity and a disruption of the barrier function of the intestinal vascular endothelium.

## Limitations of the Study

There were a number of drawbacks in the study. (Makin and Orban de Xivry, 2019)

1. **The search strategy to identify the main papers was not rigorous**. We used a few search terms and the search strategy was not sophisticated. We justify this by the balance of getting quick answers in a pandemic and also by our limited resources. Had we detected a larger number of papers then the analysis would have taken much longer and so this offered a pragmatic solution. Despite the simplicity of the search strategy, we did identify relevant papers which provided a range of useful results. The less rigorous approach to the search strategy was justified by our novel methodology which enabled us to extract useful information from the papers and to validate this using a further literature search and review.
2. **The search terms in the main search may have biased the outcome**. We used search terms which could have biased the study towards identifying thromboembolic events in specific anatomical locations such as strokes and pulmonary emboli. Despite this, we identified a number of aortic and medical-device-related thromboembolic events. This together with the identification of a number of cohort studies suggests that our search strategy was successful in identifying a range of relevant pathologies which were not restricted to the search terms. This resulted in part from the broad nature of the search terms. Additionally the search terms provided only one of the databases indices for the returned papers. A wider search using more anatomical locations may yield further results and there is evidence of this from the subsequent identification of an ophthalmic artery embolic event although this does not negate the positive findings from the conducted searches.
3. **The search strategy for exploring the various hypotheses for the aetiology was unstructured and prone to bias**. We acknowledge the limitations of the unstructured search strategy. The goal of this search was not to be definitive in answering questions but primarily to explore and test the various hypotheses that had been generated as the validation stage of the validated abridged thematic analysis (VATA). The hypotheses are diverse and cover many theoretical domains and had we not utilised a pragmatic approach this would not have been possible. The search strategy subserved the goal of continuing the qualitative analysis by triangulating with other sources in the same knowledge domains. There is a widely recognised phenomenon of positive findings being published in the clinical literature which could have biased us towards overestimating the strength of evidence for a hypothesis. Whilst we acknowledge this we would point out that certain hypotheses were discounted by this process and that we propose a methodology for extending the model which would enable others to correct the errors resulting from biases with more rigorous approaches. Additionally we had also included systematic reviews and meta-analyses which typically address these issues.
4. **In the qualitative analysis, the absence of coding increases the risk of omissions from the identified themes or else misinterpretation**. The omission of coding was a pragmatic decision which is based on theoretical considerations as outlined (see also the section on philosophical considerations). Whilst it may be prone to omissions and misinterpretation, the meaning we have sought is in established knowledge domains and we have triangulated this with other research publications when looking at aetiologies to determine content validity. This method of confirming content validity is not present in other approaches we have seen even when they have included coding. Whilst there is a risk of omission due to oversight, we have judged the benefits to outweigh the risks in terms of rapidly summarising themes from a large number of papers where this would not have been practically possible with the usual coding process. Again we have provided a strategic methodology that enables other researchers to address any shortcomings in our work and to thereby improve the overall understanding.
5. **The patients in the 56 main papers were all hospital-based and therefore the findings do not generalise to all patients with COVID-19**. We acknowledge for the quantitative aspects of the study and the qualitative analysis of the clinical features of the thromboembolic events that the patient sample were people with more severe COVID-19. However a number of patients had experienced thromboembolic events after a period of being asymptomatic or having mild symptoms. Additionally the VATA and model-building drew on a wider literature base. Although not generalisable to all patients with COVID-19 we have characterised a syndrome that results from COVID-19.

## Hypotheses

Here we outline hypotheses generated from the findings. The purpose of this section is to provide clinicians and scientists with hypotheses for further testing. We provide comments on the strength of the evidence informing these hypotheses.

**Hypothesis 1:** *COVID-19 causes an arterial vasculopathy that promotes a hypercoagulable state*.

Our findings revealed a significant difference in the frequency of reported venous and arterial thromboembolic/ischaemic events in the selected papers we identified. An adult case of Kawasaki-like disease in COVID-19 has been reported (Shaigany et al, 2020) and we have reviewed the evidence for endothelial dysfunction, endotheliitis and vasculitis all of which suggest the involvement of the vasculature in the pathogenesis of COVID-19. The above hypothesis does not negate the presence of venous thromboembolism but instead refers to the strong evidence for arterial thromboembolism. In so doing, we have also taken the unusual step of including the pulmonary arteries as separate from the venous circulation purely on the grounds of the anatomical structure of the arteries which shares an embryological origin with the aorta. This distinction is important in terms of the location and density of the ACE-2 receptors in smooth muscle cells in arterial walls in comparison with the veins as well as the difference in blood flow dynamics and shear stress.

**Hypothesis 2:** *COVID-19 is associated with arterial thromboembolic events in five main 5 territories – mesenteric ischaemia, cerebrovascular events including the carotid artery involvement, myocardial ischaemia/infarct, pulmonary emboli and acute limb ischaemia* From the 56 main papers, we found evidence for thromboembolic events in all of these territories. This does not negate the occurrence of thromboembolic events in other arterial territories including the aorta but it emphasises clinically important locations with evidence from the literature. In most of the studies there was not a control group although there was indirect evidence of an increase in local incidence and where there were control groups this provided further supportive evidence. The occurrence in these territories would have implications for diagnostics, community and hospital surveillance and public health campaigns and it is therefore important to test this hypothesis further. Systematic reviews or meta-analyses could investigate the question of incidence, the involvement of other territories as well as the likelihood of diagnosis in different settings. Further case studies and case series would be invaluable in providing further supporting evidence, data on key variables as per the recommendations section and also providing additional clinical acumen for qualitative analyses.

**Hypothesis 3:** *The arterial ischaemia pathology in the 5 main territories identified in COVID-19 cases is associated with high mortality*.

From the analysis of 34 papers, we found that the arterial ischaemic pathology in the 5 main territories was 7.6-35%. The overall mortality is much higher than expected in COVID-19 and suggests that the sample is not representative of the total population of people who develop COVID-19 (we include asymptomatic people in this generalisation). This may be due to selection bias in the cases that have been selected for publication all of whom are in a hospital setting. However the arterial ischaemic pathology may be less likely in the total population than the hospital setting alone.

**Hypothesis 4:** *COVID-19 predisposes to large vessel stroke particularly involving the middle cerebral arteries*.

From the analysis of 34 papers, we found that stroke occurred in 27.7% of cases. This owed partly to the selection bias from the search strategy. Of those cases that were reported, the majority were large vessel strokes predominantly affecting the middle cerebral arteries. Additionally as this is a select population in a hospital setting with more severe COVID-19, the significance of the percentage of cases is limited to this setting and severity. However (Oxley et al, 2020) draw attention to the unusually high incidence of large vessel stroke in young patients in their case series and find a large increase in comparison with other time periods. Additionally a number of patients presented after a period of being asymptomatic in the community.

**Hypothesis 5:** *The carotid arteries may be a source of large vessel thromboembolism in COVID-19-related stroke*.

From the analysis of the 56 main papers, we identified a number of cases affecting the carotid arteries even in the absence of plaques and this together with the location is of significance. Further case reports would be helpful in identifying any associations such as cardiac emboli, evidence of direct viral invasion of the carotid endothelium or physiological evidence of involvement of the carotid body. Additionally it has been suggested that in the case of large vessel strokes, carotid involvement is excluded in COVID-19. Competing hypotheses would include carotid artery involvement as an incidental finding or extension of vascular involvement to neighbouring arterial territories (e.g. middle cerebral artery) consistent with a vasculopathy.

**Hypothesis 6:** *Thrombi extend into multiple arterial branches producing multi-territory ischaemic events including multi-territory stroke and desert foot*.

From the qualitative analysis of the 56 main papers, we identified multi-territory involvement in the same patient. These findings were commented on by authors. Further evidence would be needed in the form of case reports and estimation of prevalence.

**Hypothesis 7:** *Thrombi have a high risk of embolisation compared to thrombi resulting from common thrombogenic pathologies*.

From the qualitative analysis of the 56 main papers, one group reported a high risk of embolisation during mechanical thrombectomy in large vessel stroke confirmed by different surgeons and using varied techniques. If the thrombi have a high risk of embolisation then in combination with a putative extension of vascular involvement along arterial walls this would increase the risk of involvement of multiple territories. Further case reports and quantification of the effect would provide further valuable evidence and the latter could be investigated using a meta-analysis or systematic review.

**Hypothesis 8:** *Women are more likely than men to die from pulmonary emboli in COVID-19*.

This hypothesis is based on an analysis of 34 studies and with a relatively small number of cases. This is not an a priori hypothesis and would need to be tested in a more robust way with a systematic review or meta-analysis or with original data and should be regarded with an abundance of caution until such evidence is available.

**Hypothesis 9:** *Men are more likely than women to die from acute myocardial infarcts in COVID-19*.

This hypothesis is based on an analysis of 34 studies and as with hypothesis 8 is not an a priori hypothesis,and is based on small numbers and should again be viewed with an abundance of caution.

**Hypothesis 10:** *Women are more likely than men to die from thromboembolic/ischaemic events involving the splanchnic artery territories in COVID-19*.

Again this is based on the analysis in the 34 studies and should be considered with due caution.

**Hypothesis 11:** *Women are more likely than men to die from strokes in COVID-19*.

Again this is based on the analysis in the 34 studies and should be interpreted with the caveats for hypothesis 8.

**Hypothesis 12**: *Women are more likely than men to die from acute limb ischaemia in COVID-19*.

This is based on the analysis in the 34 studies and should be considered with the caveats for hypothesis 8.

**Hypothesis 13:** *Women are more likely than men to die from aortic thromboembolic events in COVID-19*.

This is based on the analysis in the 34 studies and should be considered with the caveats for hypothesis 8.

**Hypothesis 14:** *Thromboembolic events are underreported in COVID-19*.

(Lodigiani et al, 2020) make this suggestion. Our findings from the analysis of the 56 main papers would support this hypothesis as we found that the average number of thromboembolic/ischaemic events in each patient was 1.3. There are a number of reasons why thromboembolic events may be underreported including changes in service provision during the pandemic, occurrence in the community setting and diagnostic overshadowing.

**Hypothesis 15:** *A subset of patients with Long-COVID will experience residual consequences of ischaemic lesions which we term Residual-COVID*.

Following on from Hypothesis 2, arterial thromboemboli in the five main territories would be expected to be associated with characteristic findings which include ischaemic events such as stroke. Whilst there is a degree of plasticity with stroke depending on a number of factors, there is also an element of irreversibility due to tissue destruction. We would term this irreversible loss of function as Residual-COVID as recovery of function would not be expected. Ischaemic lesions could be thus described on the basis of the established literature in other conditions. There may be other aspects of the pathology in COVID-19 that would fit into this category.

**Hypothesis 16**: *The two-hit hypothesis*

Based on our findings, we hypothesise that a pre-existing vulnerability to coagulopathy combines synergistically with COVID-19 to increase the risk of coagulopathy further.

## Next Steps

The purpose of a scoping review is to review the subject area in preparation for further research. In this regards, we divide the proposed next steps into three. Firstly there are the immediate recommendations for reporting in papers in this area so as to move towards a standardised approach which will facilitate data aggregation. Secondly we recommend specific processes that will facilitate a decentralised research program (DRP). Thirdly we make recommendations about developing a symbolic representation of causal links. This has a more general usage case than the COVID-19-related coagulopathy but this recommendation results from the problems that have been encountered both in terms of healthcare delivery in the pandemic as well as the challenges we have faced in writing this paper. This third type of recommendation requires a more profound change in practice and may not be achievable in the short-term but is particularly well-suited to meeting the challenges of a pandemic.

### Recommendations

A scoping review is limited in comparison with a systematic review or meta-analysis in the recommendations that can be made. Whilst we make no health policy recommendations, we are able to make recommendations specifically for the methodology of research publications in this area. This is justified as we have evaluated the methodology of a significant number of publications as reported here.

**Recommendation 1:** *Case reports or case series on clotting disorders in COVID-19 have a valuable role in advancing knowledge of COVID-19*.

In our paper, we have analysed the themes from the research literature including case reports and case series and have generated hypotheses for further testing on the basis of these findings. Case reports and case series are an accessible way for clinicians to communicate their findings in a timely way which is essential in the current pandemic. There has been criticism of case reports and case series as lacking the rigour of randomised controlled trials (RCT’s) (Jung et al, 2020). However as we have demonstrated, such publications can bring valuable insights that not even RCT’s may offer. Indeed the problems posed by the COVID-19 pandemic have disrupted research (Kimmel et al, 2020) and caused a reappraisal of the role of different types of evidence (Greenhalgh, 2020)

**Recommendation 2:** *Clinical researchers reporting on clotting disorders should use a standardised approach*.

In our paper, we demonstrated that there was significant heterogeneity in the reporting of clinical data resulting in incomplete datasets for variables of interest. For instance, the care setting (e.g. ICU) contextualises the clinical data and findings may not generalise to other settings. Therefore the absence of such data from clinical cases can pose a challenge for including the data in a systematic review or meta-analysis. The use of a standardised dataset facilitates the extraction of further insights from analysis of the aggregated data. We provide a template for reporting.

**Recommendation 3:** *We recommend reporting of individual data for large cohort studies. We found that case studies offered a rich albeit heterogenous source of data and with cohort studies, aggregation of data removed the possibility of secondary analyses of certain aspects of the data*.

**Recommendation 4:** *We recommend reporting the gender of the patient when reporting on COVID-19-related coagulopathy*.

We have detected gender differences in mortality for the different types of thromboembolic/ischaemic events and we would recommend reporting on gender.

**Recommendation 5:** *We recommend reporting on whether a diagnosis of ARDS was present or had been excluded*.

We demonstrated in our findings that a diagnosis of ARDS was not always mentioned in papers either as confirmed or excluded. Since ARDS is associated with a coagulopathy and has an important association with sepsis as well as specific treatment pathways, it is essential both in the interpretation of the clinical data and for the further analysis of aggregated data.

**Recommendation 6:** *We recommend reporting on ARDS using the Kigali modification of the Berlin definition in a non-ICU setting*.

ARDS is an important pathology to exclude or confirm when evaluating clotting disorders in COVID-19. Whilst this is mainly seen in the ICU setting, ARDS exists along a continuum and cases may develop in the general ward setting. The potential for silent hypoxia which has been described in COVID-19 reinforces the need for an exclusion of ARDS when reporting clotting disorders in COVID-19. Since the PaO2/FiO2 interpretation involves the positive end-expiratory pressure (PEEP) or CPAP values or which are typically available only in an ICU or HDU settings an alternative is required for general settings. The Kigali modification of the Berlin definition allows the calculations to be made using the ratio SpO2/FiO2 (Riviello et al, 2016) and is a simple practical tool for clinician when reporting on an important differential in a case report or case series.

**Recommendation 7:** *We recommend reporting on whether a diagnosis of Disseminated Intravascular Coagulation had been confirmed or excluded and including non-overt DIC where available*.

Disseminated intravascular coagulation is an important differential in COVID-19-related coagulopathy and a diagnosis inclusion or exclusion should be reported where available.

**Recommendation 8:** *We recommend reporting on whether a diagnosis of sepsis has been made*.

Sepsis is an important differential in COVID-19 and particularly in terms of coagulopathy.

**Recommendation 9:** *We recommend reporting on whether mechanical ventilation was used*.

The association of mechanical ventilation with pulmonary coagulopathy means that this is an important differential in any consideration of COVID-19-related coagulopathy. This also emphasises the importance of multidisciplinary publishing where the patient has been treated in multiple settings.

**Recommendation 10:** *We recommend reporting on the oxygen saturation at the time at which thromboembolic events are diagnosed*.

As hypoxia is a mediator of coagulopathy and a central feature of ARDS, the reporting of hypoxia in publications is important.

**Recommendations 11:** *We recommend publishing evidence of coinfection where possible*. There is circumstantial evidence for a contribution of co-infection to the COVID-19 related coagulopathy but this relationship has not been the focus of the literature we have reviewed. Further original research would be beneficial in exploring this relationship including whether co-infections were treated. From the identified literature there are putative candidates for organisms which have demonstrated relationships with coagulopathy.

**Recommendation 12:** *We recommend publishing evidence of cardiac structural or ECG abnormalities where available and specifying whether the pathology existed prior to the SARS-COV2 infection*.

The importance of cardiogenic shock and myocardial involvement in COVID-19-related coagulopathy has been discussed and would be an important differential.

**Recommendation 13:** *We recommend that pharmacologically active agents that may increase the risk of coagulopathy are included in the reported data in studies*.

This is a differential that should be included for the purposes of evaluation and further analysis.

**Recommendation 14:** *We recommend that data on the mobility of patients are reported in publications*.

In this regards, general comments on whether the patient was restricted to bed without activity throughout a period of hospitalisation or in the community are helpful in identifying an important aetiology.

**Recommendation 15:** *We recommend that blood pressure measurements are reported in publications*.

Given the associations of hypotension and hypertension in general with coagulopathy we would recommend the inclusion of data in the period leading up to and including the time of the thromboembolic events.

**Recommendation 16:** *We recommend that Fibrinogen levels are reported*. Fibrinogen levels are used in the diagnosis of DIC and evidence discussed previously demonstrates an elevation in COVID-19.

**Recommendation 17:** *We recommend that D-Dimer levels are reported*.

D-Dimers are used in the diagnosis of DIC and the evidence previously discussed demonstrates a significant elevation in D-Dimers in many cases of venous and arterial thromboembolism and the D-Dimers are prognostic in COVID-19. There has been discussion in the literature about standardising the reporting of D-Dimers (Lippi et al, 2015). We would recommend that the D-Dimers should be reported with the corresponding laboratory reference range, the units and whether this is expressed in D-Dimer units (DDU’s) or Fibrinogen Equivalent Units (FEU’s).

**Recommendation 18:** *We recommend that platelet levels are reported*.

Platelet levels are used in the diagnosis of DIC and platelets are central to coagulation.

**Recommendation 19:** *We recommend that prothrombin time is reported*.

Reporting the prothrombin time is useful given the role of prothrombin time in the diagnosis of DIC.

**Recommendation 20:** *We recommend that alternative complement pathway biomarkers are reported where available*.

There is emerging evidence of a role for the alternative complement pathway in COVID-19 and so further information on this will be very useful.

**Recommendation 21:** *We recommend that Protein C levels are reported where available. Protein C is useful for the evaluation of non-overt DIC which may be relevant in COVID-19 but where further information will be needed*.

**Recommendation 22:** *We recommend that Antithrombin III levels are reported where available*.

Antithrombin III is also useful for the evaluation of non-overt DIC which may be relevant in COVID-19 and further information is needed.

**Recommendation 23:** *We recommend reporting on IL-6 if available*.

IL-6 is key in many inflammatory processes that may be relevant to COVID-19 related-coagulopathy.

**Recommendation 24:** *We recommend reporting on the full extent of investigations to exclude venous and arterial thromboembolic events*.

Reporting of the full extent of investigations would be helpful in estimating the prevalence of venous and arterial thromboembolic events. In many of the studies, the patients are severely ill and the focus may be on managing other aspects of the presentation. Having an understanding of the extent of investigation for thromboembolic events would allow aggregated data to be analysed according to actively investigated arterial and venous territories.

**Recommendation 25:** *We recommend reporting on the location of the thromboembolic events*.

In a number of cases the location of ischaemic areas was reported rather than the arteries supplying those areas. Attempts to estimate the arterial territories are prone to errors due to anatomical considerations (e.g. circle of Willis connecting arterial territories) as well as individual anatomical variation although the latter is less likely to result in error.

**Recommendation 26**: *We recommend reporting the Dublin-Boston score if IL-6 and IL-10 levels are available*.

Given the controversies around the IL-6 levels, we recommend using the Dublin-Boston score given the prognostic value (McElvaney et al, 2020).

**Recommendation 27**: *We recommend using the checklist in Table 36 when publishing original data on COVID-19-related coagulopathy and refining the checklist where necessary*.

**Table 36.**
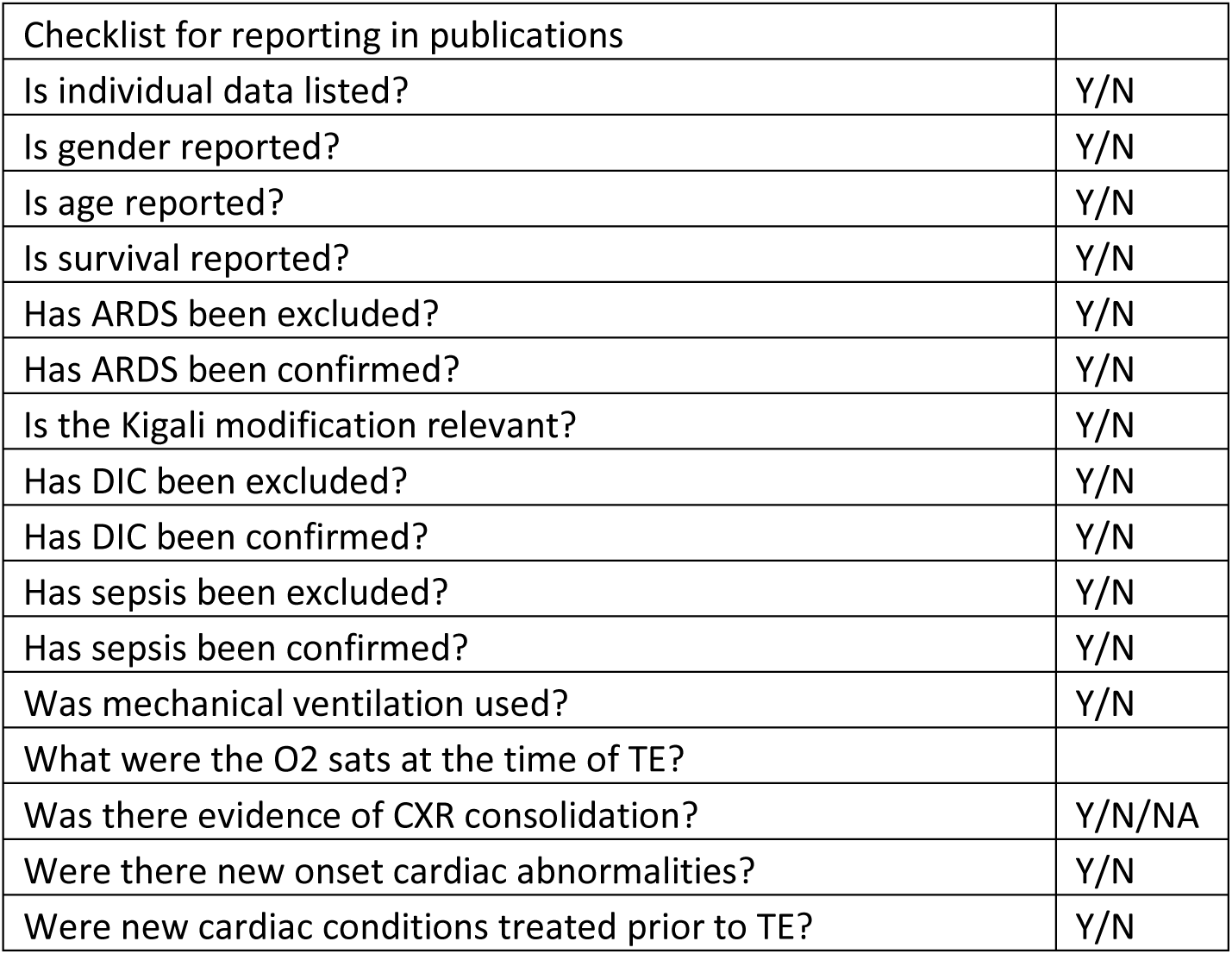
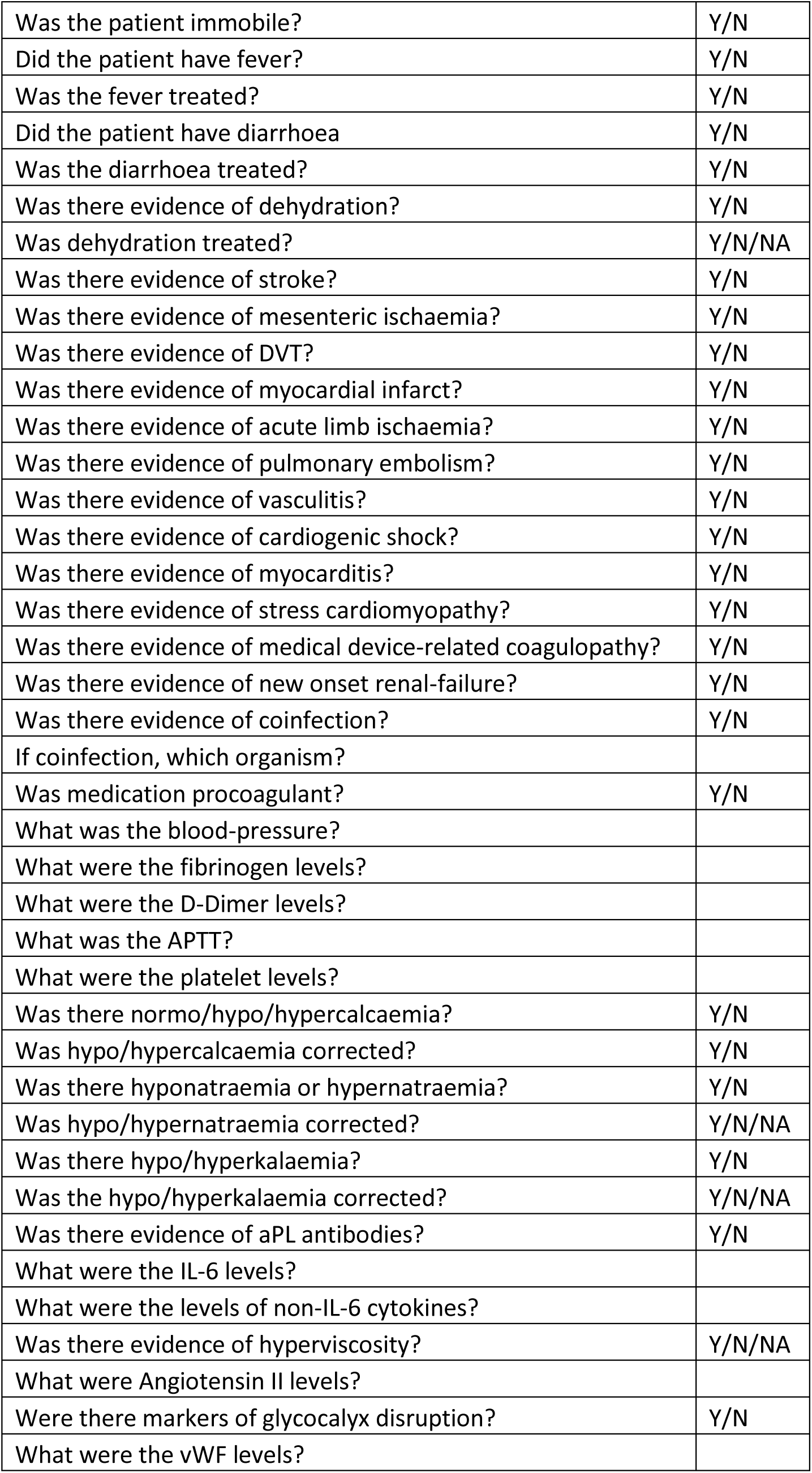
Checklist for reporting in publications, TE – Thromboembolic event.

The checklist is also provided in the supplementary data section.

### Crowdsourcing: A Model for a Decentralised Research Program (DRP)

We propose a model for a decentralised research program based on the approach in this paper. During the COVID-19 pandemic there have been a number of challenges. There have been profound impacts on research ranging from suspension of activities through to diversion of funding (Yang et al, 2020)(Jeon et al, 2020)(Keswani et al, 2020). Clinical staff have been redeployed to other areas of practice including those outside of the domain of their clinical training and services have been disrupted (Metgaloikonomos et al, 2020)(Payne et al, 2020)(Lim et al, 2020)(Coughlan et al, 2020))(Valente et al, 2020). Students have been graduated early in order to undertake clinical work during the pandemic (Sharif, 2020). COVID-19 is increasingly recognised as a complex multi-system disorder which spans multiple areas of clinical specialism and where a variable course gives rise to obscure pathologies in a small proportion of cases. Multisystem diseases, rare disease and diseases which are not well characterised are recognised as some of the most challenging to manage (Watts, 2014) (Volkova et al, 2020). Given the scale of the pandemic, a small proportion of cases translates into a very large number of actual cases.

Clinicians are on the frontline of this pandemic and the experience of clinicians is essential both in publishing findings and also translating research findings including trial outcomes into clinical practice. Our proposal focuses on the frontline clinician who is typically not well-resourced for research unless participating in large trials and who will have varying degrees of clinical research experience. We propose a crowdsourcing approach to facilitate COVID-19 research in the pandemic. Crowdsourcing approaches have already been utilised in a number of clinical areas (Thompson and Bentzien, 2020), (Si et al, 2020), (Lacourse et al, 2020) and with a crowdsourcing approach it is possible to equal or even surpass largescale well-resourced endeavours in terms of outcomes.

The crowdsourcing approach is organised through the underlying clinical model we have provided. The clinical model offers the fulcrum around which the research program is organised, negating the need for centralised oversight. Thus clinicians and researchers can identify part or all of the model to address and then update this aspect of the model at the conclusion of their input. In this manner, the clinical model can be viewed as a dynamic model which is in a state of continuous improvement from the clinical/research community. We would anticipate that after a few revisions, the clinical model may be unrecognisable from the original as the model more closely approximates reality with subsequent expert review as well as clinical and basic research findings.

### A Toolkit for Researchers

We outline briefly how clinicians and researchers can advance the model developed in this paper. We recognised that there are a great many talented clinicians and researchers around the world who would have insights, expertise and experience that would enable them to see flaws in our work. This is the nature of science and to expose these flaws and improve upon them is a measure of success in science for we approach ever more closely to the truth although possibly still only approximating. This matter is also complicated by the nature of systems biology.

We provide the outline of a toolbox which has the following components

1. ATA. The abridged thematic analysis is a simple approach which enables the clinician/researcher to rapidly identify COVID-19-related themes in the literature. This is outlined in the methodology section and the discussion sections in more detail. This can be combined with systematic reviews and meta-analyses to produce mixedmethods approaches. These in turn can inform policy and investigate more narrow domains of enquiry than we have considered here.
2. VATA. The validated abridged thematic analysis is an extension of ATA and involves the additional step of an exploratory literature search to validate the taxonomy generated by the ATA. The exploratory literature search can be refined and systematised as necessary and there is much scope for improving upon our approach.
3. Taxonomy. Our taxonomy for COVID-19-related coagulopathy is outlined in Table 35. This can be used in a number of ways. Thus a publication could be based solely on updating the whole or part of the Table on the basis of a literature review. If other taxonomies are available, then a publication could be based on the comparison of the taxonomies thereby deriving a new taxonomy based upon this comparison with justifications.
4. Diagram mapping. The diagrams offer hypotheses contained within the relationships which can inform new original studies. Diagrams can be updated with a review of the relevant literature although this overlaps with 3 since the taxonomy maps onto the diagrams. The rules of diagram mapping can be advanced and this again can form the basis of a publication as our rules are rudimentary and easily improved upon particularly in the area expanding the logical relationships. This would then inform other publications.
5. Hypotheses. The hypotheses can be tested with original data or else with reviews of the literature and form the basis for publications.
6. Gaps in Knowledge. The gaps can be systematically described with a corresponding taxonomy and prioritisation for the research community which we have not attempted here.
7. Recommendations. The literature can be reviewed to improve the recommendations that can be made for clinicians/researchers particularly in terms of the checklist.
8. Keys and locks Model. All of items 1-7 can be included in papers but particularly 3-7. In this manner a series of research papers can become interlocked with each other, one publication responding to another and a further publication responding to that one. A chain is thus formed and can be tracked through the references and be linked to successive revisions of the taxonomy or other aspects of the toolbox.
9. Aggregators. There may come a point at which there are several simultaneous versions of the taxonomy. At this point, there are risks around a decentralised research program. However this can be solved by recognition that it can be addressed by a designated role where an author compares the different taxonomies and thereby brings order, referencing the taxonomies and also providing a solution for the differences.
10. Tracking publications that use the toolbox. Papers could be shared using headings which identify the toolbox so that they are more easily found e.g. hashtags on social media platforms.

### Standardising the Communication of Pathological Mechanisms In COVID-19-related Coagulopathy

#### Developing Rules for Diagram Mapping in a DRP

In this scoping review, we have mapped a model of COVID-19-related coagulopathy onto diagrams. The diagrams provide an intuitive overview of the pathological mechanisms outlined in the model.

We used a number of simple rules for the construction of the diagrams. Although the diagrams appear rudimentary in comparison with other diagrams, the accompanying rules enable a standardisation of communication. The diagrams are symbols relating to underlying pathophysiology represented at different levels ranging from platelet activation to the more general concept of thromboinflammation. In some diagrams these have been linked to clinical signs as in the case of chilblains relating to the interferon response. By this means it is possible to represent the process from increased levels of circulating molecules through to the clinical features including signs and symptoms. We note that symptoms are a response of the mind to pathophysiological events and thus for any inclusion of symptoms or indeed psychological stressors or other factors the model would rightly be considered psychopathophysiological.

We have developed a simple set of rules for describing the relationships. Thus a simple line denotes an ‘example of’ rather than a causal relationship. We have also included causal relationships and colour coded them into weak, moderate and strong evidence both for COVID-19 specifically and separately for the more general non-COVID-19-related cases. By this simple set of rules, we are able to take our earlier analysis and translate it into simple diagrams which illustrate relationships between concepts.

Whilst this may appear straightforward enough, there is a further point to this in that the diagrams are not an end to themselves but rather as symbolic representations open up further possibilities related to a decentralised research program. As a scoping review, we consider this as a starting point. In future iterations the rule set can be modified thus offering various possibilities – they may be maintained, expanded or replaced but with the intention that they become generally more useful or else more useful for certain applications of the model. For example, the rule set can be expanded to include associations and thereby incorporate the findings from additional research studies or else allocate associations to existing causal relationships where they are considered more appropriate (e.g. elevated IL-6 and stroke).

#### The Utilisation of Induction in Diagram Mapping

In the diagram mapping, we have utilised non-COVID-19-related research to demonstrate relationships between concepts. In the COVID-19 pandemic, the medical community has been required to respond to a novel pathogen which can lead to severe illness without the advantage of years or decades of research into the treatment approaches. In this context it is possible to start with some assumptions about the virus and the subsequent illness based on knowledge of related conditions – in other words through the inductive process. The typical course of action when there is sufficient time and resources is to consider the matter from deductive principles based on the evidence of the phenomenon in question. With SARS-COV2, there is a well-established body of evidence from related conditions and principles derived from decades and even centuries of research into haemostasis, immunology and the presentation of clinically relevant thromboembolic events. There are significant difficulties in applying this body of knowledge to the treatment if the virus and subsequent illness behave differently than in related conditions. However from the perspective of a scoping review, the purpose is to inform research going forwards and induction provides a large number of hypotheses to test in SARS-COV2 that otherwise may not be apparent.

Therefore construction of a mixed deductive-inductive model for empirical observations in COVID-19 has the potential to realise a number of advantages over a deductive-only model. In our paper, this inductive model began in the process of validation of the abridged thematic analysis and translated into an intuitive visual format in the process of diagram mapping. In the inductive model we are generalising from non-COVID-19-related pathological mechanisms to COVID-19. This poses the risk of harm if this theory is applied to practice. However we are careful to limit the application of the inductive model in the sense that we are creating a highly theoretical model which is not directly informing practice including policy (particularly as this is a scoping review).

The purpose of the inductive model is to provide an underlying framework for understanding the COVID-19-related pathology and to generate hypotheses for testing in COVID-19. To give an example, there is evidence that Angiotensin II leads to thromboinflammation in nonCOVID-19-related research although we did not find this evidence in the literature we examined for COVID-19 although it is entirely plausible and can be extrapolated through induction. However if this were to form the basis for a therapeutic trial we may consider it too early to jump to such conclusions and this is the purpose of the diagram mapping. Instead the next step would be to formulate the hypothesis that Angiotensin II leads to thromboinflammation in COVID-19 and investigate this in the usual way through case studies, case series, meta-analyses, systematic reviews, narrative reviews and various types of trials or cohort studies. In so doing, we are able to utilise the vast amount of literature on the related areas to generate further hypotheses specific to COVID-19 rather than discounting this literature as irrelevant.

By using induction we may also generate a number of possible mechanisms as we examine each causal link. In this manner we may end up with a lot of ultimately irrelevant causal links. The diagrammatic mapping does not remove the irrelevancies but serves as a framework for communicating the reality of research findings which can be controversial, contradicted, irrelevant or turn up unexpected links to other areas of theory. Thus the mapping exercise simply gives us a way to visualise the complexity that alright exists but otherwise may not be so obvious. The question instead is how do we use the mapping to manage this complexity.

#### Symbolic Manipulation: A New Level of Analysis

The generation of symbolic diagrams may seem somewhat limited in application but it provides the clinician with the beginnings of an invaluable tool for clinical analysis. In clinical analysis, we are used to dealing with the analysis of laboratory results, imaging findings, symptoms and signs, other aspects of the clinical history as well as relating this to our readings of the clinical literature. The analysis involves direct clinical data or else a discussion of that data in text and diagrams. By developing this symbolic system we are able to perform a level of analysis which is two steps removed from the direct data. Whilst this can also be accomplished through spoken language, language is not designed specifically for this purpose but is instead a tool for generic application. In other words, the symbolic system is a specialised symbolic language designed with the intention of communicating clinical concepts and facilitating abstract analysis. A new level of analysis is possible which involves the manipulation of abstract symbols which signify clinical meaning. This opens up new avenues including the ability of the clinician to rapidly develop pattern recognition depending on the dataset and for the possibility of symbolic computation.

#### Identifying Areas of Further Research from the Diagram Mapping

As our scoping review was broad in nature, the diagrammatic mapping covers many subjects but to a limited depth. By inspection of the diagrams, the clinician or research is able at a glance to see areas that require improvement or with which they disagree.

#### Towards a Standardised Symbolic Clinical Notation

During the writing of this paper, we made a number of observations about the process of characterising the COVID-19-related coagulopathy.

1. Different explanatory mechanisms for the coagulopathy are posited by different authors
2. The explanatory mechanisms differed between clinical specialities in a number of cases reflecting different insights.
3. The explanatory mechanisms were described in language/text in the papers we examined.
4. Diagrams were used in many of the papers to support the text-based description of the mechanisms. The diagrams differed in their construction, the components which were illustrated and the relationships and this included descriptions of the same concepts.
5. In the process of diagram construction, there were a number of valid approaches that we could use to illustrate key concepts including relationships and this required investment of resources such as time.
6. As per the definition of pandemic, the international community is affected and at the time of writing there are 215 countries, territories and collectivities that have documented cases of COVID-19, with over 1 million deaths (Worldometer, 2020).
7. Internationally there are at least 7097 living languages (Ethnologue, 2020).
8. It is common practice in the clinical literature to limit literature searches to the language of the authors.
9. Theoretical practice forms the underpinnings of clinical practice including evidencebased medicine/practice (EBM) although the theoretical emphasis of EBM is on the evidence for interventions. The evaluation of evidence for underlying mechanisms typically falls within the remit of specialised scientific communities. Specialist scientific communities typically utilise their own specialist terminologies and paradigms (Kuhn, 1962).
10. Different specialities within medicine typically interface with the relevant scientific communities.
11. Specialist knowledge within clinical specialities and scientific communities together with language-bound knowledge creates knowledge silos. Knowledge silos act as barriers to developing an understanding of multi-system clinical disorders where an oversight of knowledge is required. Having an oversight of knowledge can be likened to having a map of knowledge which enables orientation.
12. COVID-19 is a complex, multi-system disorder which involves changes in the host immune and haemostatic responses. The immune and haemostatic systems communicate with each other and can themselves be associated with multi-system disorders. Additionally there is evidence of direct viral injury to multiple systems.

In summary, our observations led us to conclude that there is a need for standardisation of the descriptive notation for clinically relevant pathological processes. The descriptive notation would need to describe relevant biopsychosocial domains and would need to be language independent. In short, this would need to be a clinical symbolic notation with each symbol capturing clinically relevant logical relationships or constructs. This would solve a number of the problems identified including the ability for the international clinical community to rapidly communicate important concepts (e.g. during public health emergencies) as well as facilitating cross-speciality communications. The symbolic notation would also facilitate education and training.

#### Standardised Symbolic Clinical Notation as a Tool for Precision Medicine

In the final part of the next steps, we look not to the immediate future but to the intermediate and even long term future and consider the benefits of a languageless clinical symbolic notation and how this may be reached. This follows on from the diagram mapping that has been of practical benefit in communicating our results.

In the pandemic, diagnostic overshadowing is possible given the multi-system nature of COVID-19. Diagnostic overshadowing as a term was originally used in the field of Learning Disability and in other areas of mental health to describe the attribution of physical illness to mental illness (Shefer et al, 2014). We use the term diagnostic overshadowing in a more general sense to describe the attribution of one illness to another regardless of whether this is a physical or mental illness and as a result of factors including symptom overlap, incapacity (e.g. to provide a history) and urgency of treatment for main differential (e.g. in the resuscitation setting). The occurrence of diagnostic overshadowing in the original context is likely to represent the challenges of hierarchical as opposed to multiaxial diagnostic systems and an analogy can be drawn with polycoagulopathy.

The consideration of a polycoagulopathy as opposed to a unitary coagulopathy presents the same challenges both clinically and in the research setting. In both the research setting this also raises the question of how we should compare the pairing of inductive with deductive reasoning versus inductive with abductive reasoning. In clinical medicine we are dealing with the whole, also referred to as holistic or integrated. In effect we are dealing with systems biology and this is done routinely in clinical practice. When dealing with this in clinical practice, the clinician will readily understand what we mean by clinical intuition and that clinical medicine is an art as well as a science and a technology (application of clinical knowledge). The clinician must quickly deal with clinical matters by means of the clinical assessment which involves the history, examination and an analysis of the results of the various investigations. The clinician thereby builds up an intuitive understanding of the whole picture, with an often understated but necessary reference to the systems biology of the patient. Consider a case in which there is an acute deterioration in renal function. How will this impact on the clearance of medication? How will this impact on the history and in what way will that impact on the interpretation of other aspects of the history? How will this in turn impact on the evaluation of cognition as a marker of mind and brain function? These few questions hint at the many layers of complexity that the clinician must routinely evaluate.

There is a hidden layer in communication whereby the many links that the clinician makes in an instant are hidden from view. The clinician may be unaware of these tiny steps as they arrive quickly at the correct diagnosis. The question of how the mind runs through many tens or even hundreds of steps to arrive at an apparently simple answer to a question is addressed elegantly by (Hofstadter, 1979). On encountering a clinical problem, the clinician’s mind races intuitively through many steps. The years and decades of meticulous study, training and experience will lead the mind’s direction of inquiry almost imperceptibly but if that is unpacked we may expect to see a depth of understanding and consideration of systems biology.

We may draw an analogy to the classic studies of psychology Professor Van der Groot who investigated the abilities of chess players. His subjects ranged from novices to former world champions such as Alexander Alekhine and Dr Max Euwe. Van der Groot found that the expert player was able to recall substantially more of chess positions than novices even after a short glance at the board (de Groot, 1965). Subsequent work by (Chase and Simon, 1973) determined that brain divided the chess positions into chunks and that stronger players utilised ‘larger’ chunks. As per (Hofstadter, 1979), (de Groot, 1965) and (Chase and Simon, 1973), Dr Sigmund Freud and Dr Carl Jung both understood the importance of symbols in the unconscious mind (Freud, 1913)(Jung, 1964). We hereby propose that a clinical symbolic notation would provide clinicians with a medium for expressing the hitherto unconscious aspects of their clinical reasoning and enable an improved ability to train and develop this intuition.

We have in the current system a move from the analysis of the data to the diagnosis with intermediate written accounts predominantly in the form of a generalised written language albeit with specialist terminology. Spoken and written language is general in function and although a specialised clinical terminology exists, this causes the clinician to depend on verbal intelligence to communicate with writing as a correlate. The brain is capable of a range of modes of thinking which includes verbal and non-verbal. The generation of a symbolic clinical language would offer not only a shorthand method of communicating but over time as this symbolic language is developed, it would enable the clinician to develop in several ways. Firstly the clinician would increase their exposure to clinical data-sets as in each clinical encounter there is an opportunity to produce a symbolic structure (collection of symbols) that correlates with the clinical assessment. Although there are many steps in-between which would require further development, this enables in principle the possibility of service level evaluation according to abstract representations of clinical presentations. With the construction of databases, the clinical assessment could also be searched against other similar presentations which have been annotated and linked for instance to the relevant clinical literature.. Recording a history would involve documenting with symbols, whilst retaining the ability to record text and where special training would be required to enhance the symbolic recording. The history taking (including documentation) would become more efficient and look different to current practice, enriching the level of detail possible and opening up new visas to contribution to this symbolic language by the international scientific community. A physician on one side of the world would be able to provide clinical expert advice to another physician on the other side of the world who may have limited bandwidth in their internet connection and where there is no spoken language in common.

#### The Application of Symbolic Computation in Precision Medicine

With clinical symbols having meaning and attached to a set of logical rules including their interrelationship with other symbols, they have computable properties. Provided there is a sufficient set of symbols and that they become routinely used in clinical practice, then the computable properties of these symbols offers up completely new approaches in clinical medicine. The clinician may without recourse to spoken or written language, assess their clinical intuition according to their interaction with an abstract symbolic set.

Precision medicine is the application of “individual variability in genes, environment, and lifestyle” into the treatment of cancer and then into other areas of medicine (Collins et al, 2016). With the expansion of knowledge resulting from other branches of science, precision medicine can be interpreted more broadly as the application of an ever increasing body of scientific knowledge to medicine. This knowledge occupies the space between the clinical data and the diagnosis, this intermediate zone that the clinician may also deal with intuitively. With the abundant scientific data available, it is possible to answer questions about the data computationally and there is no shortage of questions that can be asked. However it is this intermediate zone between clinical data and diagnosis and the intersection with the vast amount of scientific data that provides the most clinically relevant area. A clinical symbolic notation allows this area to be mapped out continuously. If this were mapped out on a venn diagram with the intermediate zone and the total body of biological science data, then the intersection would be an area of particular interest. This intersection would be an area where symbolic computation solutions would be expected to develop more rapidly given the potential for clinical application. Additionally areas of basic science which were able to demonstrate links to this intersection would potentially be more able to secure research funding. A directory linked to this intersection would enable clinical groups to more easily establish connections with relevant researchers and vice versa.

The symbolic notation would enable many of the missing steps in clinical thinking to be captured as a practical symbolic language would exist to capture this with the aid of simple software tools such as autocompletion, frequently used structures, keyboard shortcuts and menus. Decision-support tools would reach a more sophisticated level as the computation reaches closer to biological reality and thereby becomes more ecologically valid.

Education would be transformed. (Sadler and Regan, 2019) have written about the success of AlphaZero, an artificial intelligence chess program that was programmed to teach itself to play chess and which has achieved a considerable degree of success in this endeavour playing with human-like strategic manoeuvres which at times are incomprehensible, long before reaching positions reminiscent of some of the greatest strategic chess players such as Anatoly Karpov. (Sadler and Regan, 2019) have demonstrated in their work that valuable insights can be obtained for human chess players to learn from and improve their play based upon an analysis of the games of AlphaZero. Chess is a useful analogy for medicine as both involve considerable cognitive demands and where methodical study can improve performance. To draw an analogy, if similar achievements were possible for programs that were applied to clinical symbolic notation datasets then medical students of the future could receive training from software programs with deep insights into clinical medicine. By assessing clinical intuition using symbolic structures, algorithms could be developed which are able to ever more effectively train the doctors of the future to improve their clinical intuition in ways which could potentially be incomprehensible to humans or even tailored so that they are understandable. This would enable the possibility of accelerated programs which is of direct relevance to the early graduation of medical students that we have witnessed in this pandemic.

Lastly it is important to note that first and foremost this notation is for the frontline clinician and the primary driver should be that the notation in the most basic format should be intuitive enough for the clinician to be able to use including students. While silos may develop, they would be distinct from the basic notational system which would be universally available.

#### Pathways and Barriers to a Standardised Symbolic Clinical Notation

The road to a languageless clinical symbolic notation faces many challenges but also partially mapped routes. The field of semiotics which is closely related to the philosophy of pragmatism is the study of symbols (Chandler, 2017). With the development of Emojis (Danesi, 2016), while practically useful and bringing benefits to mass communication, demonstrates the success of such an endeavour. The use of Emoji’s has been closely aligned with the use of mobile devices and social media. Standardisation of clinical practice has precedents. Thus Karl Jaspers transformed the practice of psychiatry by developing the field of psychopathology after he observed different approaches to psychiatric history taking (Jaspers, 1997). The International Statistical Institute was responsible for the development of the International Classification of Diseases (WHO, 2020) demonstrating that other disciplines can have a profound influence on the course of medicine.

There are also profound barriers to the development of a standardised symbolic clinical notation SSCN. The most important principle is that any symbolic clinical notation system becomes a de facto language and that in turn is dependent on a social contract. Thus the SSCN cannot develop simply on the basis of a range of potential benefits but occurs secondary to a cultural change. In modern healthcare practice, such changes in culture require funding as IT changes to support sustainable infrastructure requires investment. This is usually tied up with performance data and so realistically these changes are likely to happen when the utilisation of a standardised symbolic clinical notation becomes a billable activity. This in turn requires considerable planning. Any alternatives would require creative funding or open-source approaches.

A different route is for the SSCN to be developed by related groups such as in research settings. (Kuhn, 1962) comments that individual scientific communities develop their own specialised languages and so if this is developed by smaller communities, the notation may be quite specialised and perhaps not useful to clinicians. Indeed such notations exist within chemistry for instance. This is distinct from the notation that is required in clinical medicine which covers multiple ontological levels and requires different logical rule sets. The solution may to better define the transitional zone and for the research communities to work towards a portion of their symbolic notation which can be understood and used by clinicians. This would provide a potential route to research funding. If sufficient research groups adopted this approach, the transitional zone would slowly emerge and the foundation would be cemented by congresses which bring together the different groups to bring about standardisation. From this point it may then be possible to build a case for billable activities and subsequent mainstreaming.

There are other routes also. The use of social media may enable the development of apps to experiment with this approach for clinicians using social media. Additional the use of GitHub could draw upon the international developer community to create open source solutions that address many of the challenges of an SSCN. The challenges include the construction of a symbolic system which may eventually need to describe thousands of concepts and which therefore may require large numbers of pixels and standardised approaches for graphical design (e.g. rules for utilisation of a grid-like framework).

## Summary

We conclude with a summary of the main findings in this paper and determine from our evidence that SARS-COV2 infection leads to a viral clotting fever syndrome with high mortality. COVID-19 is a polycoagulopathy with multiple clotting mechanisms (MCM’s) causing serious clinical thromboembolic sequalae. Our findings are based on a population that was hospitalised and where there was evidence of more advanced age. Fever occurred in most but not all of the patients with this presentation and therefore the clotting fever syndrome is an indication that if both fever and clotting events occur that COVID-19 should be considered as a differential. The absence of fever or thromboembolic events should not discount COVID-19 as a differential and the respiratory as well as other manifestations contribute to a variable presentation. The D-dimer is a key prognostic indicator in COVID-19 and bears special significance as a marker of underlying thromboembolic events as per our results. In the most severe cases, the D-Dimers were elevated up to some 200-fold over the upper limit of the reference range and had significant clinical correlates.

Overall we found an average of 1.3 thromboembolic events per patient but these were predominantly arterial rather than venous in nature albeit where we had classified pulmonary emboli as arterial rather than venous on the basis of the vessel anatomy rather than the consideration of the flow of blood towards or away from the heart. This is particularly relevant in COVID-19 given the distribution of ACE-2 receptors in the smooth muscle cells which in turn are more densely arranged in the arterial vasculature. Of note is that we found not a single case of pulmonary vein thrombosis in the 56 main papers whilst pulmonary arterial emboli were found in abundance.

We found that there were five main groups of arterial thromboembolic events according to arterial territories – splanchnic arteries, coronary arteries, pulmonary arteries, the femoral and other arteries of the lower limbs as well as the cortical and subcortical arterial supply but particularly the middle cerebral arteries. The involvement of these territories leads to characteristic signs and symptoms which are well-established in the literature and where further work can lead to clinical algorithms and the development of public health messages with sufficient evidence. We also identified carotid artery thromboemboli in association with strokes and many authors drew attention to this important finding. Thrombi were also found in the aorta and the heart. We found sporadic reports of the involvement of the veins with the exception of the deep veins in the lower limbs which were more frequently identified. Of special note is the occurrence of thromboembolic/clotting events in association with medical devices which we use broadly to include catheters, ECMO centrifuges and dialysis machines. The occurrence of upper limb DVT’s was predominantly associated with catheters.

A combination of more general factors in the setting of either mild or severe illness may also contribute. The insensible fluid loss from diarrhoea and vomiting may lead to dehydration and electrolyte disturbances. Periods of immobility may increase the risk of a lower limb DVT. Hypokalaemia and hyponatraemia may both predispose to atrial fibrillation which in turn results in cardiac muscle stasis and generation of thrombi and subsequent thromboembolic events. The disruption of the RAA system may impact on the correction of these disturbances.

On analysis of a subset of papers, we were able to quantify other findings albeit on the basis of a small sample size which limits our ability to generalise these findings and suggests that these findings need to be further validated. We found sex differences amongst the occurrence of arterial thromboembolic events as well as differences in mortality. We draw special attention to the occurrence of acute limb ischaemia in COVID-19 which is associated with a particularly high mortality albeit with evidence of selection bias in the data we analysed and which merits further attention.

The qualitative analysis identified the general clinical findings with an experiential (clinical) account. Thus clots were described in unusual locations, occurring in multiple territories and with unusual findings such as desert foot. During interventions, the thromboemboli were found to be friable, breaking away and further embolising into new territories. Repeated occlusion of arteries was identified after recanalisation and in one case the clot was altogether undetachable from the arterial wall necessitating an alternative surgical procedure. Thromboemboli occurred despite prophylactic anticoagulation. Patients would present with serious clinical thromboembolic events after being either asymptomatic or else with an apparently mild course of the illness.

In terms of the validated abridged thematic analysis, we were able to assess the content validity of the proposed aetiological mechanisms by means of an examination of the literature. This was not limited to the papers screened in an exploratory literature search but also with reference to the other papers we had examined for the purposes of this scoping review. By means of an iterative process utilised in thematic analysis we were then able to assemble the selected mechanisms into a taxonomy. Furthermore we were able to translate the taxonomy into a rudimentary symbolic representation, a process which may be of use to other clinicians and researchers investigating this field of enquiry as the model can be extended and refined.

In theoretical terms, we utilised Virchow’s triad as a means of structuring the taxonomy. We were able to confirm overarching themes of hypercoagulability, vascular wall involvement and to a lesser degree stasis. The primary pathology would appear to be the viral invasion of the endothelium which is mediated via the ACE-2 receptors which together with heparin sulfate affords the virus a route for entry into the cells. Once inside the cells, the virus appropriates the cellular machinery and escapes RNA intracellular sensing mechanisms by means of the construction of replication organelles including double membrane vesicles. The RNA sensing mechanisms typically trigger an interferon response upon successful detection and it should be noted that the interferon response or lack thereof appears critical in determining the subsequent course of the illness. An exaggerated interferon response may result in a mild illness and possibly chilblains as a dermatological finding. A delayed interferon response may result in a more severe course and this may be of more relevance to the risk of coagulopathy and the concept of immunosenescence may also be relevant.

Following the invasion of the endothelium, there is found to be endotheliitis and also various degrees of thrombosis in the small blood vessels. From our examination of the literature there appear to be many details requiring clarification but there are a few events which appear to be more robustly supported in the literature. Firstly the endothelium is specialised according to the organ in which it is located. In the lungs, the blood vessels have a physiological function of ventilation-perfusion matching whilst in the kidneys this forms part of the filtration mechanism. The viral invasion appears to degrade the function of the endothelium and it would appear that together with a disruption of the RAA axis likely resulting from the utilisation of the ACE-2 receptors, there is a subsequent degradation of ventilation-perfusion matching and glomerulofiltration if those organs are affected. The ventilation-perfusion mismatch is likely to account for the phenomenon of silent hypoxia which is of significant consequence in clinical management and hypoxaemia contributes to the risk of a coagulopathy. A disruption in glomerulofiltration results in impaired plasma filtration with significant consequences for various homeostatic mechanisms including those of the electrolytes.

The consequences of the invasion of the endothelium by the virus can also be considered in terms of thromboinflammation. The construct of thromboinflammation is well supported in the literature and it is clear that platelet activation plays a significant role in COVID-19. The key question here is the role of the endothelial glycocalyx. There is evidence of disruption of the glycocalyx in COVID-19 and this would result in a local prothrombotic environment and would lead to platelet activation. One of the components of the glycocalyx is heparin sulfate which is required for viral entry into the cells and so the disruption of the glycocalyx would be more likely secondary to viral invasion of the endothelium. A special role may be played by hyaluronan with evidence that in COVID-19 it leads to the creation of a gel in the lungs which may contribute to hypoxaemia. With the seeding of the virus in the lungs and then in the endothelium of other blood vessels, thromboinflammation would result in the generation of a thrombus. The properties of the thrombus would predispose to embolisation including a possible contribution from components of the locally disrupted glycocalyx. The arterial wall would be an ongoing source of thrombogenesis. The role of susceptibility factors is also of importance and in particular endothelial dysfunction is related to the metabolic syndrome. This likely contributes to a low grade inflammatory, prothrombotic potential and a possible vulnerability to viral invasion.

Another important aspect of the pathology is the role of the neutrophils. One clear finding is the presence of NETs and when taken together with the evidence of apoptosis and related mechanisms, a key feature of COVID-19 is exposure of intracellular material to the extracellular environment. Thus while the virus has carefully avoided detection in the intracellular space through the replication organelles, the intracellular material of the host cells is exposed. There is thus the potential for the host immune system to generate an autoimmune response. Furthermore if the tissue destruction is overwhelming and the clearance insufficient then there may be a large amount of circulating antibody complexes leading to a type-III hypersensitivity response in a minority of cases. This in turn can lead to deposition of immune complexes in the skin, kidneys and vasculature with the latter resulting in a vasculitis which can predispose to further thromboembolic events.

Finally returning to the original question in this paper, we found evidence of thromboembolic events with and without each of sepsis, DIC and ARDS.

We conclude with Tables 37 and 38 which summarise the main results of the qualitative and quantitative analysis and these should be used in conjunction with Table 35 and Figures 10-21 and with the section on the limitations of the study.

**Table 37.** Main findings of Quantitative Analysis, f – female, m – male, TE - thromboembolism.

| **Main Findings of Quantitative Analysis** |
| --- |
| 10523 patients in all 56 studies |
| 456 patients with COVID-19 and thromboembolic events |
| 586 thromboembolic events (TE's) |
| Average of 1.3 events per patient |
| Breakdown of thromboembolic events in Table 8 |
| Thromboembolism with and without ARDS |
| Thromboembolism with and without septicaemia |
| Thromboembolism with and without DIC |
| 34 studies with detailed demographic data on 119 patients |
| Average age of 119 patients - 67 |
| Average age of patients who died - 73 |
| Average age of patients who survived - 63 |
| % of deaths in group with thromboembolism - 38% |
| % of deaths in group with myocardial infarct - 36% |
| % of deaths in group with pulmonary embolism - 25% |
| % of deaths in group with splanchnic artery thromboembolism - 60% |
| % of deaths in group with acute limb ischaemia - 79% |
| % of deaths in group with DVT - 26% |
| Higher % of f v m deaths with aortic thromboembolism |
| Higher % of m v f deaths with myocardial infarcts |
| Higher % of f v m deaths with pulmonary emboli |
| Higher % of f v m deaths with splanchnic thromboemboli |
| Higher % of f v m deaths with acute limb ischaemia |
| Significance for mortality v survival with splanchnic TE's |
| 27 published values for D-Dimers with reference range |
| Mean D-Dimer of 22 x upper limit of reference range with TE's |
| 91/150 patients with COVID-19 and TE's with fever |

**Table 38.** Main Findings of Abridged Thematic Analysis, ALI – acute limb ischaemia.

| **Main Findings in Abridged Thematic Analysis** |
| --- |
| No known risk factors |
| Thromboembolic event despite anticoagulation/antiplatelet therapy |
| High in-hospital mortality |
| Asymptomatic prior to thromboembolic event |
| Cryptogenic/Without any source of thromboembolism |
| Multiterritory stroke |
| Rethrombosis |
| Mild symptoms prior to presentation |
| Minimal or no improvement after revascularisation for stroke |
| No recanalisation after one pass for stroke |
| Clot fragmentation with embolisation with intervention |
| Unusual location of clots |
| Desert foot |
| Low rate of successful revascularisation for ALI |
| Thrombosis of a graft |
| Clotting of medical devices |

## Supporting information

Diagram Mapping

## Data Availability

We have made data available.

## Acknowledgements

We would like to extend our immense gratitude to family, friends and colleagues whose support has been essential in managing the many challenges we have faced in this pandemic.

Sepsis

(Papers identified)

ACE-2 mediated pathways via lung and heart binding

6. Cytokine storm/cytokine

1. Central Nervous System

a. Direct virus nervous system injury

a. Direct Invasion of the Myocardium

a. Viral enteroneuropathy

a. Respiratory pathology

b. Respiratory failure leading to myocardial mismatch perfusion

a. Microangiopathy leading to ischaemia

7. Immobilisation/Bed rest

Alternative Complement Pathway

Hypothesis Generation

Hypothesis 1

Recommendations

Recommendation 1

