## Supplementary material for "Characterising COVID-19 as a Viral Clotting Fever: A Mixed Methods Scoping Review": Diagram Mapping

### Slide 1
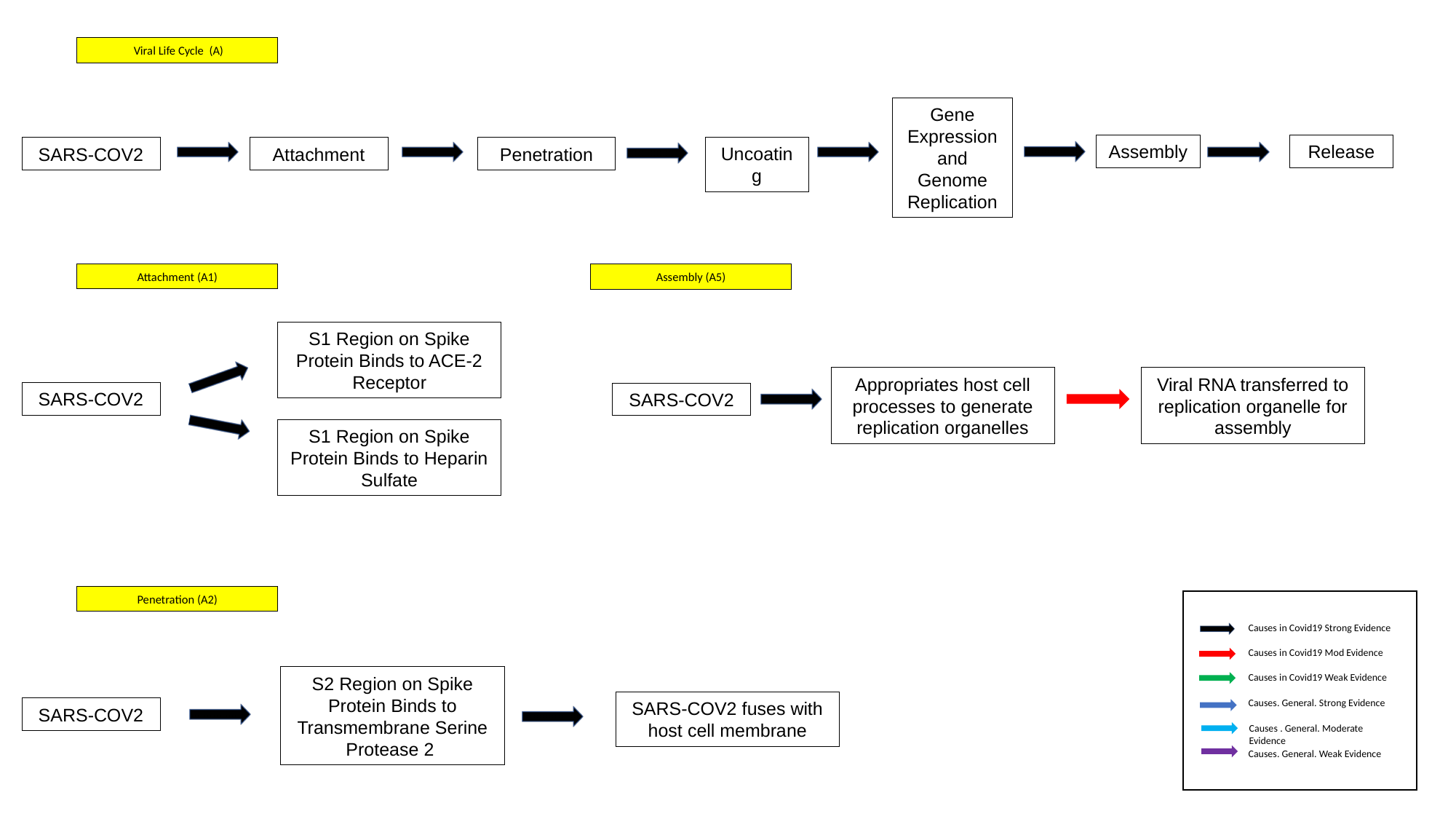

Viral Life Cycle (A)
Gene Expression and Genome Replication
Assembly
Release
SARS-COV2
Uncoating
Attachment
Penetration
Attachment (A1)
Assembly (A5)
S1 Region on Spike Protein Binds to ACE-2 Receptor
Viral RNA transferred to replication organelle for assembly
Appropriates host cell processes to generate replication organelles
SARS-COV2
SARS-COV2
S1 Region on Spike Protein Binds to Heparin Sulfate
Penetration (A2)
Causes in Covid19 Strong Evidence
Causes in Covid19 Mod Evidence
Causes in Covid19 Weak Evidence
S2 Region on Spike Protein Binds to Transmembrane Serine Protease 2
Causes. General. Strong Evidence
SARS-COV2 fuses with host cell membrane
SARS-COV2
Causes . General. Moderate Evidence
Causes. General. Weak Evidence

### Slide 2
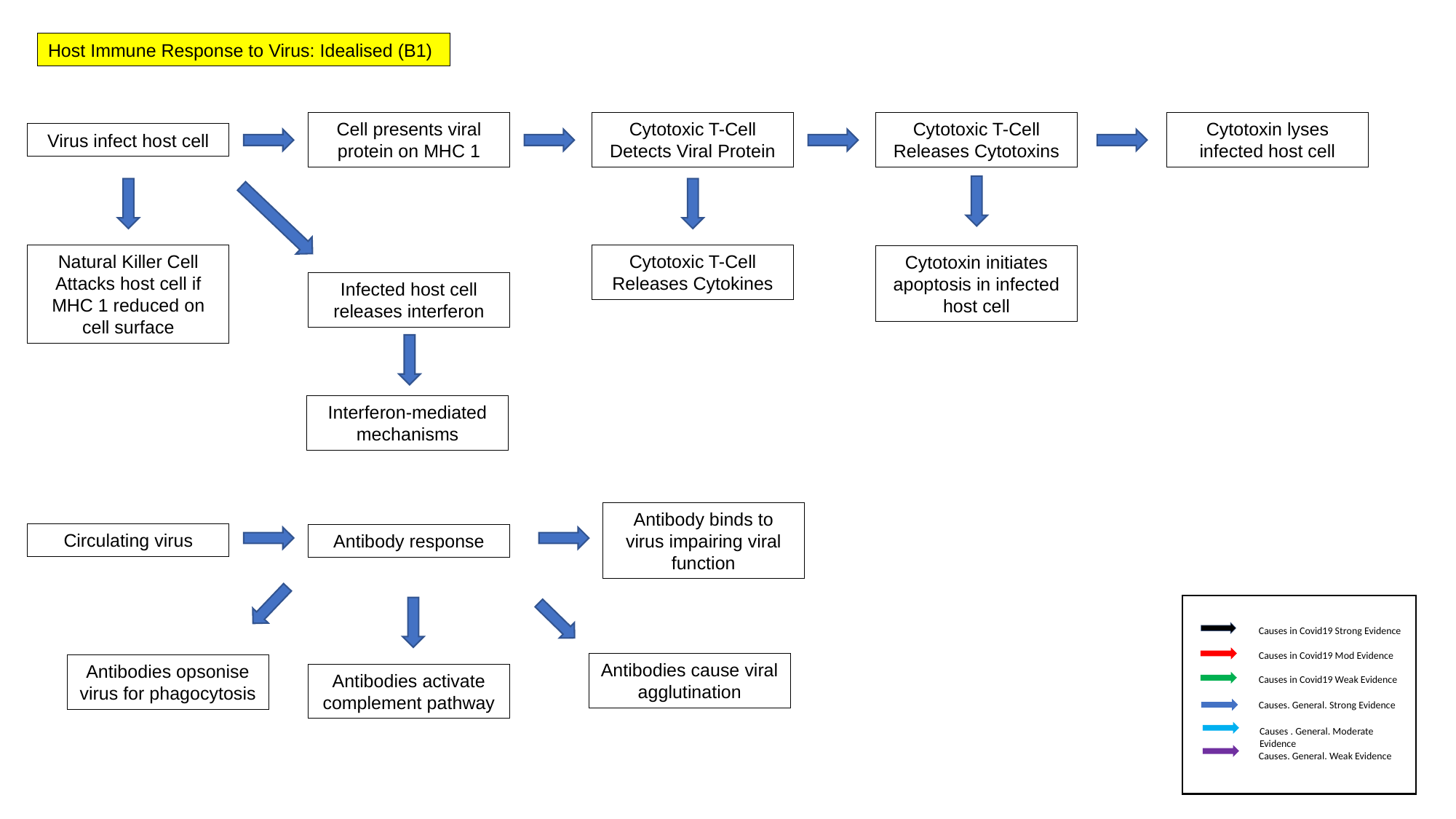

Host Immune Response to Virus: Idealised (B1)
Cell presents viral protein on MHC 1
Cytotoxic T-Cell Detects Viral Protein
Cytotoxic T-Cell Releases Cytotoxins
Cytotoxin lyses infected host cell
Virus infect host cell
Cytotoxic T-Cell Releases Cytokines
Natural Killer Cell Attacks host cell if MHC 1 reduced on cell surface
Cytotoxin initiates apoptosis in infected host cell
Infected host cell releases interferon
Interferon-mediated mechanisms
Antibody binds to virus impairing viral function
Circulating virus
Antibody response
Causes in Covid19 Strong Evidence
Causes in Covid19 Mod Evidence
Antibodies cause viral agglutination
Antibodies opsonise virus for phagocytosis
Antibodies activate complement pathway
Causes in Covid19 Weak Evidence
Causes. General. Strong Evidence
Causes . General. Moderate Evidence
Causes. General. Weak Evidence

### Slide 3
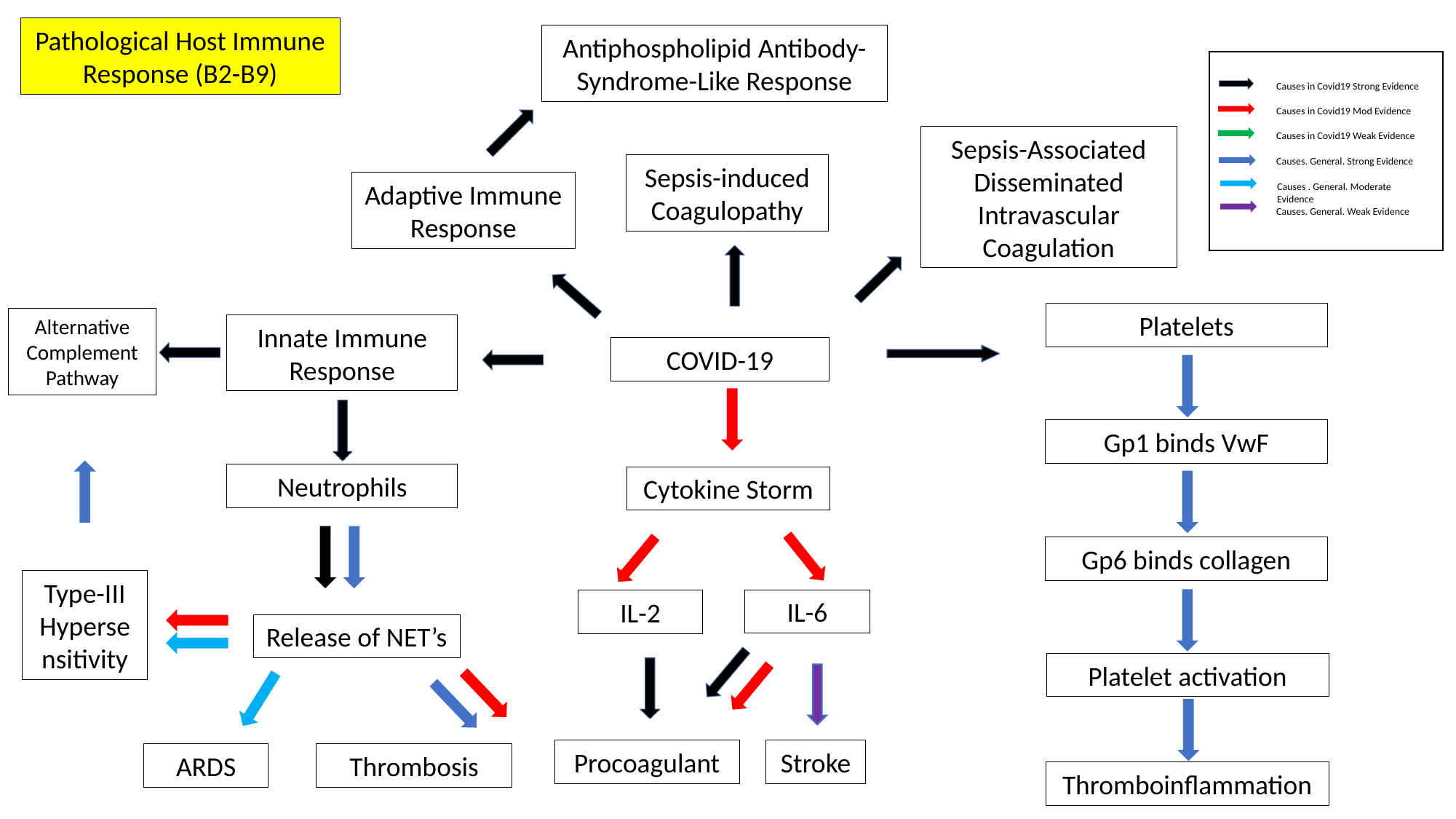

Pathological Host Immune Response (B2-B9)
Antiphospholipid Antibody-Syndrome-Like Response
Causes in Covid19 Strong Evidence
Causes in Covid19 Mod Evidence
Causes in Covid19 Weak Evidence
Sepsis-Associated Disseminated Intravascular Coagulation
Causes. General. Strong Evidence
Sepsis-induced Coagulopathy
Adaptive Immune Response
Causes . General. Moderate Evidence
Causes. General. Weak Evidence
Platelets
Alternative Complement Pathway
Innate Immune Response
COVID-19
Gp1 binds VwF
Neutrophils
Cytokine Storm
Gp6 binds collagen
Type-III Hypersensitivity
IL-6
IL-2
Release of NET’s
Platelet activation
Procoagulant
Stroke
ARDS
Thrombosis
Thromboinflammation

### Slide 4
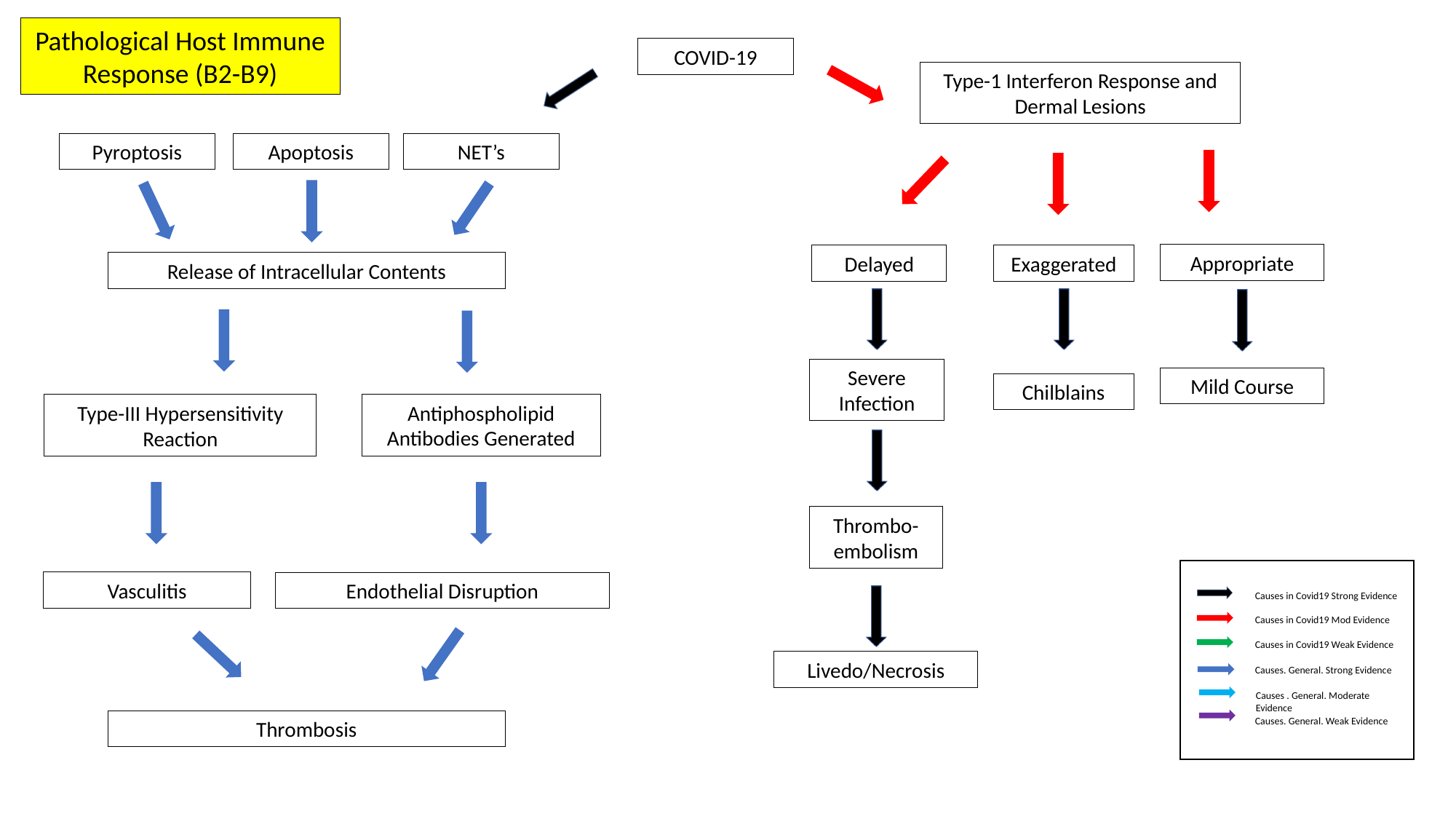

Pathological Host Immune Response (B2-B9)
COVID-19
Type-1 Interferon Response and Dermal Lesions
Pyroptosis
Apoptosis
NET’s
Appropriate
Delayed
Exaggerated
Release of Intracellular Contents
Severe Infection
Mild Course
Chilblains
Antiphospholipid Antibodies Generated
Type-III Hypersensitivity Reaction
Thrombo-
embolism
Vasculitis
Endothelial Disruption
Causes in Covid19 Strong Evidence
Causes in Covid19 Mod Evidence
Causes in Covid19 Weak Evidence
Livedo/Necrosis
Causes. General. Strong Evidence
Causes . General. Moderate Evidence
Causes. General. Weak Evidence
Thrombosis

### Slide 5
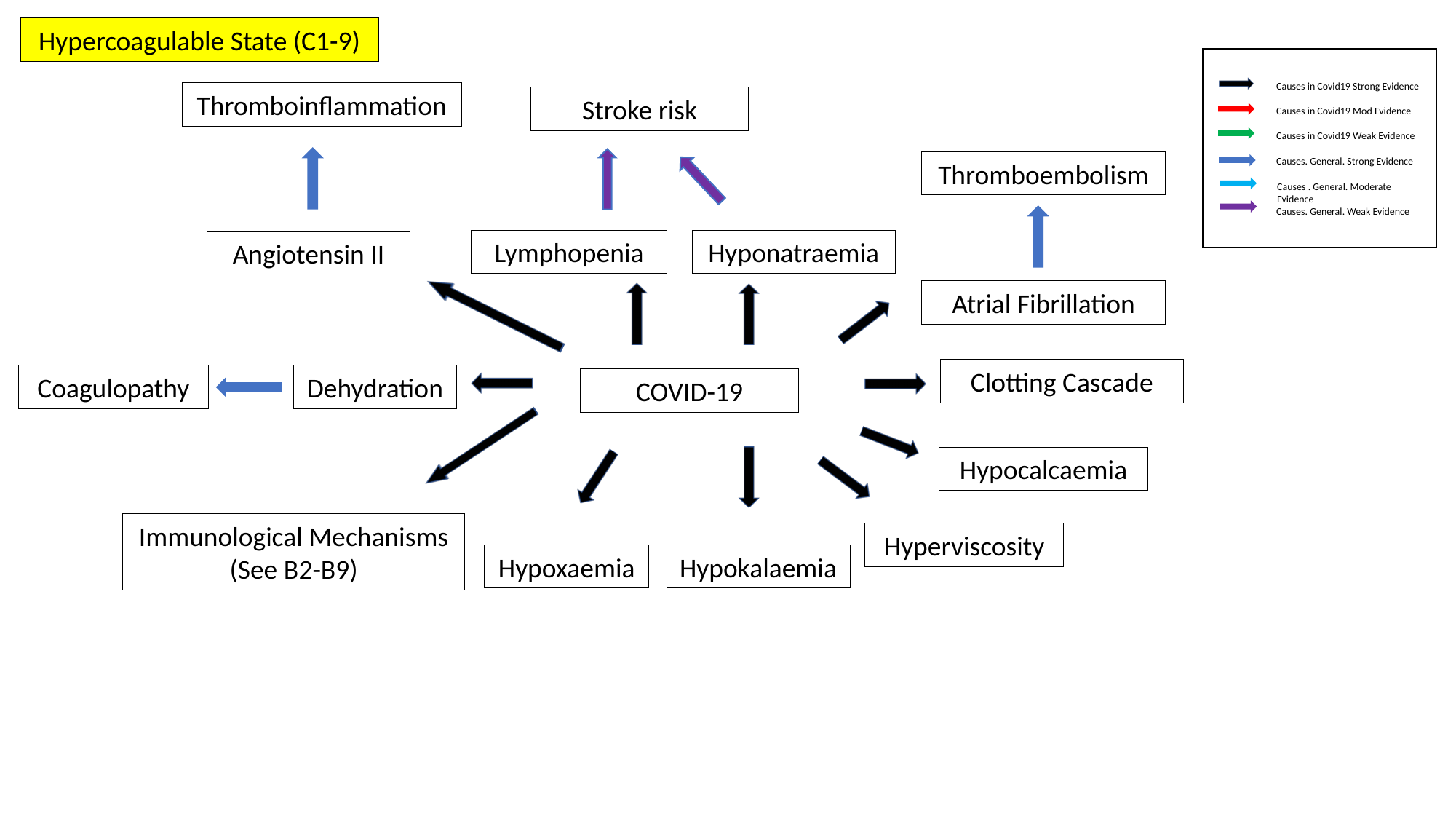

Hypercoagulable State (C1-9)
Causes in Covid19 Strong Evidence
Thromboinflammation
Stroke risk
Causes in Covid19 Mod Evidence
Causes in Covid19 Weak Evidence
Causes. General. Strong Evidence
Thromboembolism
Causes . General. Moderate Evidence
Causes. General. Weak Evidence
Lymphopenia
Hyponatraemia
Angiotensin II
Atrial Fibrillation
Clotting Cascade
Coagulopathy
Dehydration
COVID-19
Hypocalcaemia
Immunological Mechanisms (See B2-B9)
Hyperviscosity
Hypoxaemia
Hypokalaemia

### Slide 6
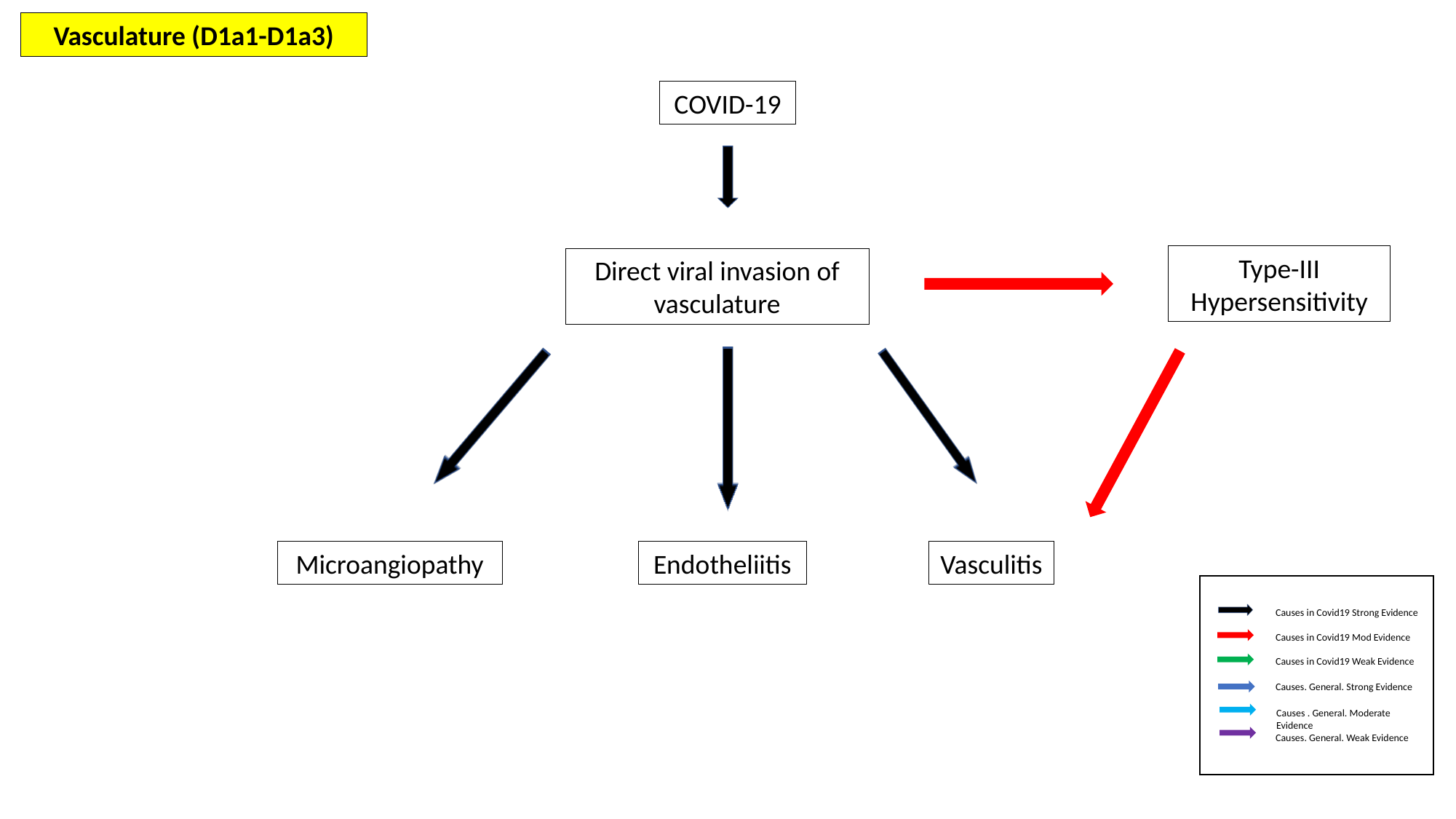

Vasculature (D1a1-D1a3)
COVID-19
Type-III Hypersensitivity
Direct viral invasion of vasculature
Microangiopathy
Endotheliitis
Vasculitis
Causes in Covid19 Strong Evidence
Causes in Covid19 Mod Evidence
Causes in Covid19 Weak Evidence
Causes. General. Strong Evidence
Causes . General. Moderate Evidence
Causes. General. Weak Evidence

### Slide 7
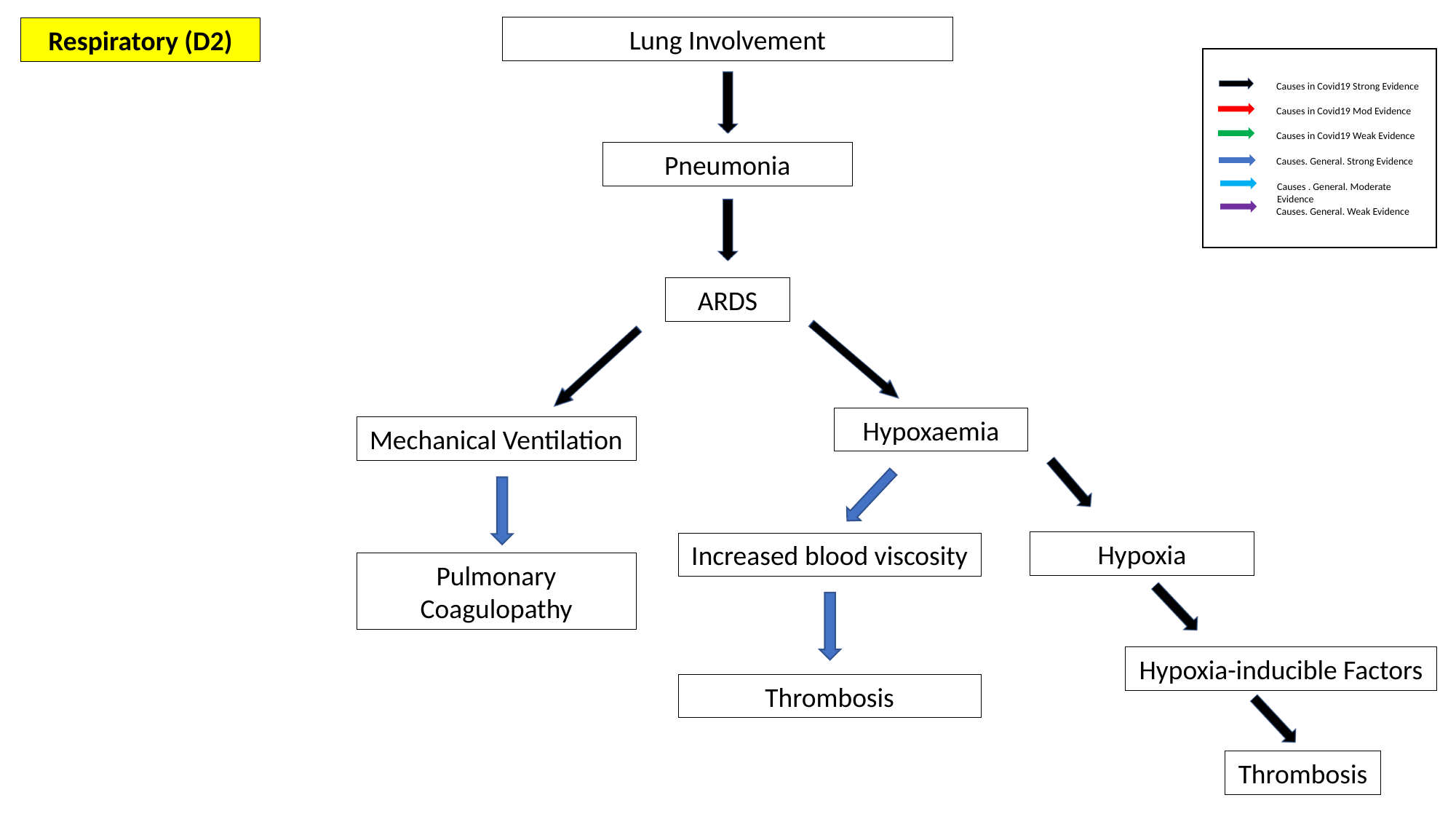

Lung Involvement
Respiratory (D2)
Causes in Covid19 Strong Evidence
Causes in Covid19 Mod Evidence
Causes in Covid19 Weak Evidence
Pneumonia
Causes. General. Strong Evidence
Causes . General. Moderate Evidence
Causes. General. Weak Evidence
ARDS
Hypoxaemia
Mechanical Ventilation
Hypoxia
Increased blood viscosity
Pulmonary Coagulopathy
Hypoxia-inducible Factors
Thrombosis
Thrombosis

### Slide 8
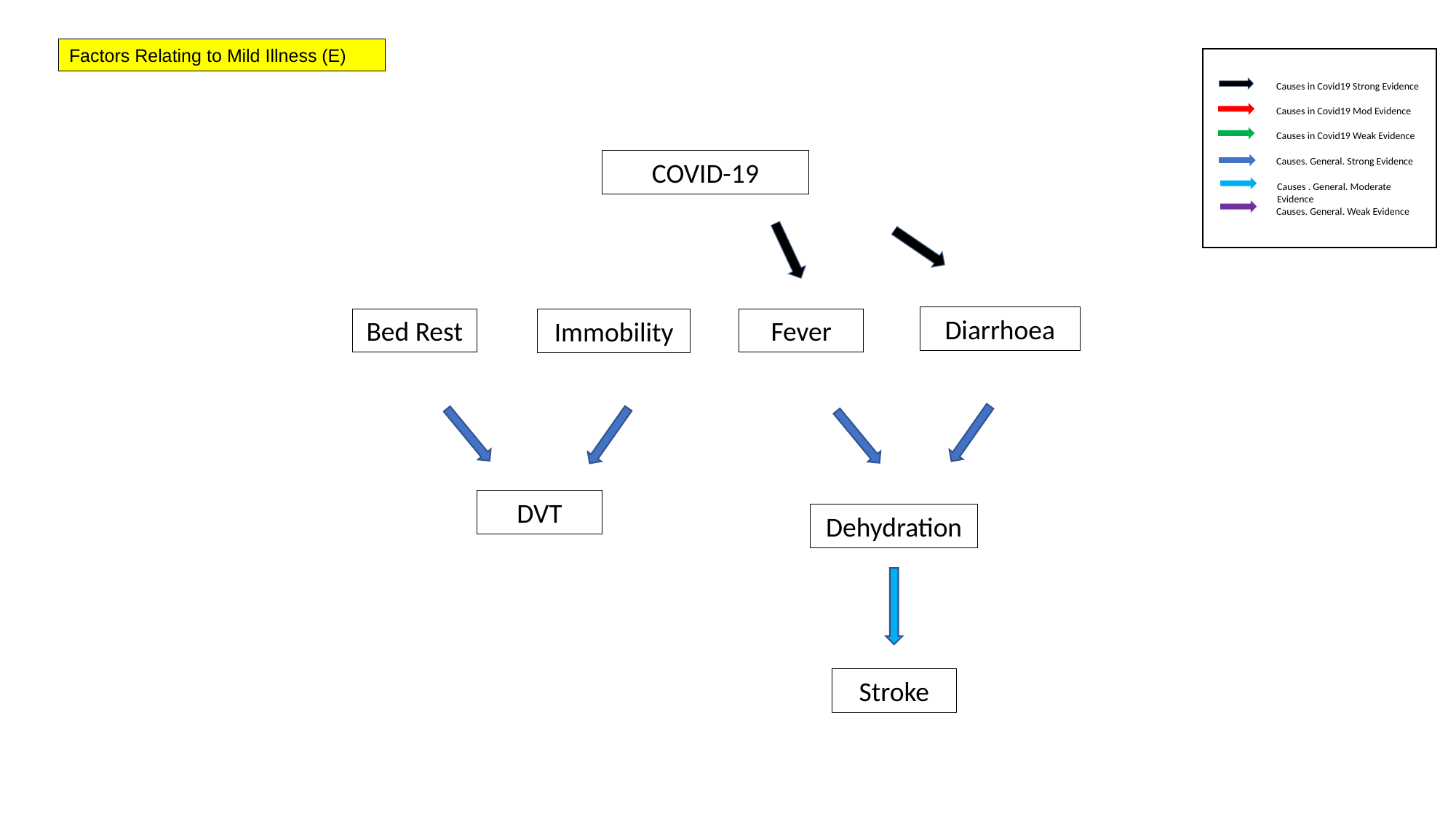

Factors Relating to Mild Illness (E)
Causes in Covid19 Strong Evidence
Causes in Covid19 Mod Evidence
Causes in Covid19 Weak Evidence
Causes. General. Strong Evidence
COVID-19
Causes . General. Moderate Evidence
Causes. General. Weak Evidence
Diarrhoea
Bed Rest
Fever
Immobility
DVT
Dehydration
Stroke

### Slide 9
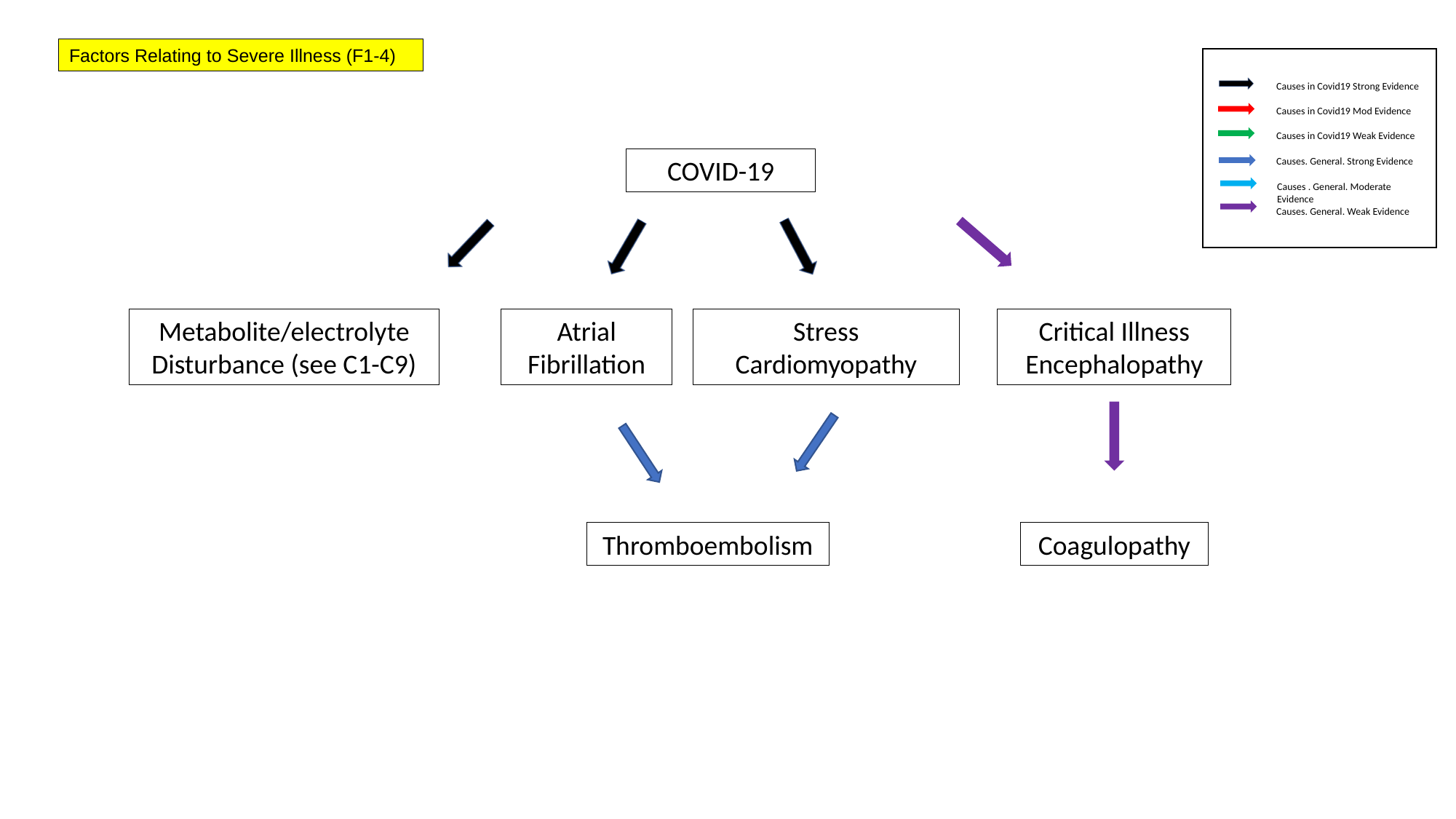

Factors Relating to Severe Illness (F1-4)
Causes in Covid19 Strong Evidence
Causes in Covid19 Mod Evidence
Causes in Covid19 Weak Evidence
COVID-19
Causes. General. Strong Evidence
Causes . General. Moderate Evidence
Causes. General. Weak Evidence
Metabolite/electrolyte Disturbance (see C1-C9)
Atrial Fibrillation
Stress Cardiomyopathy
Critical Illness Encephalopathy
Thromboembolism
Coagulopathy

### Slide 10
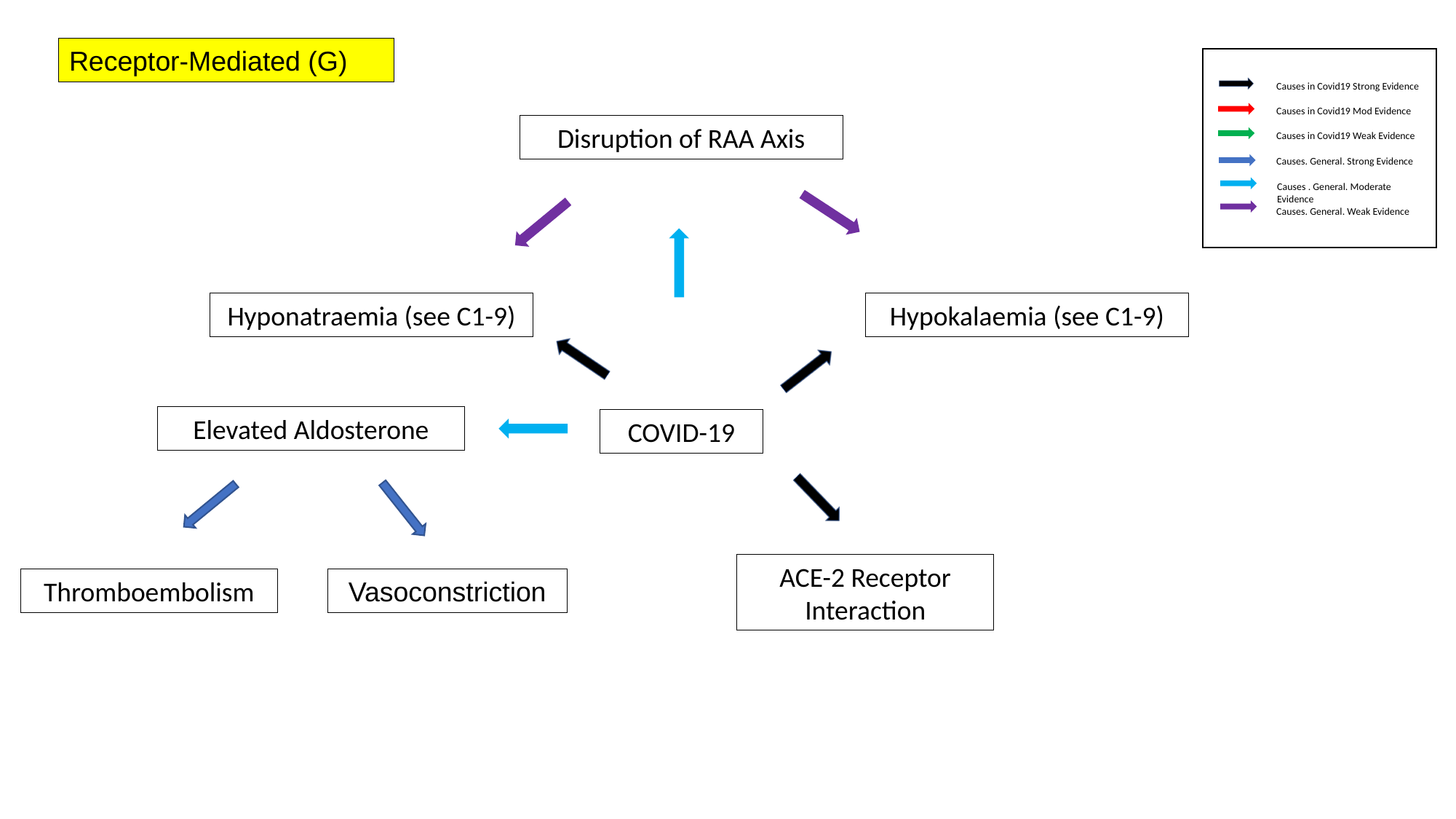

Receptor-Mediated (G)
Causes in Covid19 Strong Evidence
Causes in Covid19 Mod Evidence
Disruption of RAA Axis
Causes in Covid19 Weak Evidence
Causes. General. Strong Evidence
Causes . General. Moderate Evidence
Causes. General. Weak Evidence
Hyponatraemia (see C1-9)
Hypokalaemia (see C1-9)
Elevated Aldosterone
COVID-19
ACE-2 Receptor Interaction
Thromboembolism
Vasoconstriction

### Slide 11
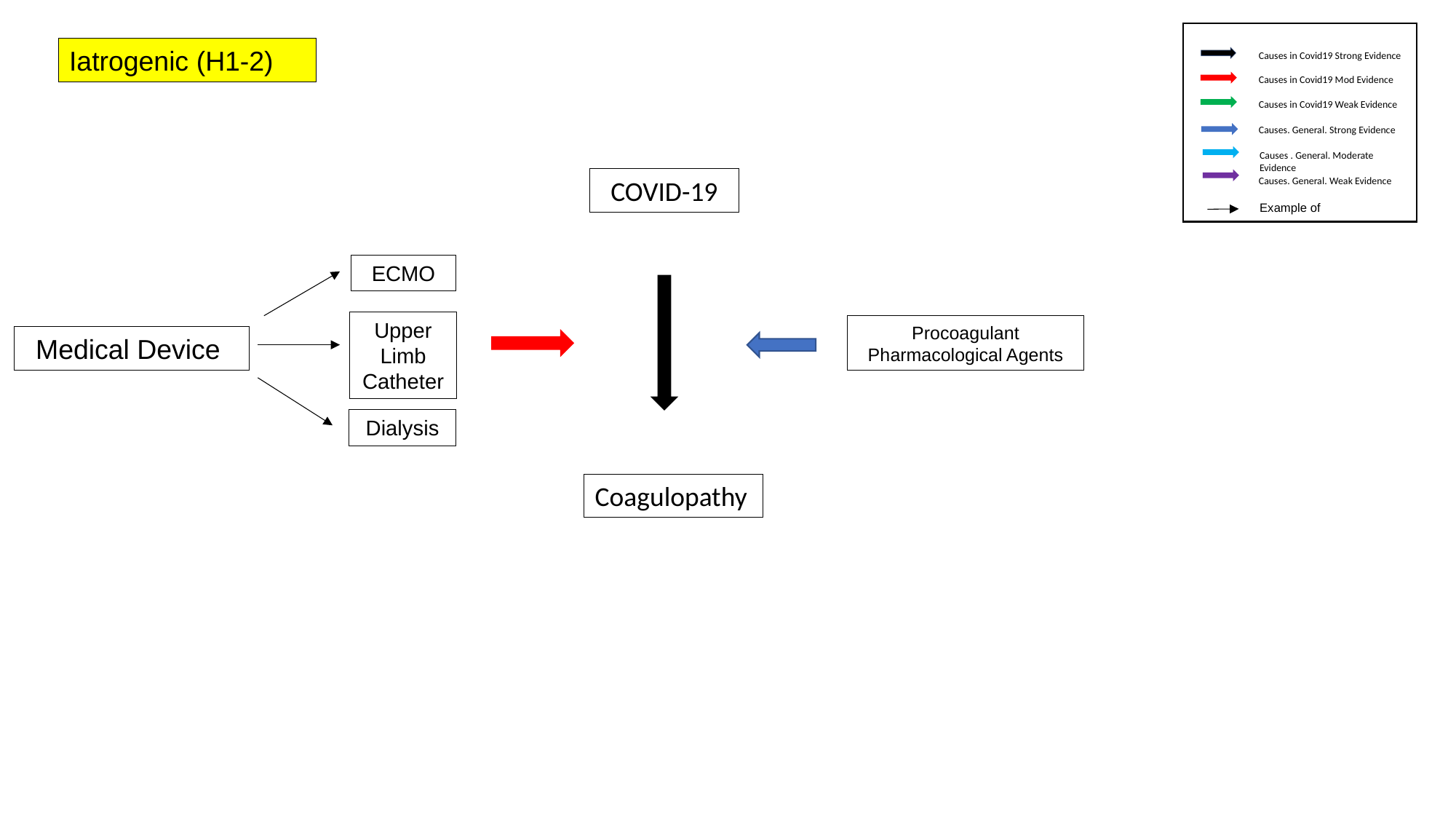

Iatrogenic (H1-2)
Causes in Covid19 Strong Evidence
Causes in Covid19 Mod Evidence
Causes in Covid19 Weak Evidence
Causes. General. Strong Evidence
Causes . General. Moderate Evidence
COVID-19
Causes. General. Weak Evidence
Example of
ECMO
Upper Limb Catheter
Procoagulant Pharmacological Agents
Medical Device
Dialysis
Coagulopathy

### Slide 12
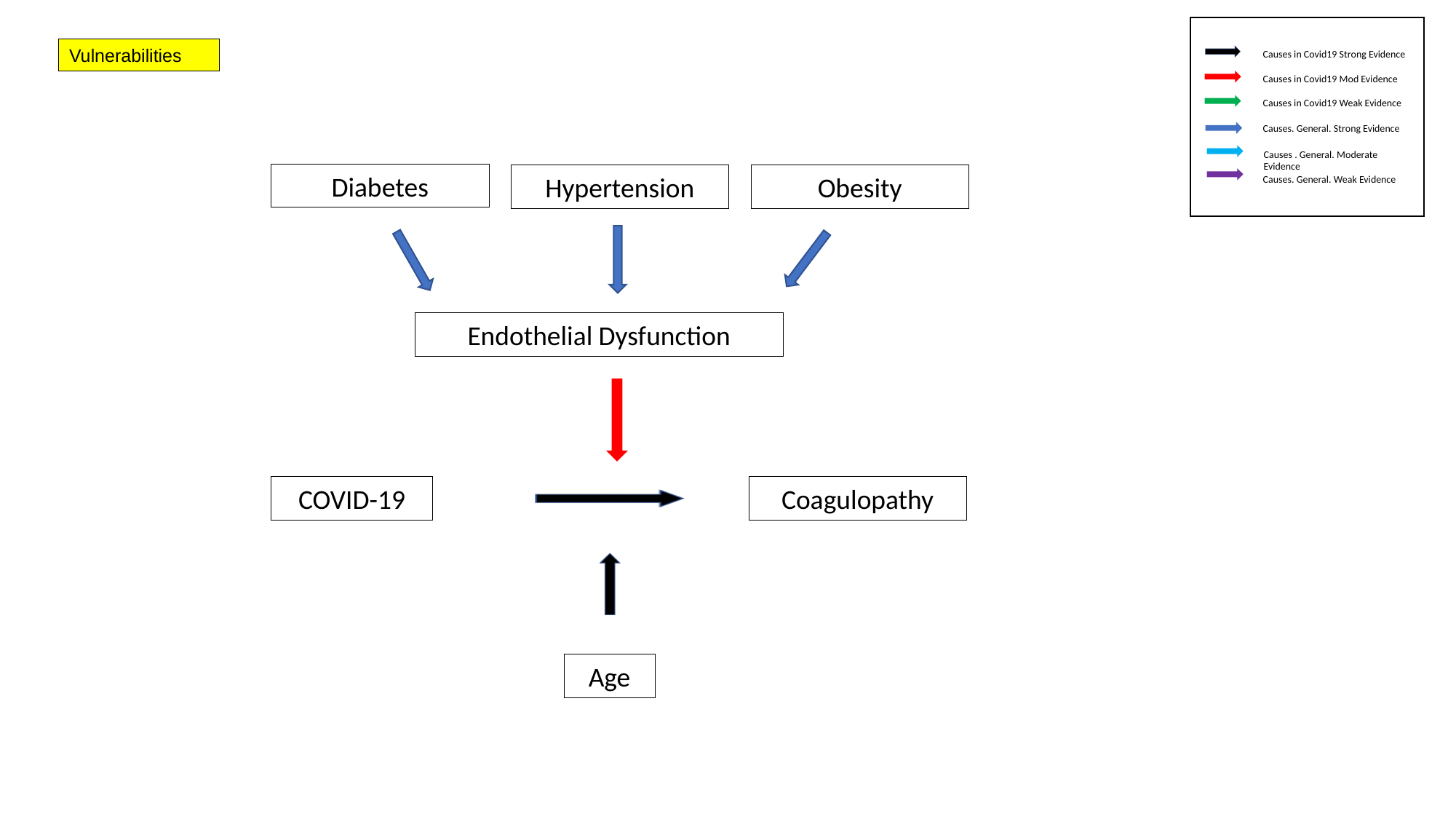

Vulnerabilities
Causes in Covid19 Strong Evidence
Causes in Covid19 Mod Evidence
Causes in Covid19 Weak Evidence
Causes. General. Strong Evidence
Causes . General. Moderate Evidence
Diabetes
Hypertension
Obesity
Causes. General. Weak Evidence
Endothelial Dysfunction
COVID-19
Coagulopathy
Age
